# Error-Controlled Decision Policies for Medical Foundation Models

**DOI:** 10.64898/2026.02.23.26346927

**Authors:** Intae Moon, Ying Jin, Mustafa Saleh, Alexander Gusev, Marinka Zitnik

**Affiliations:** Department of Biomedical Informatics, Harvard Medical School, Boston, MA, USA; Broad Institute of MIT and Harvard, Cambridge, MA, USA; Department of Statistics and Data Science, University of Pennsylvania, Philadelphia, PA, USA; Division of Population Sciences, Dana-Farber Cancer Institute, Boston, MA, USA; Kempner Institute for the Study of Natural and Artificial Intelligence, Harvard University, Allston, MA, USA; Harvard Data Science Initiative, Cambridge, MA, USA

## Abstract

To use foundation models in medicine, clinicians need decision policies that act on model predictions within an explicit error budget, such as a limit on the rate of false positive calls. High average prediction accuracy does not guarantee safe decisions, because errors can concentrate among the patients whose predictions directly guide care and lead to harm and inefficient use of healthcare resources. Here we introduce StratCP, a stratified conformal prediction approach that decides, for each patient, whether a foundation model prediction can directly guide care or whether the patient should be deferred to further evaluation. StratCP selects patients whose predictions can be acted on while controlling the false discovery rate at a level set by the user. For deferred patients, it returns prediction sets that contain the true disease state at the target rate to guide confirmatory testing or expert review. When clinical guidelines define relationships among disease states, StratCP uses a utility graph to form clinically coherent prediction sets while maintaining coverage. We evaluate StratCP with foundation models for ophthalmology, neuro-oncology, and longitudinal health records across diagnosis, biomarker, operational, laboratory, and survival prediction tasks. StratCP controls the false discovery rate among selected patients and achieves the target coverage among deferred patients, whereas standard conformal prediction exceeds the error budget. When a predictor is applied to a new cohort, calibrating StratCP on as few as 50 labeled slides from that cohort maintains error control and prediction-set coverage without retraining. In an independent neuro-oncology cohort, StratCP selects predictions of IDH status and integrated diagnosis from H&E slides that meet the error budget, showing potential for earlier assessment during routine pathology workup. StratCP provides decision policies that specify, with controlled error, when foundation model predictions can guide care.

## Main

Medical foundation models (FMs) have been developed across diverse clinical data modalities, including retinal imaging [1], whole-slide histopathology [2, 3], and longitudinal electronic health records (EHRs) [4–7]. Several FMs have progressed to prospective evaluation and integration within health system workflows [8–12]. Routine clinical use, however, shifts the objective from benchmark performance to *actionable* outputs that support safe decisions within prespecified error budgets. Beyond how accurate a model is on average, clinicians need to know when it is appropriate to act on its predictions. An FM predictor that cannot meet the required error rate for all patients may still meet it for a subset of patients, which motivates selective action instead of acting on every prediction or on none. Clinicians therefore need explicit guidance on when to act, when to defer, and which follow-up to pursue [13–15]. Without such decision policies, model predictions may lead to unnecessary or harmful interventions, delay appropriate care, and waste limited diagnostic resources [16, 17].

Medical FMs typically produce point predictions and do not provide uncertainty estimates to indicate when a prediction is reliable enough to support a clinical action [13, 14, 18]. A small number of recent FMs report uncertainty through dedicated uncertainty heads [19] or model ensembles [20], but these uncertainty scores can be miscalibrated and rarely come with guarantees tied to an error budget set by the user [21–23]. Such guarantees are especially important when FM predictors are reused across cohorts or clinical settings, because performance in the training cohort does not necessarily establish performance in the patient population where the model is used [24, 25]. Conformal prediction (CP) is a distribution-free approach that addresses this need by building prediction sets with finite-sample coverage guarantees from the point predictions of a model [26, 27]. Given a fixed predictor and a small labeled dataset from the setting where the model is used, CP constructs prediction sets post hoc, without retraining, that contain the true label at a coverage level set by the user, for example for 95% of patients. This guarantee has motivated the use of CP in clinical decision support [28–33]. The guarantee of CP, however, is marginal, meaning that it holds on average over all patients and not within a subset of patients selected for a clinical decision.

Reliable decisions on whether to act on FM predictions or defer require two guarantees. Among patients whose predicted disease status is used to guide care, the fraction of incorrect predictions must stay within the error budget. Among deferred patients, prediction sets must achieve the target coverage so that they can guide confirmatory testing or expert review. CP provides neither guarantee, because marginal coverage over all patients does not ensure error control or coverage within subsets of patients considered for clinical decisions [34, 35]. For example, 95% marginal coverage over all patients does not imply that 95% of the predictions selected for action are correct. At a given error budget, the practical value of a decision policy also depends on how many patients it can select for action while maintaining error control.

To provide these guarantees, we introduce StratCP, a stratified conformal prediction approach that decides, for each patient, whether an FM prediction can be acted on or whether the patient should be deferred for follow-up evaluation. In the action arm, StratCP selects patients whose predictions can guide care while controlling the false discovery rate (FDR), the expected fraction of incorrect predictions among selected patients, at a level set by the user, such as 5%. The number of selected patients measures how far the model can support direct action at this error level. In the deferral arm, StratCP returns prediction sets with coverage conditional on deferral, so that among deferred patients the set contains the true disease status at the rate set by the user. StratCP builds on conformal selection [36] and selection-conditional conformal prediction [35] and combines them into a decision policy with guarantees for selected and deferred patients.

We use StratCP with foundation models for retinal imaging [1], H&E whole-slide histopathology [2], and longitudinal EHRs [37], and evaluate it across diagnosis, biomarker prediction, operational and laboratory outcomes, and survival prognosis. StratCP controls the FDR among selected patients and returns calibrated prediction sets for deferred patients. When diagnostic guidelines define relationships among disease statuses, StratCP encodes these relationships in a utility graph and uses the graph to group related statuses in the prediction sets for deferred patients, such as tumor subtypes with the same or an adjacent WHO grade, while maintaining coverage.

In IDH mutation prediction, StratCP keeps the FDR within the 5% error budget, whereas `CP` exceeds it (FDR, 0.046 versus 0.096). When an IDH predictor trained on one cohort is applied to a new cohort, calibrating StratCP on as few as 50 labeled slides from the new cohort maintains error control among selected slides and coverage among deferred slides without retraining the predictor. We also evaluate StratCP in an independent Dana-Farber Cancer Institute (DFCI) cohort, where IDH status, molecular test results, and the integrated diagnosis, which combines histology with molecular findings such as IDH mutation and 1p/19q codeletion, were reported a median of 17.5, 20.5, and 32 days after tumor sample collection, respectively. StratCP selects predictions of IDH status and the integrated diagnosis from H&E slides within the error budget at the stage of histopathologic review, which precedes these results and could support earlier assessment during routine pathology workup. Together, these results show that StratCP provides decision policies with controlled error across data modalities, patient cohorts, and clinical workflows, and that these policies can be calibrated locally for any fixed FM predictor.

## Results

### Deciding when to act on foundation model predictions

StratCP operates on the predictions of a medical foundation model and decides, for each patient, whether to act on the prediction or defer the patient to further evaluation within an error budget set by the user (Fig. 1a–c). In the action arm, StratCP selects predictions that can directly guide care while controlling the false discovery rate (FDR), the expected fraction of incorrect predictions among selected patients, at a target level such as 5%. Patients whose predictions are not selected enter the deferral arm, where StratCP constructs a prediction set for each patient. To calibrate these sets, StratCP uses labeled calibration patients who would also be deferred under the same selection rule, so that among deferred patients the set contains the true disease status at the target coverage level, such as 95% (Methods). When clinical guidelines define relationships among disease statuses, an optional utility graph favors prediction sets that group clinically related statuses while maintaining coverage (Fig. 1a; Methods).

**Figure 1:**
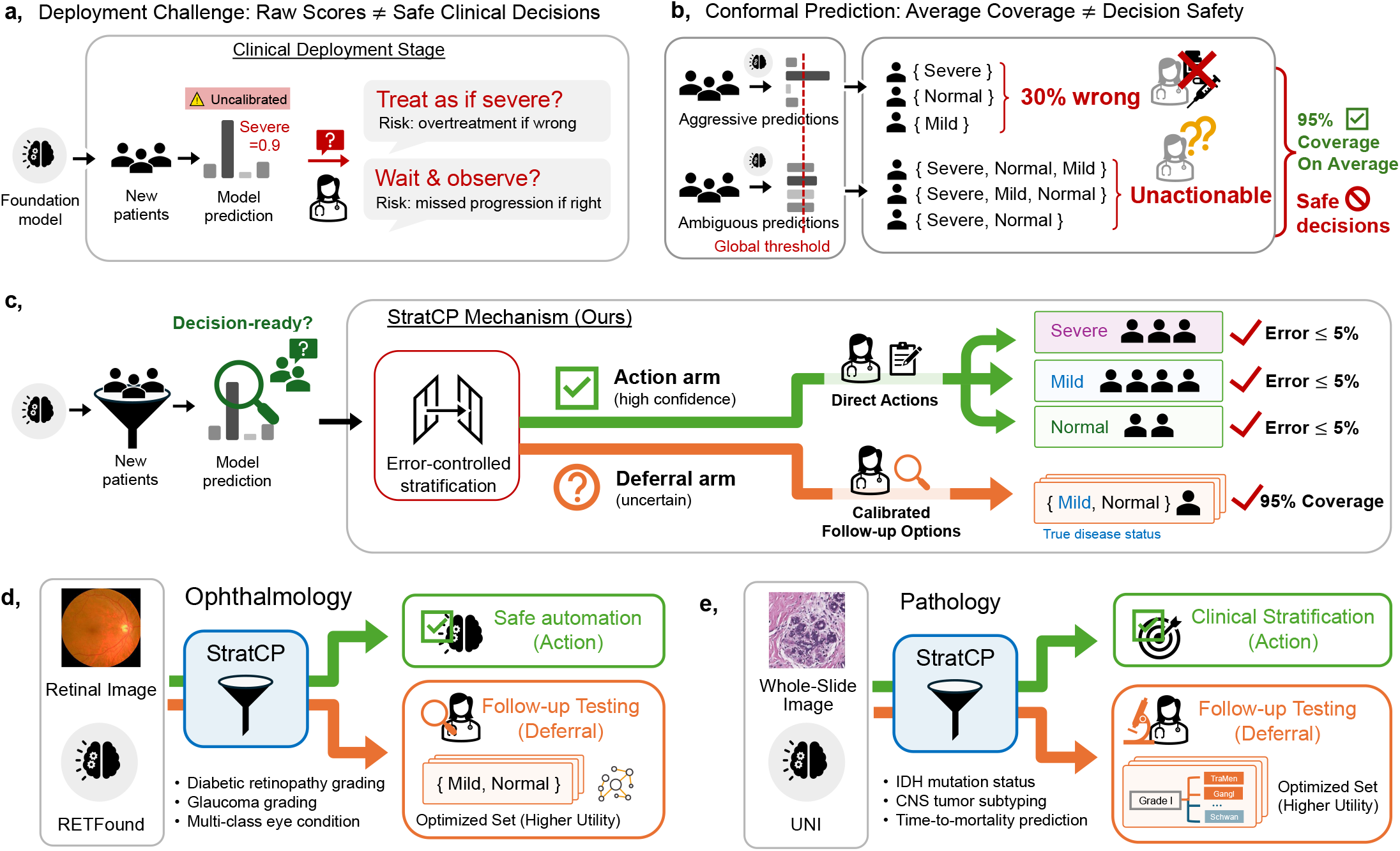
Overview of StratCP: from foundation model (FM) predictions to error-budgeted action and calibrated deferral. **a**, Raw FM scores do not by themselves support safe clinical decisions: a high predicted risk may prompt treatment escalation, whereas a lower score may prompt monitoring, and both choices can be harmful when miscalibrated. **b**, Standard conformal prediction provides average (marginal) coverage, but the error rate can vary across predicted disease statuses. Prediction sets containing multiple disease statuses may not be directly actionable when they combine diagnoses with different management pathways. **c**, StratCP performs error-controlled stratification: in the *action arm*, it selects predictions eligible for downstream action while controlling error among selected predictions within a user-specified budget (e.g., ≤ 5%) for each disease status; in the *deferral arm*, it returns prediction sets for deferred patients that achieve the target coverage (e.g., contain the true disease status for 95% of deferred patients), supporting calibrated follow-up. **d**, Ophthalmology: paired with RETFound [1] on retinal images, StratCP supports diabetic retinopathy staging, glaucoma staging, and multi-class eye-condition diagnosis by selecting patients in the action arm under the error budget and returning calibrated prediction sets for deferred patients; an optional utility module can reorder candidate disease statuses within each set to provide clinically related differentials (e.g., shared follow-up actions). **e**, Neuro-oncology: paired with UNI [2] on H&E whole-slide images, StratCP supports biomarker prediction (IDH mutation status), CNS tumor subtyping, and time-to-mortality prognosis by selecting predictions in the action arm under the error budget and deferring the remainder to calibrated prediction sets (or one-sided lower prediction bounds for prognosis). For CNS tumor subtyping, WHO grade-based relationships, such as shared grade, can be used to construct grade-coherent prediction sets.

In what follows, we evaluate StratCP across foundation models and clinical prediction tasks. We first apply StratCP to RETFound [1] for diabetic retinopathy grading, glaucoma staging, and the diagnosis of 39 eye conditions, and to UNI [2] for IDH mutation status and CNS tumor subtyping from H&E whole-slide images (Fig. 1d–e; Table 1). We then extend the evaluation to structured longitudinal EHRs with CLMBR-T-base [37] and to survival prediction with censored outcomes. Finally, we examine calibration of a fixed histopathology predictor after transfer to a new cohort, and a retrospective analysis of pathology workup in an independent clinical cohort.

**Table 1.**
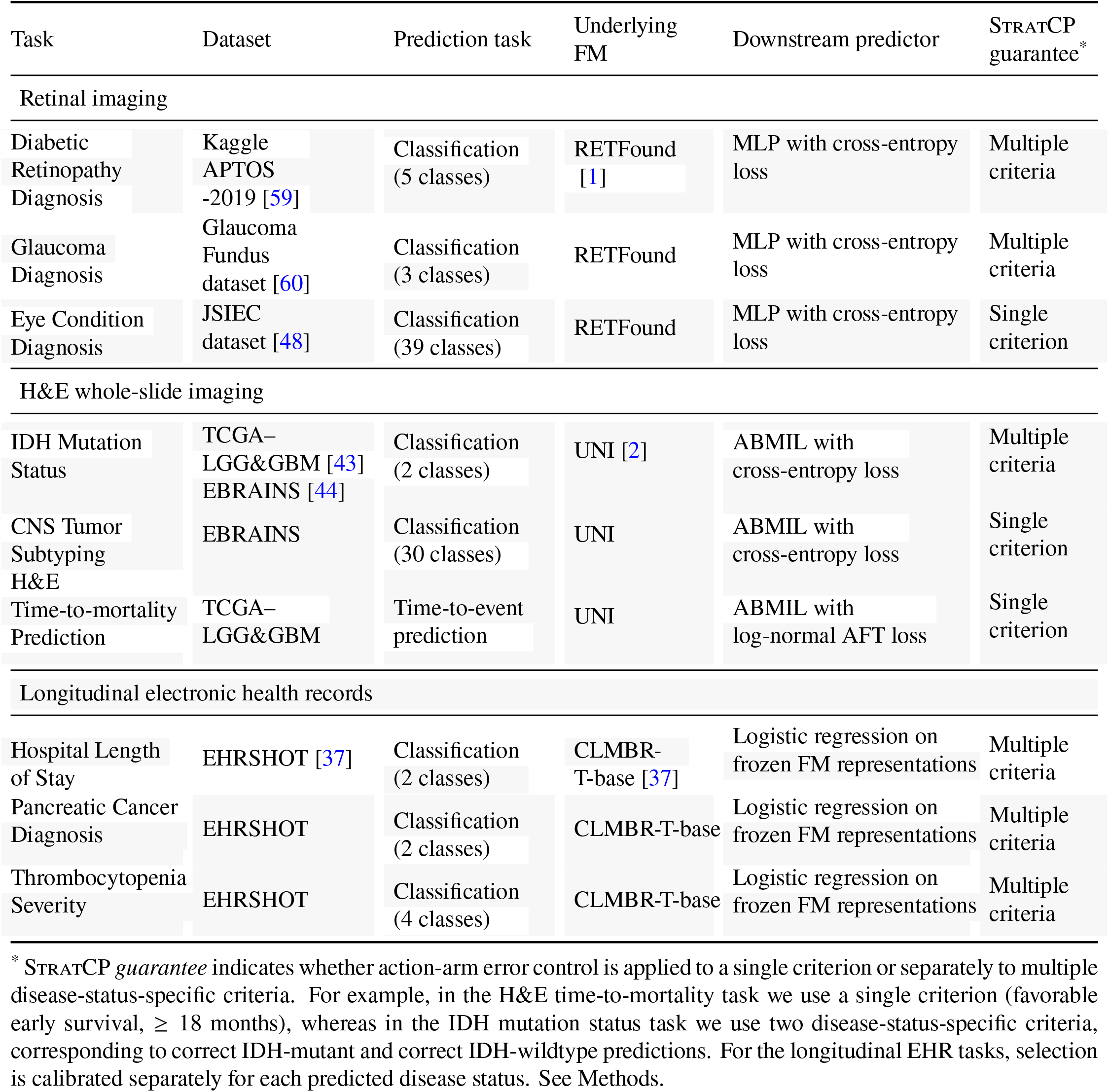
Prediction tasks, datasets, underlying foundation models (FMs), and downstream predictors used to evaluate StratCP across retinal imaging, H&E whole-slide imaging, and longitudinal electronic health records. In the *Downstream predictor* column, each FM provides representations that are used as inputs to the task-specific prediction model. “MLP” denotes a multilayer perceptron, and “ABMIL” denotes attention-based multiple instance learning [58], which aggregates patch-level embeddings for WSIs.

Throughout, we evaluate validity and efficiency. In the action arm, validity is quantified by FDR, the fraction of selected predictions that are incorrect on average (equivalently, selected-set coverage is one minus FDR). In the deferral arm, validity is quantified by coverage, the fraction of prediction sets that contain the true disease status [27]. Efficiency reflects how much useful information is provided at a given validity level: higher efficiency corresponds to selecting more patients under the same error budget and, among deferred patients, returning smaller prediction sets [38]. Our primary requirement is validity, meeting the pre-specified error or coverage target, before comparing efficiency among valid methods (Fig. 2c).

**Figure 2:**
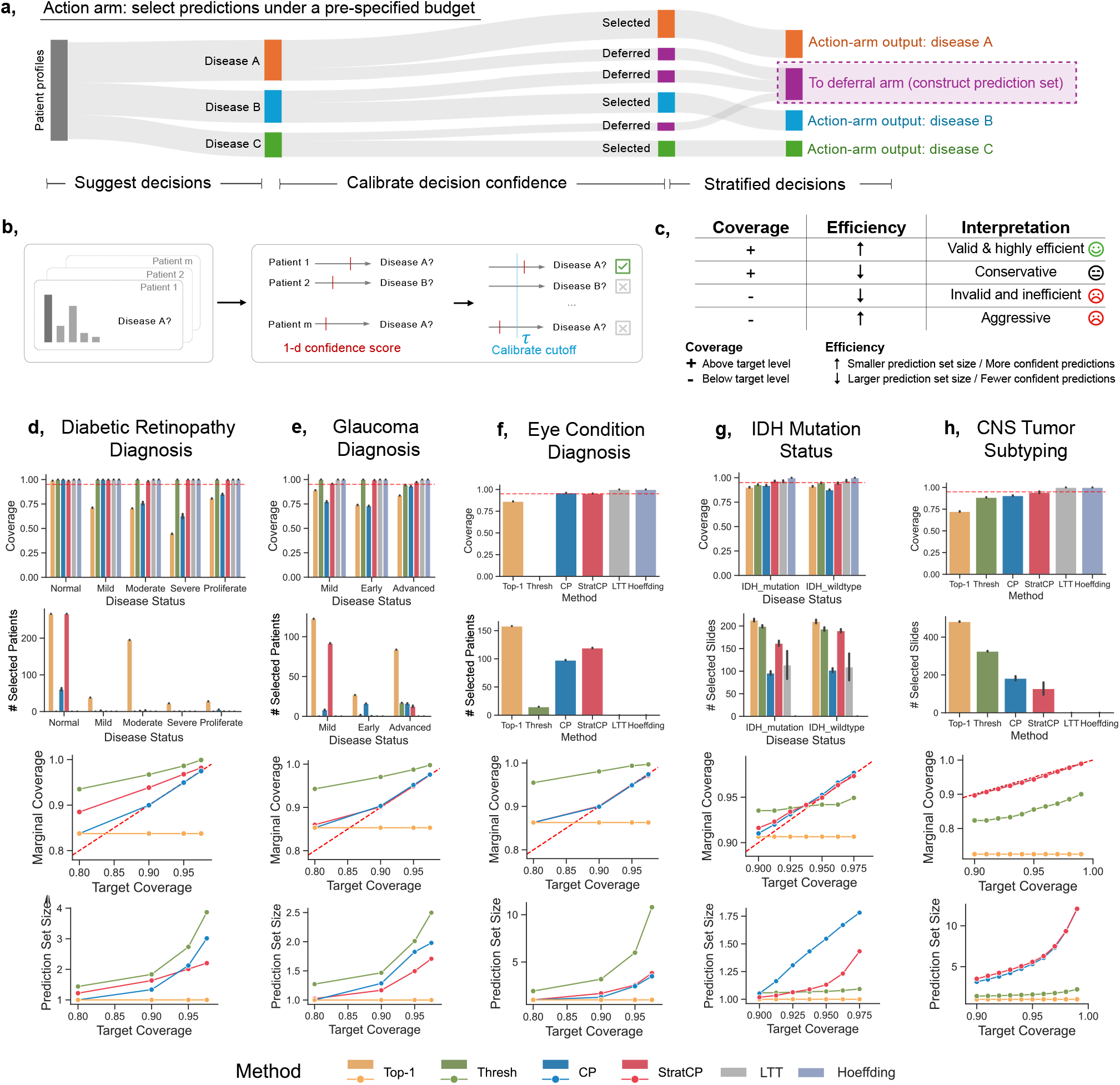
StratCP controls pre-specified error rates across ophthalmology and neuro-oncology tasks. **a**, In the action arm, StratCP maps FM patient predictions to disease statuses and selects patients eligible for action under a pre-specified error budget. Predictions not selected in the action arm are routed to the deferral arm, where StratCP returns prediction sets calibrated to achieve the target coverage (e.g., the set contains the true disease status for 95% of deferred patients). **b**, StratCP constructs a one-dimensional confidence score and calibrates a decision threshold using labeled calibration data, following a post-selection calibration procedure [35]. Predictions with scores above the threshold are selected in the action arm (Methods). **c**, Guide to interpreting performance in terms of coverage and efficiency. A method is *valid* if it achieves coverage at least the target level and *efficient* if it selects more patients or slides in the action arm or produces smaller prediction sets in the deferral arm. A method is *conservative* if its coverage exceeds the target level but efficiency is low. A method is *invalid and inefficient* if its coverage is below the target level and efficiency is low. A method is *aggressive* if its coverage is below the target level even though efficiency is high. **d**, Diabetic retinopathy severity grading. From top to bottom: (i) coverage (1 – false discovery rate) within each predicted stage among selected patients; (ii) the number of selected predictions per predicted stage; (iii) marginal coverage over all held-out patients across target levels; and (iv) marginal prediction-set size over all patients across target levels. See Methods for metric definitions. **e–h**, Same metrics as in **d** for glaucoma diagnosis (**e**), eye-condition diagnosis (**f**), IDH mutation status prediction (**g**), and CNS tumor subtyping (**h**). Results are averaged across 500 repeated calibration/test splits. Error bars indicate ±1 standard error of the mean across repeated data splits.

We compare StratCP with five existing approaches at an error budget of *α* = 0.05. `Top-1` reports the disease status with the highest predicted probability for every patient and never defers, which is the most common approach for medical FM predictions [1, 2, 4, 6, 7, 9, 39–41]. `Thresh` adds disease statuses in order of decreasing predicted probability until their cumulative probability exceeds 0.95, and selects a prediction for action only when the resulting set contains a single status. Conformal prediction (`CP`) constructs prediction sets with 95% marginal coverage and likewise selects predictions whose sets contain a single status. For the action arm, we also compare with learn then test (`LTT`) [42], which uses labeled calibration data to test candidate selection thresholds against the target error, and with `Hoeffding`, which selects predictions only when an upper confidence bound on the error among selected patients, derived from Hoeffding’s inequality, falls below the target (Methods).

### Selecting reliable retinal and histopathology predictions for action

Across the three retinal classification tasks, StratCP controlled action-arm error at the pre-specified 5% level, selecting patients for action only in disease statuses for which the calibration data supported the required error budget (Fig. 2d–f). All three tasks used fine-tuned RETFound models [1] applied to retinal images. In diabetic retinopathy grading, for example, StratCP selected a mean of 266 patients predicted as normal with an FDR of 0.011, corresponding to 98.9% coverage among selected predictions, while selecting no patients predicted as severe or proliferative, for which the underlying FM was less reliable (Fig. 2d; Supplementary Table S1 for patient counts and FM performance). In contrast, `Top-1` acted on all predictions and incurred FDRs of 0.555 and 0.194 for severe and proliferative disease, respectively. Results were consistent in glaucoma grading and 39-class eye-condition diagnosis (Fig. 2e–f; Supplementary Tables S2–S3). In glaucoma grading, StratCP selected predictions across all three disease stages while maintaining FDR at or below 0.047, whereas `Top-1` and `CP` exceeded the 5% error budget in multiple stages. In eye-condition diagnosis, both StratCP and `CP` remained within the 5% error budget, but StratCP selected more patients on average (119.2 versus 97.5; FDR, 0.048 versus 0.040), indicating greater selection efficiency among methods satisfying the error requirement. By comparison, `LTT` and `Hoeffding`-based selection met the action-arm error requirement but were more conservative across the retinal tasks, often selecting few or no predictions (Fig. 2d–f). Clinically, StratCP enables direct FM-supported disease-status assessment for patients whose predictions meet the required error budget, while avoiding definitive assignments when predictive reliability is insufficient.

We next evaluated action-arm error control and selection efficiency in H&E whole-slide histopathology. For IDH mutation status prediction, we evaluated whole-slide images from TCGA [43] and EBRAINS [44] using UNI [2] as the foundation model backbone (Table 1; Methods; Supplementary Table S4 for slide counts and per-class model performance). StratCP selected a mean of 162 IDH-mutant and 184 IDH-wildtype slides with FDRs of 0.046 and 0.047, respectively (Fig. 2g). `CP`, despite providing marginal coverage over all slides, exceeded the action-arm error budget for both disease statuses (FDR, 0.096 and 0.108). `LTT` and `Hoeffding` met the action-arm error requirement but selected substantially fewer slides than StratCP, showing a more conservative use of the same error budget (Fig. 2g). StratCP also produced smaller marginal prediction sets than `CP` across target levels (Fig. 2g, bottom panel), indicating more compact prediction sets overall.

In the more challenging 30-class CNS tumor subtyping task, StratCP likewise remained within the 5% error budget while retaining a non-empty action arm. Using UNI [2] on EBRAINS slides with WHO-defined subtypes [44, 45] (Table 1; Methods; Supplementary Table S7), StratCP selected a mean of 143 slides with an FDR of 0.047, whereas `CP`, `Thresh`, and `Top-1` exceeded the budget (FDR, 0.090, 0.119, and 0.275, respectively; Fig. 2h). `LTT` and `Hoeffding` satisfied the action-arm error requirement but selected no slides, whereas StratCP retained a non-empty action arm under the same target. Across target coverage levels, StratCP also maintained nominal marginal coverage over the full test population, with prediction sets comparable in size to `CP`. Therefore, StratCP can identify subsets of patients whose FM predictions can support disease-status assessment under a pre-specified error budget, while deferring cases that cannot be supported at the same error level.

### Clinically coherent prediction sets for deferred patients

In the deferral arm, StratCP returns prediction sets for patients or slides not selected for action, calibrated to achieve the target coverage level within the deferred population. Specifically, the prediction set contains the true disease status at the target rate (95% here; Fig. 3a; Methods), thereby providing calibrated differentials to guide follow-up. We compare StratCP with `Thresh` and conformal prediction (`CP`), evaluating each method only among patients it independently defers. `Top-1` is not applicable because it acts on every prediction and therefore has no deferral arm, while `LTT` and `Hoeffding` are selection-only baselines and do not return prediction sets for deferred patients. Across retinal tasks using RETFound [1], StratCP maintained coverage near the 95% target while deferring fewer patients than `CP` and `Thresh` (Fig. 3b–d). In diabetic retinopathy grading, for example, StratCP attained 95.0% coverage among 279.7 deferred patients, compared with 94.2% among 478 patients for `CP` and 98.6% among 550 patients for `Thresh` (Fig. 3b). The prediction sets returned by StratCP were correspondingly larger for these more difficult deferred cases (mean size, 2.98 versus 2.28 for `CP`), reflecting the uncertainty required to maintain coverage after selection. Consistent results were observed in glaucoma and 39-class eye-condition diagnosis, for which StratCP maintained coverage near the target while generally deferring fewer patients than the comparison methods (Fig. 3c–d).

**Figure 3:**
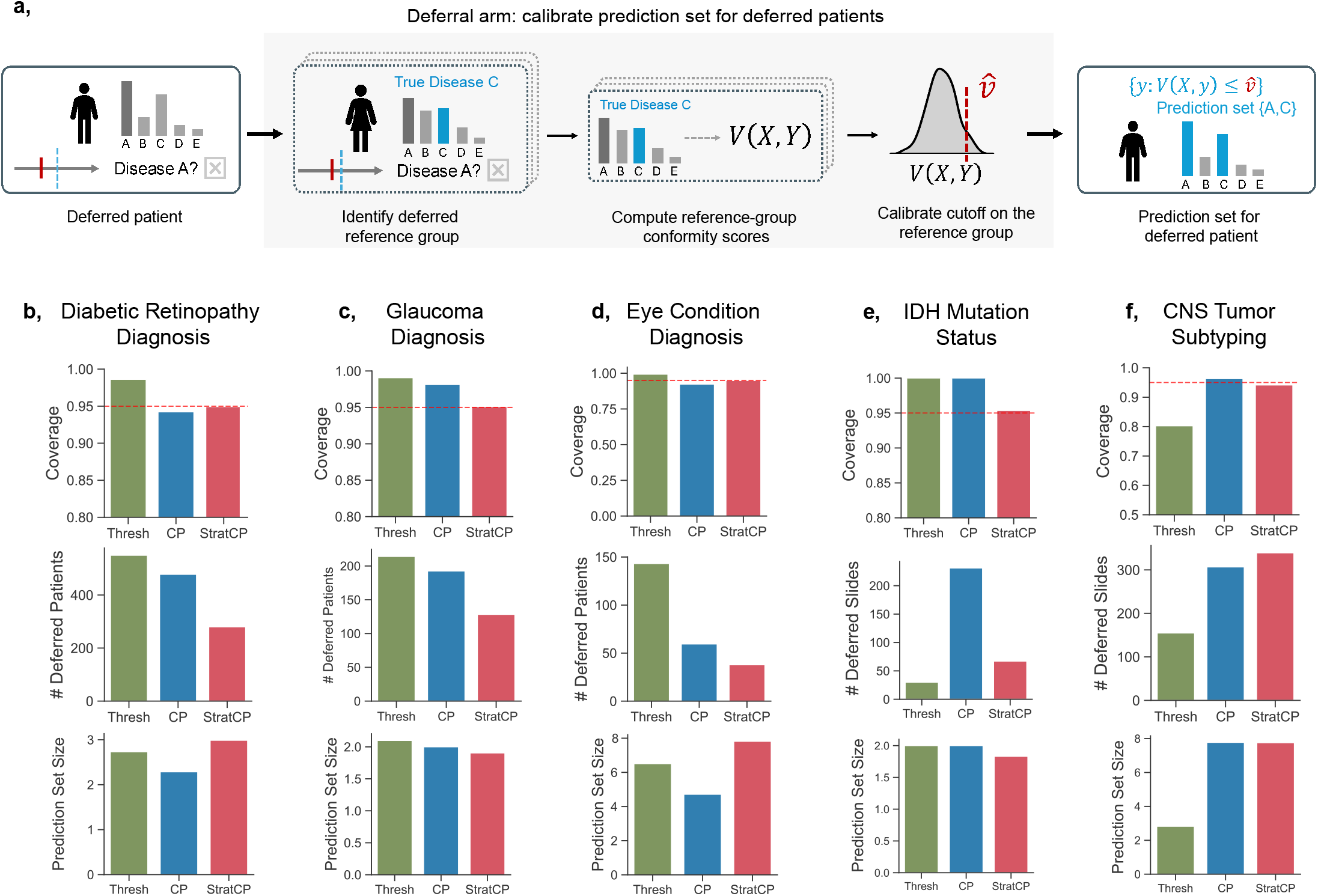
StratCP returns calibrated prediction sets for deferred patients. **a**, Deferral arm: for a new deferred patient or slide, StratCP calibrates a prediction set using labeled calibration samples that would also be deferred under the same action-arm selection rule; the resulting sets achieve the target coverage level within the deferred group (e.g., contain the true disease status for 95% of deferred patients or slides; Methods). **b**, Diabetic retinopathy: from top to bottom, deferred-set coverage (fraction of prediction sets containing the true disease status; target 95%), number of deferred patients, and mean prediction-set size at a target coverage of 95%. StratCP meets the 95% coverage target on deferred patients. **c–f**, Same metrics as in **b** for glaucoma diagnosis (**c**), eye-condition diagnosis (**d**), IDH mutation status prediction (**e**), and CNS tumor subtyping (**f**).

In neuro-oncology, StratCP paired with UNI [2] similarly maintained deferred-set coverage near the 95% target for IDH mutation status and CNS tumor subtyping (Fig. 3e–f). For IDH status, StratCP deferred 67 slides with a mean prediction-set size of 1.83, whereas `CP` deferred 231 slides with a mean set size of 2.00; `Thresh` deferred fewer slides but exceeded the error requirement in the action arm (Fig. 2g). For CNS tumor subtyping, StratCP and `CP` achieved similar deferred coverage (0.941 and 0.963) and prediction-set sizes (7.76 and 7.77), whereas `Thresh` under-covered (0.802; Fig. 3f). Thus, for patients not selected in the action arm, StratCP provides calibrated disease differentials that can guide confirmatory testing or expert review rather than forcing a definitive disease-status assignment.

Having established calibrated coverage in the deferral arm, we next asked whether these prediction sets could also be organized to reflect clinically meaningful relationships among candidate disease statuses. An optional utility module incorporates such structure through a utility graph, in which nodes represent disease statuses and edge weights encode how clinically appropriate two statuses are to include together, for example because they represent adjacent severity stages or share downstream management, while preserving the target coverage guarantee (Extended Data Fig. 1a; Methods). In diabetic retinopathy grading, an ordinal utility graph favored prediction sets containing consecutive severity stages while maintaining coverage (Extended Data Fig. 1b– d). In eye-condition diagnosis, a management-action utility graph based on shared downstream actions [46–48] increased management coherence at matched prediction-set sizes (Extended Data Fig. 1e–g; Supplementary Fig. S1; Methods). In CNS tumor subtyping, a WHO grade-based graph [45] increased the fraction of sets whose subtypes fell within the same or an adjacent grade: for set sizes 2–4, this fraction increased from 0.76 to 0.92, and for sizes 5–7, from 0.16 to 0.62, while coverage was preserved (Extended Data Fig. 1h–j). Therefore, StratCP not only preserves calibrated uncertainty after deferral, but can also structure that uncertainty into clinically coherent differentials that support more focused follow-up.

### Calibration in new cohorts and application to health records

We next examined whether StratCP supports FM predictors that are transferred to a new patient cohort or used with a different clinical data type, first for a fixed histopathology predictor transferred to an independent cohort and then for predictors built from longitudinal EHR data.

We trained an IDH predictor built on UNI [2] with TCGA slides and applied it to the independent EBRAINS cohort. The predictor reached an AUROC of 0.951 in held-out TCGA slides and 0.941 in EBRAINS, and slide embeddings differed between the two cohorts (Fig. 4a–b; Supplementary Tables S5–S6). We then calibrated StratCP on labeled EBRAINS slides and evaluated it on held-out EBRAINS slides, using calibration sets of 50 to 400 slides (Methods). At *α* = 0.05, StratCP kept the FDR among selected slides below 0.05 for every calibration set size. With only 50 labeled EBRAINS slides, it selected a mean of 156 held-out slides (Fig. 4c–d). In contrast, `CP` exceeded the error budget for IDH-mutant and IDH-wildtype predictions, whereas `LTT` and `Hoeffding` were not able to select any slides for action at any calibration set size. Among deferred slides, StratCP achieved coverage close to the 0.95 target for every calibration set size (0.949 to 0.952; Fig. 4e). Clinically, a small labeled sample from a new cohort can identify which IDH predictions from H&E slides can be acted on within the error budget and which should be deferred to confirmatory testing.

**Figure 4:**
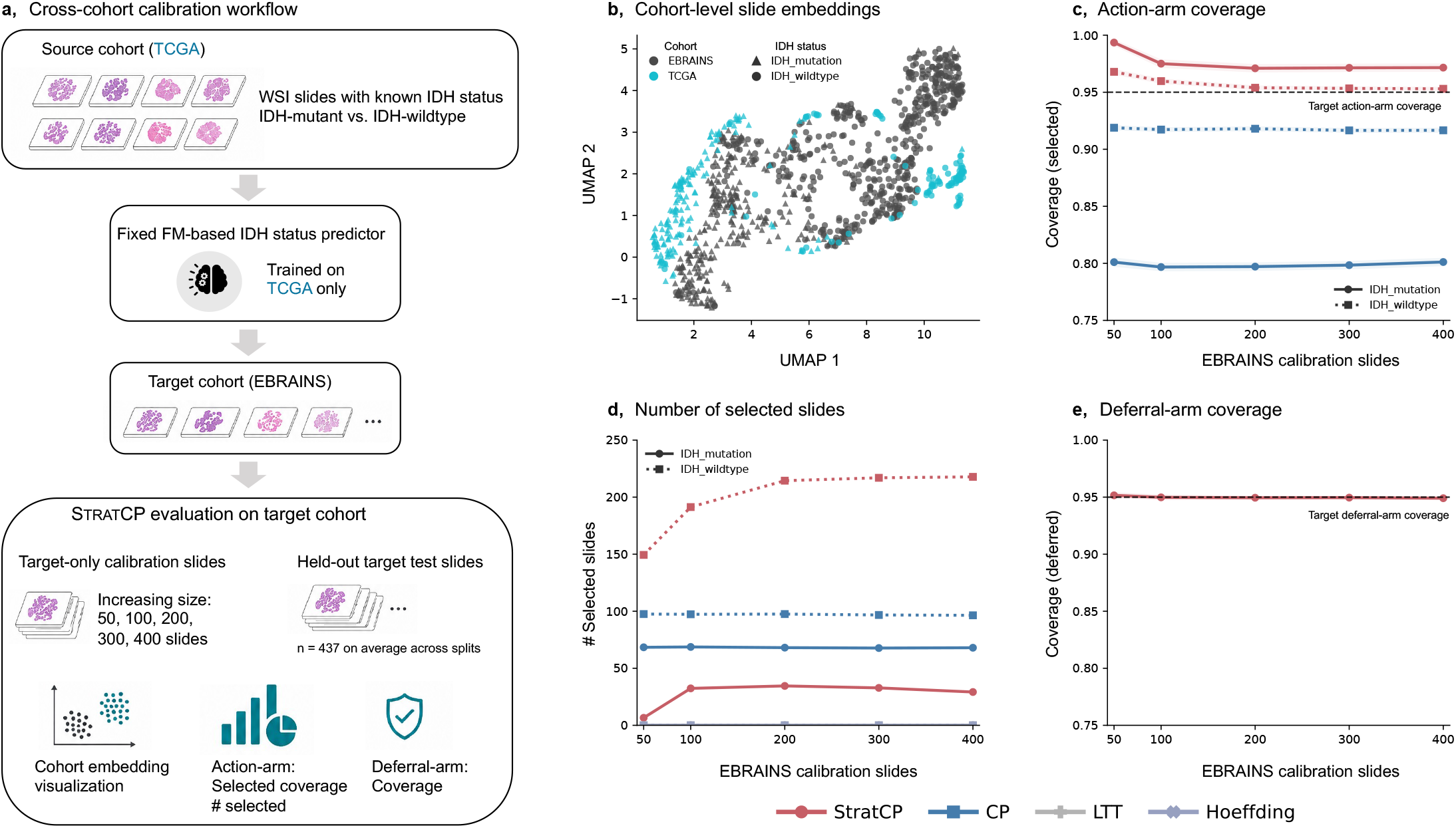
StratCP maintains coverage for selected and deferred slides after cross-cohort transfer in IDH mutation status prediction. **a**, Schematic of the cross-cohort evaluation design. A fixed UNI-based ABMIL slide-level IDH predictor trained on TCGA was applied to EBRAINS without retraining, with StratCP calibrated using labeled EBRAINS slides only. Target-only calibration sets of 50, 100, 200, 300, or 400 EBRAINS slides were used to evaluate action-arm selection and deferral-arm coverage as target-cohort calibration-set size increased. **b**, UMAP visualization of patient-level representations from TCGA and EBRAINS, obtained by averaging slide embeddings within each patient. Each point represents one patient, with color indicating cohort and shape indicating IDH status. **c**, Coverage for selected slides, defined as 1 − FDR, for IDH-mutant and IDH-wildtype predictions across EBRAINS calibration-set sizes. The dashed horizontal line denotes the nominal 95% target coverage. **d**, Number of selected slides across EBRAINS calibration-set sizes. **e**, Coverage among deferred slides across calibration-set sizes; the dashed horizontal line denotes the nominal 95% target coverage.

We next used StratCP with CLMBR-T-base [37], a foundation model for longitudinal EHR records. We considered three tasks: hospital length of stay (< 7 versus ≥ 7 days), new pancreatic-cancer diagnosis (no cancer versus cancer), and thrombocytopenia severity prediction (normal, mild, moderate, or severe) (Extended Data Fig. 4a; Supplementary Table S8–S9). At *α* = 0.05, StratCP controlled action-arm error at or below the pre-specified target in each prediction class in which it selected patients and abstained when the calibration data did not support error-controlled action (Extended Data Fig. 4b–c). For example, it selected 75.5% of predictions for shorter hospital stay with an FDR of 0.047 and 42.3% of predictions for normal platelet status with an FDR of 0.049, while abstaining from predictions of pancreatic cancer and the less reliably predicted thrombocytopenia severity levels. By comparison, `CP, Top-1`, and `Thresh` exceeded the action-arm error budget in several prediction classes, whereas `LTT` and `Hoeffding` selected no patients across the three tasks.

Among deferred patients, StratCP achieved 0.950 coverage across all three tasks (Extended Data Fig. 4d). For ordinal thrombocytopenia severity, utility enhancement further favored prediction sets containing adjacent severity levels while preserving coverage (Extended Data Fig. 4e).

Together, these results show that StratCP can translate fixed FM-based predictions into error-controlled action and calibrated deferral under cohort shift and in EHR data, supporting action when the pre-specified error budget can be met while directing remaining patients to further clinical evaluation.

### Error-controlled survival prediction in diffuse glioma

We next asked whether StratCP could extend from disease-status classification to time-to-event prediction. Adult-type diffuse gliomas are associated with substantial morbidity and mortality, making early prognostication relevant to patient counseling and clinical management [49–51]. Using UNI [2] representations from H&E whole-slide images, we trained an ABMIL-based lognormal accelerated failure time (AFT) survival model using a right censoring likelihood (Methods; Supplementary Table S10). We defined favorable early survival as survival for ≥ 18 months, a clinically relevant horizon that exceeds the typical median overall survival of glioblastoma under contemporary standard-of-care therapy [52]. StratCP then selects patients predicted to achieve favorable early survival while controlling action-arm error and, for deferred patients, returns a one-sided lower prediction bound (LPB) defining the calibrated survival interval [LPB, ∞) (Fig. 5a–b). The arms account for right censoring using inverse probability of censoring weighting.

**Figure 5:**
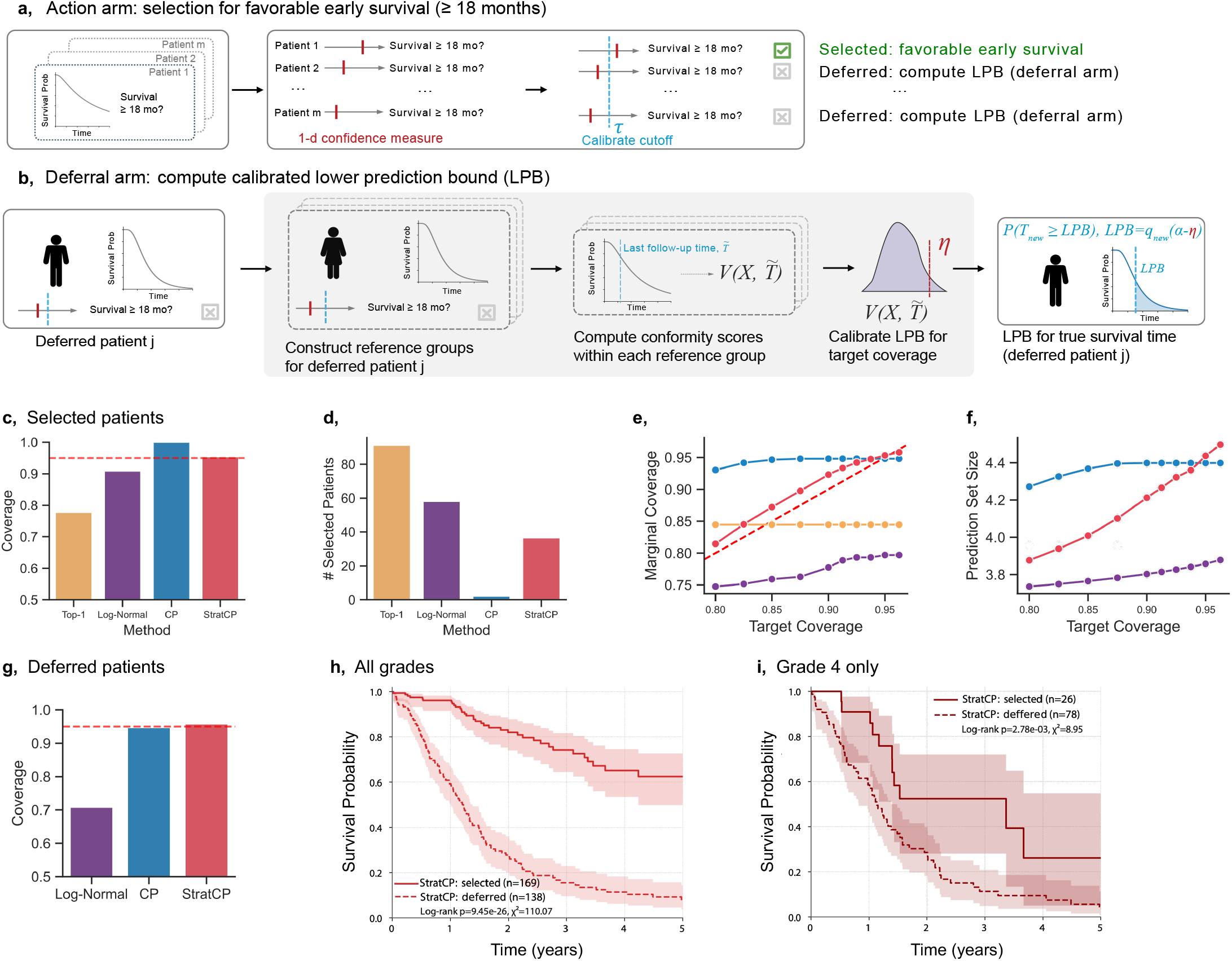
StratCP identifies patients with favorable early survival ( ≥ 18 months) from H&E whole-slide images and returns calibrated lower bounds for deferred patients. **a**, Action arm. The model maps each patient to a one-dimensional confidence score for survival beyond 18 months and selects patients using a calibrated cutoff to control the false discovery rate (FDR) at 5% (Methods). Selected patients receive a favorable early-survival prediction for downstream assessment; remaining patients are deferred. **b**, Deferral arm. For a deferred patient *j*, StratCP constructs patient-specific reference groups, calibrates conformity-score thresholds, and reports a one-sided lower prediction interval [LPB, ∞) (Methods). **c**, Censoring-adjusted coverage among selected patients for favorable early survival ( ≥ 18 months); the dashed horizontal line denotes the 95% target. **d**, Number of selected patients at the 5% error budget. **e**, Marginal coverage across all test patients over target levels, where selected patients receive the set [18 months,∞) and deferred patients receive [LPB, ∞). **f**, Prediction-set size across target coverage levels, measured as the mean interval length (in years) from the LPB to the fixed study end time (5 years); smaller values indicate tighter lower bounds. **g**, Censoring-adjusted coverage among deferred patients, defined by whether survival is at least the calibrated LPB, at a target coverage of 95%. **h–i**, Survival curves. Kaplan–Meier estimates comparing selected versus deferred patients across splits, with shaded regions denoting 95% confidence intervals: **h**, all grades; **i**, Grade 4. Significance is assessed by log-rank test. *Baselines*. `Top-1` (select if estimated survival at 18 months ≥ 0.5), `LNQ` (log-normal quantile; parametric selector and bound), and `CP` (selects when the conformal lower bound is ≥ 18 months). Right censoring is handled with inverse probability of censoring weighting (IPCW; Methods).

In the action arm, at *α* = 0.05, StratCP selected patients with an FDR of 0.046, whereas `Top-1` and the parametric log-normal quantile baseline (`LNQ`) exceeded the FDR target (0.223 and 0.092, respectively; Fig. 5c). StratCP selected substantially more patients than conformal prediction (`CP`) while maintaining the FDR requirement (36.6 versus 2.1 patients on average; Fig. 5d). StratCP and `CP` also maintained nominal marginal coverage over the full cohort across error target levels, whereas `LNQ` did not (Fig. 5e). Among deferred patients, StratCP achieved coverage of 0.958, comparable to `CP` at 0.947 and substantially higher than `LNQ` at 0.708, while returning prediction sets of similar size to `CP` (Fig. 5f–g).

Patients selected by StratCP also exhibited significantly longer observed survival than deferred patients (log-rank *p* = 9.45 × 10^−26^; Fig. 5h). This separation remained significant among Grade 4 gliomas (log-rank *p* = 2.78 × 10^−3^; Fig. 5i). StratCP selects patients whose predicted survival of at least 18 months can guide prognostic assessment within the error budget, and gives deferred patients a calibrated lower bound on survival time that can inform counseling and further care. These results also demonstrate StratCP for the complex task of survival prediction with censored outcomes.

### Glioma diagnosis from H&E slides before molecular testing

High-grade adult-type diffuse gliomas are aggressive tumors for which integrated disease classification defines clinically distinct disease states with implications for prognosis and treatment planning. Under the WHO guideline, classification of these tumors requires integration of histomorphology with defining molecular features, including IDH mutation and 1p/19q codeletion [45, 50]. Integrated diagnosis involves a sequential evaluation of H&E histopathology, immunohistochemical findings, and molecular testing (Fig. 6a).

**Figure 6:**
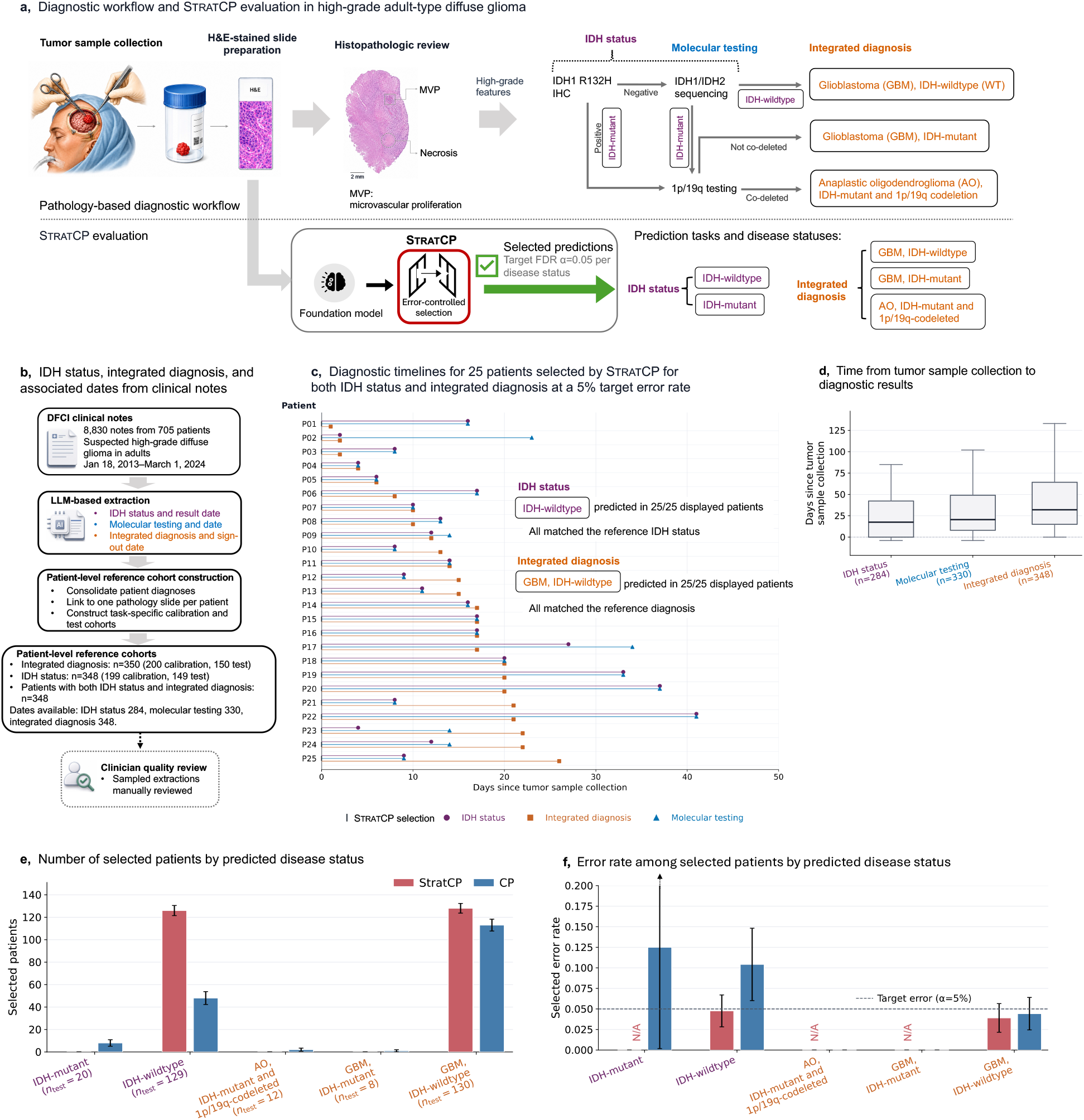
StratCP enables error-controlled disease-status prediction during the pathology work-up of high-grade adult-type diffuse glioma. **a**, Schematic of the pathology-based diagnostic workflow, from tumor sample collection and H&E-stained slide preparation through histopathologic review, IDH status determination, molecular testing, and integrated diagnosis. For retrospective evaluation, StratCP selected predictions for IDH status and integrated diagnosis at a target error rate of *α* = 0.05, with the selection time referenced to tumor sample collection. **b**, LLM-based extraction of IDH status, molecular testing, integrated diagnosis, and associated dates from 8,830 clinical notes representing 705 patients. After patient-level consolidation and slide linkage (Methods), the integrated-diagnosis and IDH analysis cohorts comprised 350 and 348 patients, respectively, with 348 patients shared between tasks. Among these 348 patients, dated IDH IHC and molecular results were available for 284 and 330 patients, respectively. Sampled extractions underwent manual review by a board-certified clinician. **c**, Diagnostic timelines for 25 patients randomly sampled from those selected by StratCP for both tasks. Day 0 denotes tumor sample collection, to which the StratCP selection time was referenced for this retrospective analysis. Displayed patients had IDH-status, molecular-testing, and integrated-diagnosis intervals between 0 and 60 days and were ordered by time to integrated diagnosis.**d**, Time from tumor sample collection to IDH-status, molecular-testing, and integrated-diagnosis results. Boxes show medians and interquartile ranges; whiskers denote the 10th–90th percentiles.**e**, Number of patients selected by StratCP or CP in each predicted disease-status category. *n*_test_ denotes the number of held-out test patients with the corresponding reference disease status.**f**, Error rate among selected patients by predicted disease status. Error bars denote ±1 bootstrap standard error estimated from 2,000 patient-level resamples; N/A indicates no selected patients.

In a complementary analysis of TCGA and EBRAINS slides, StratCP supported morphology-stratified H&E-based disease-status prediction under a 5% error budget (Extended Data Fig. 3; Supplementary Table S11). We therefore retrospectively evaluated whether StratCP could select H&E-based predictions of IDH status and integrated diagnosis in an independent Dana-Farber Cancer Institute (DFCI) cohort using FM-based predictors fine-tuned on TCGA and EBRAINS (Supplementary Table S12 for DFCI cohort composition and predictive performance), and characterized the time from tumor sample collection to routine diagnostic results to assess the potential window for earlier clinical assessment.

We reconstructed diagnostic timelines from 8,830 clinical notes representing 705 adults evaluated for high-grade diffuse glioma (Fig. 6b). An LLM-based extraction pipeline was used to extract IDH status, molecular testing, integrated diagnosis, and associated dates (Methods). Manual clinician review of sampled extractions showed 100% agreement for IDH status and 93.3% agreement for integrated diagnosis, with a median date difference of 0 days across all evaluated milestones (Methods; Extended Data Fig. 2; Supplementary Table S13). Patient-level consolidation and slide linkage yielded 350 patients for integrated-diagnosis evaluation and 348 patients for IDH-status evaluation, with the latter 348 patients also forming the paired cohort (Methods). Dated integrated diagnosis, IDH status, and molecular results were available for 348, 284, and 330 patients, respectively. StratCP was calibrated and evaluated using a temporal split by tumor collection date (Methods). Representative patient timelines illustrate the sequence from tumor sample collection through IDH, molecular, and integrated diagnostic results (Fig. 6c). Across patients with available dates, the median times from sample collection were 17.5 days to IDH status, 20.5 days to molecular results, and 32.0 days to integrated diagnosis (Fig. 6d). Because H&E-based predictions can be generated once slides become available, these intervals indicate a potential window for earlier model-supported assessment during routine work-up.

In the action arm, at *α* = 0.05, because the DFCI evaluation used a single pre-specified temporal split, we report the selected error rate rather than an empirical FDR averaged across repeated splits (Methods). StratCP selected 126 of 149 test patients as IDH-wildtype, with a selected error rate of 0.048, whereas `CP` selected 48 patients with an error rate of 0.104 (Fig. 6e–f). For integrated diagnosis, StratCP selected 128 of 150 test patients as glioblastoma, IDH-wildtype, with a selected error rate of 0.039, compared with 113 patients and an error rate of 0.044 for `CP`. Patient-level bootstrap standard errors are shown in Fig. 6f. StratCP abstained from selection across both tasks when the calibration data did not support the pre-specified error level. StratCP therefore identified patient subsets for whom H&E-based IDH-status and integrated-diagnosis predictions could support earlier assessment with selected error rates consistent with the prespecified 5% error budget, while abstaining otherwise.

## Discussion

Using foundation models to guide care raises a question beyond the average accuracy of model predictions, namely when a foundation model prediction can support clinical action and when clinicians should defer the decision to confirmatory testing or expert review. StratCP addresses this question by selecting patients whose predictions can guide care with controlled error and by returning calibrated prediction sets for the remaining patients. In the action arm, StratCP selects patients whose predictions can guide care while controlling the false discovery rate within an error budget set by the user. For other patients, the deferral arm returns prediction sets that contain the true disease status at the target rate. Across diagnosis, biomarker prediction, survival prognosis, and EHR outcomes, StratCP produces decision policies that specify when to act on a prediction and which alternative disease statuses to consider when deferring. Our results also show that StratCP can be calibrated with labeled data from a new cohort without retraining the FM predictor.

StratCP extends conformal prediction from guarantees that hold on average over all patients (marginal coverage) to guarantees that hold for the patients affected by each clinical decision. Marginal coverage of conformal prediction does not ensure error control among the subset of patients whose FM predictions are selected for action, and in several tasks standard conformal prediction exceeded the error budget for specific disease statuses. StratCP gives FMs that other-wise return only point predictions a principled way to defer uncertain cases to confirmatory testing or expert review [53]. Utility graphs can further organize the prediction sets of deferred patients according to clinical relationships among disease statuses while maintaining coverage.

StratCP works with any foundation model, because it operates on predictions from an FM predictor without retraining the underlying model. This feature allows StratCP to be used across clinical sites, adapted as data and workflows change, and integrated at decision points in clinical care. Consistent with this flexibility, when a fixed IDH predictor was transferred from TCGA to EBRAINS, calibration on as few as 50 labeled EBRAINS slides maintained error control among selected slides and calibrated prediction sets for deferred slides within the error budget. The EHR analyses further extended StratCP beyond imaging to structured longitudinal clinical data. In the retrospective DFCI cohort, StratCP selected large groups of patients for earlier assessment of IDH status and integrated diagnosis from H&E slides during routine pathology workup.

Our study has limitations. The guarantees of StratCP rely on exchangeability between patients in the calibration set and patients in the target set to whom StratCP is applied, and distribution shifts across sites or technical factors, such as staining protocols or scanners, can weaken error control and coverage. Although calibration on the target cohort maintained error control and coverage in the TCGA to EBRAINS analysis, this evaluation does not establish robustness to arbitrary distribution shifts. FMs alone do not resolve covariate or label shift [54], so local calibration and ongoing performance monitoring are important when patient populations or workflows change [55]. Performance also depends on label quality and on having sufficient calibration examples for rare disease statuses. When labels are noisy or calibration data are sparse, prediction sets can widen and the action arm of StratCP may select few or no patients for some statuses. In survival prediction, error control among selected patients and coverage among deferred patients rely on inverse probability of censoring weighting and were evaluated empirically, because the finite-sample guarantees do not extend directly to right-censored outcomes. The utility graphs of StratCP encode expert knowledge of relationships among disease statuses, but this knowledge may vary across institutions and change over time, so the graphs need regular review to stay aligned with current clinical practice. StratCP also inherits the predictive limits of the underlying foundation model. Weaker models lead to larger prediction sets and more deferral, whereas stronger models support action for more patients under the same error budget. Finally, the DFCI evaluation was retrospective, used a single temporal split, and selected predictions only for IDH-wildtype tumors, which formed the majority of this cohort. Prospective studies are needed to assess how H&E assessment guided by StratCP affects diagnostic turnaround, confirmatory testing, clinical decisions, and patient outcomes.

Future work could extend decision policies with controlled error to additional clinical settings and endpoints. Multisite studies could evaluate how well StratCP transfers across patient populations and workflows, including how often local recalibration is needed as conditions change.

Another direction is online error control, in which the FDR among selected patients is controlled as new patients arrive one at a time, instead of for a fixed test cohort. In cancer care, future extensions could include biomarker prediction and diagnostic assessment in settings with limited access to confirmatory testing, as well as prediction of treatment response, disease progression, and adverse events [11, 56, 57]. StratCP provides a general approach for acting on foundation model predictions with controlled error and for guiding follow-up with calibrated prediction sets when predictions are not reliable enough for action.

## Acknowledgements

We thank David M. Meredith, MD, PhD, for feedback on the neuropathology work-up analysis. We gratefully acknowledge the support of NIH R01-HD108794, NSF CAREER 2339524, Harvard Data Science Initiative, Amazon Faculty Research, Google Research Scholar Program, AstraZeneca Research, Roche Alliance with Distinguished Scientists (ROADS) Program, Sanofi iDEA-iTECH Award, GlaxoSmithKline Award, Boehringer Ingelheim Award, Merck Award, Optum AI Research Collaboration Award, Pfizer Research, Gates Foundation (INV-079038), Chan Zuckerberg Initiative, John and Virginia Kaneb Fellowship at Harvard Medical School, Biswas Computational Biology Initiative in partnership with the Milken Institute, Harvard Medical School Dean’s Innovation Fund for the Use of Artificial Intelligence, and the Kempner Institute for the Study of Natural and Artificial Intelligence at Harvard University. This work was delivered as part of the Aurora project supported by the Cancer Grand Challenges partnership funded by Cancer Research UK (CGCAI1-Mar26/100001). Any opinions, findings, conclusions or recommendations expressed in this material are those of the authors and do not necessarily reflect the views of the funders.

## Data availability

Processed data used in the paper, including model predictions and true labels for the held-out (test + calibration) set for the retinal diagnosis tasks, are available via the project website at https://zitniklab.hms.harvard.edu/projects/StratCP. The raw datasets for the retinal diagnosis tasks are publicly available at the following sources: diabetic retinopathy diagnosis (https://www.kaggle.com/competitions/aptos2019-blindness-detection/data), glaucoma diagnosis (https://dataverse.harvard.edu/dataset.xhtml?persistentId=doi:10.7910/DVN/1YRRAC), and eye condition diagnosis (https://zenodo.org/records/3477553). For neuro-oncology tasks, whole-slide images (WSIs) were obtained from TCGA-LGG and TCGA-GBM (publicly available via the NIH GDC portal: https://portal.gdc.cancer.gov/ [43]) and from the EBRAINS repository (available upon approved request: https://search.kg.ebrains.eu/ [44]). For the longitudinal EHR analyses, the EHRSHOT dataset [37] is available through Stanford Redivis under credentialed access (https://stanford.redivis.com/datasets/53gc-8rhx41kgt), subject to the applicable EHRSHOT data-use and access requirements. The retrospective DFCI/BWH clinical and pathology data used for the independent neuro-oncology evaluation were obtained from institutional resources under IRB approval and are not publicly available because of patient-privacy and institutional data-governance restrictions. Access would require applicable institutional approvals and data-use agreements.

## Code availability

The Python implementation of StratCP is available at the project website: https://zitniklab.hms.harvard.edu/projects/StratCP. The code to reproduce the reported results, along with usage examples, is provided at https://github.com/mims-harvard/StratCP. For ophthalmology tasks, the fine-tuned RETFound model checkpoints for each diagnosis task and data split are available at https://github.com/rmaphoh/RETFound. For neuro-oncology tasks, we used the UNI foundation model available on HuggingFace (https://huggingface.co/MahmoodLab/UNI). Fine-tuned ABMIL model checkpoints for these tasks are also available via the project GitHub page. For the longitudinal EHR tasks, we used the CLMBR-T-base foundation model [37], which is available through the Stanford Shah Lab Hugging Face repository (https://huggingface.co/StanfordShahLab/clmbr-t-base). Access to the model weights is gated and requires acceptance of the EHRSHOT Credentialed Health Data License and completion of the associated credentialing requirements.

## Author contributions

I.M. processed neuro-oncology H&E whole-slide images (WSIs), conducted analyses and benchmarking across the neuro-oncology tasks, and designed and implemented the time-to-event regression methodology. Y.J. retrieved, processed, and analyzed retinal disease diagnosis data and models. Y.J. designed statistical methods, implemented the general framework, and benchmarked StratCP on retinal disease diagnosis tasks. M.S. advised on the DFCI/BWH neuropathology work-up analysis and reviewed patient notes and LLM-based extractions. A.G. advised on the neuropathology work-up analysis and the use of the in-house cohort. I.M., Y.J., and M.Z. conceptualized and designed the study. All authors contributed to writing and revising the manuscript.

## Competing interests

None.

**Extended Data Fig. 1:**
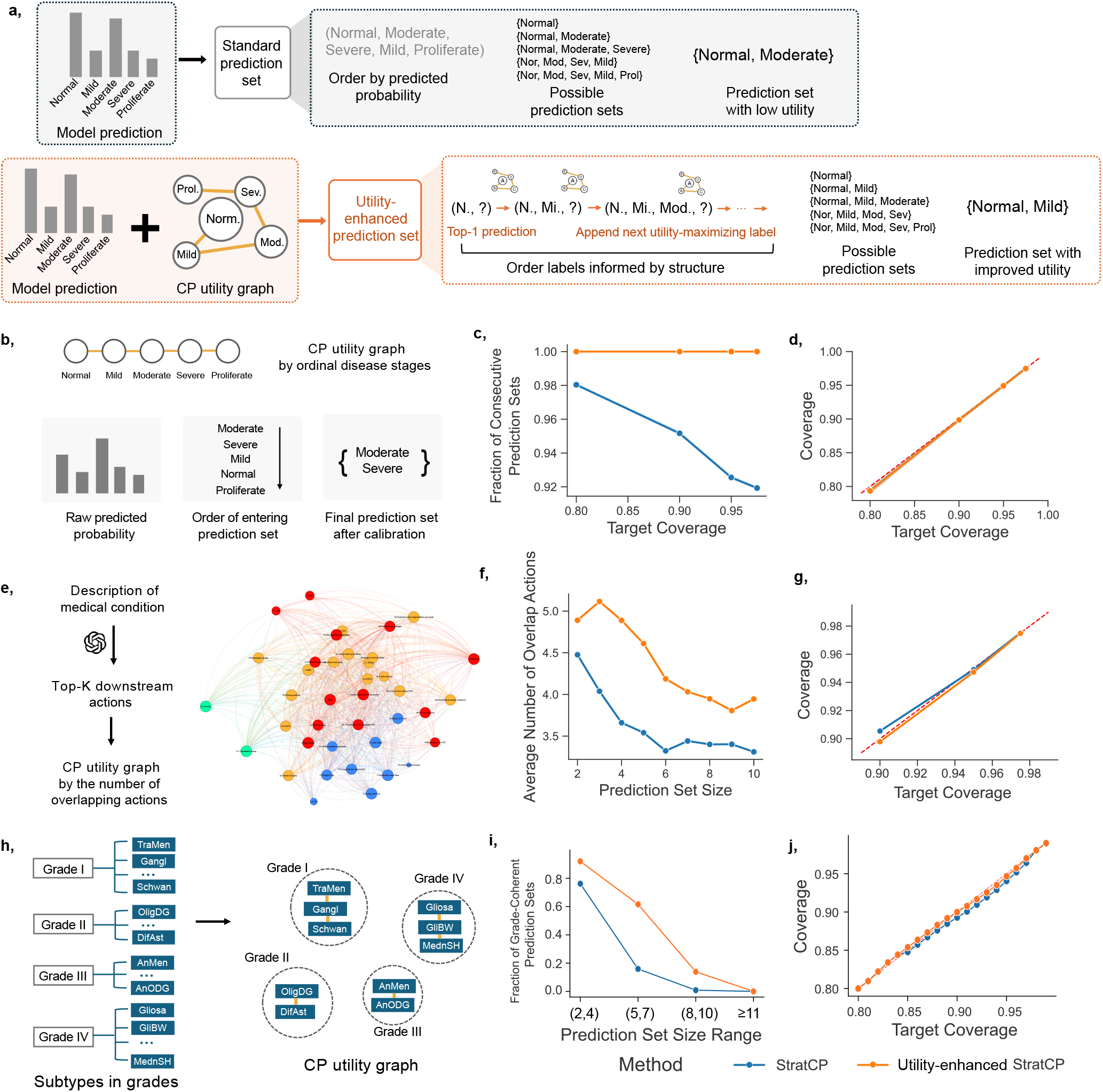
StratCP improves the clinical coherence of deferred prediction sets using utility graphs. **a**, Utility enhancement module. Given a utility graph encoding clinically meaningful relationships among disease statuses, StratCP constructs a utility-informed ordering of candidate disease statuses. Starting from the disease status with the highest FM-predicted probability, StratCP iteratively adds disease statuses that maximize utility relative to those already selected; the calibrated prefix of this ordering forms the deferred prediction set (Methods). **b**, Diabetic retinopathy grading uses an ordinal utility graph favoring adjacent severity stages. **c**, Fraction of prediction sets containing consecutive stages across target levels. **d**, Empirical coverage after utility enhancement. **e**, Eye-condition classification uses a management-action utility graph, with edge weights reflecting overlap in LLM-derived downstream management actions between disease-status pairs (Methods). **f**, Mean number of overlapping management actions within deferred prediction sets at matched set size. **g**, Empirical coverage after utility enhancement. **h**, CNS tumor subtyping uses a WHO grade-based utility graph connecting subtypes assigned to the same WHO grade. Subtypes from different grades are not connected (Methods). **i**, Fraction of deferred prediction sets containing the top-1 predicted subtype and satisfying the same-or-adjacent-grade coherence criterion, defined as a maximum within-set grade difference < 2, across prediction-set-size bins. **j**, Empirical coverage after utility enhancement. Blue denotes standard StratCP and orange utility-enhanced StratCP. Dashed diagonals in **d, g**, and **j** indicate equality between empirical and target coverage.

**Extended Data Fig. 2:**
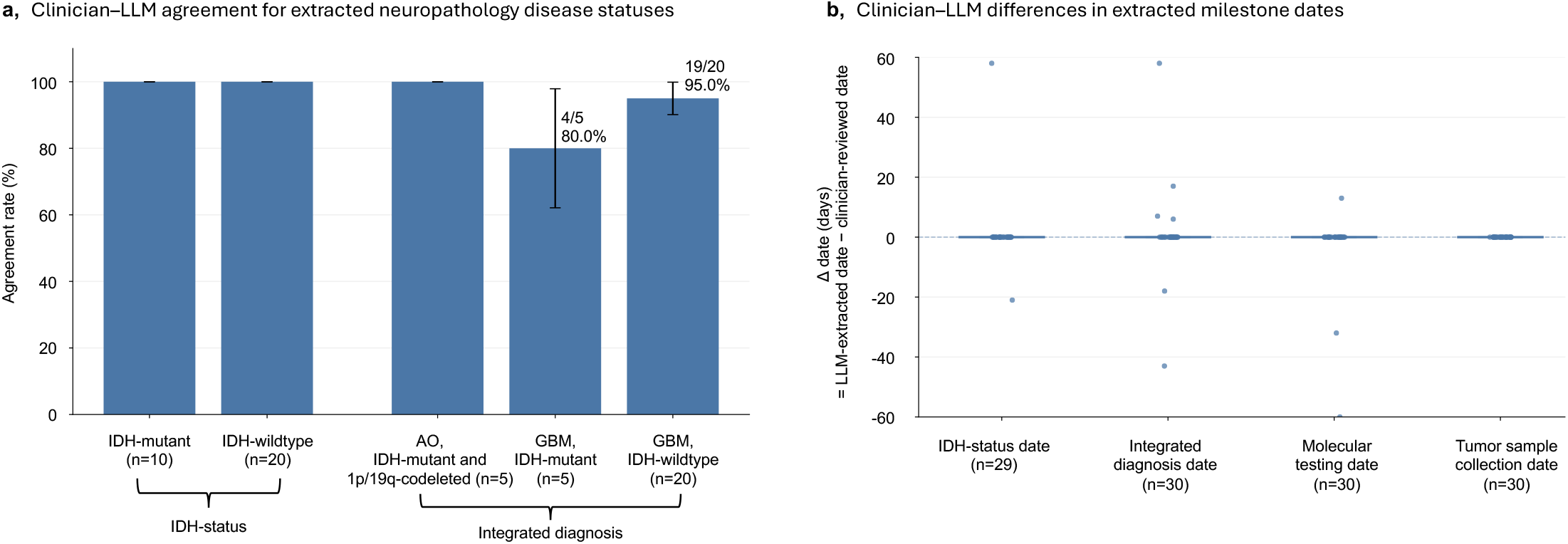
Clinician validation of LLM-extracted neuropathology work-up disease statuses and milestone dates. 30 patient cases were randomly selected from the patient-level reference cohort (*n* = 282) with complete disease-status and milestone-date information, using disease-status-stratified sampling to ensure adequate representation of less common disease statuses (see Methods). **a**, Agreement between LLM-extracted disease statuses and clinician review, stratified by the LLM-extracted IDH status and integrated diagnosis. Bars show the proportion of reviewed patient cases in agreement with clinician assessment. Error bars indicate standard errors. Numbers below each category denote the number of reviewed cases. **b**, Differences between LLM-extracted and clinician-adjudicated milestone dates for IDH status, integrated diagnosis, molecular testing, and tumor sample collection, calculated as the LLM-extracted date minus the clinician-adjudicated date. Points represent individual patients, and the horizontal reference line denotes zero-day difference. The median difference was zero days for all milestone-date categories. Individual points outside the displayed range of −60 to 60 days are not shown. See Supplementary Methods for additional details on clinician–LLM agreement and adjudication.

**Extended Data Fig. 3:**
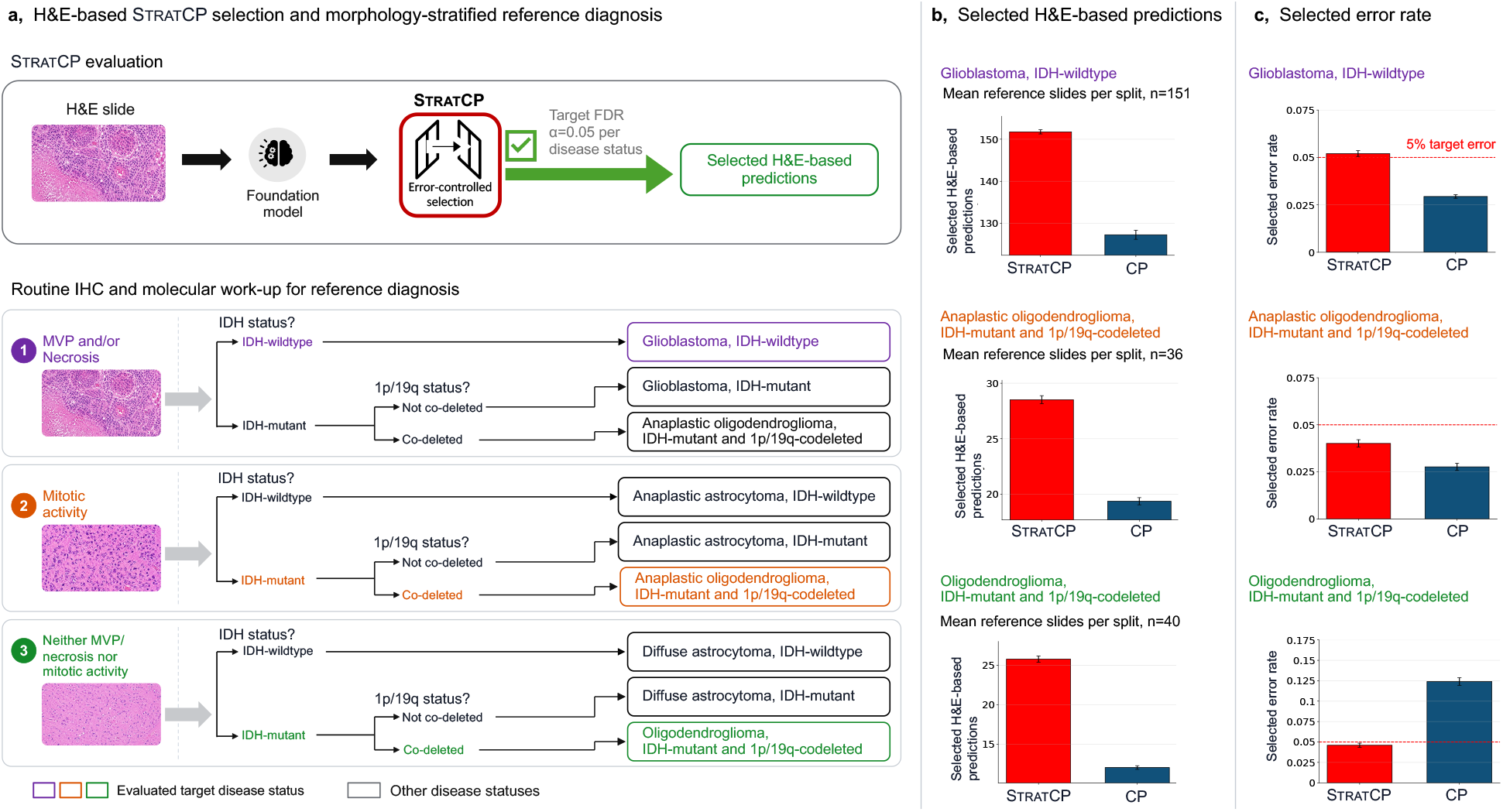
StratCP selects H&E-based disease-status predictions within morphology-stratified diagnostic pathways. **a**, Schematic of the retrospective evaluation using TCGA and EBRAINS H&E slides. A fixed FM-based H&E predictor generates disease-status scores from each slide, and StratCP applies error-controlled selection at a target error rate of *α* = 0.05 per disease status. Routine immunohistochemical (IHC) and molecular work-up defines the reference disease status against which selected predictions are evaluated. Reference diagnoses are stratified by the presence of microvascular proliferation (MVP) and/or necrosis, mitotic activity, or neither feature, followed by IDH and 1p/19q status. Colored outlines indicate the evaluated target disease status within each morphology stratum; gray outlines indicate other reference disease statuses. Concurrent loss of chromosome arms 1p and 19q is denoted as 1p/19q codeletion. **b**, Mean number of H&E-based predictions selected by StratCP and the conformal prediction baseline (`CP`) for glioblastoma, IDH-wildtype; anaplastic oligodendroglioma, IDH-mutant and 1p/19q-codeleted; and oligodendroglioma, IDH-mutant and 1p/19q-codeleted. `CP` selects a slide only when its conformal prediction set is a singleton. Disease-status headings report the mean number of reference slides per split. **c**, Selected error rates (i.e., selected-set FDR) for the same disease statuses. Red bars denote StratCP and blue bars denote CP; the dashed red line marks the prespecified 5% target error rate. Results are averaged across 500 repeated calibration/test splits, and error bars denote standard errors across splits.

**Extended Data Fig. 4:**
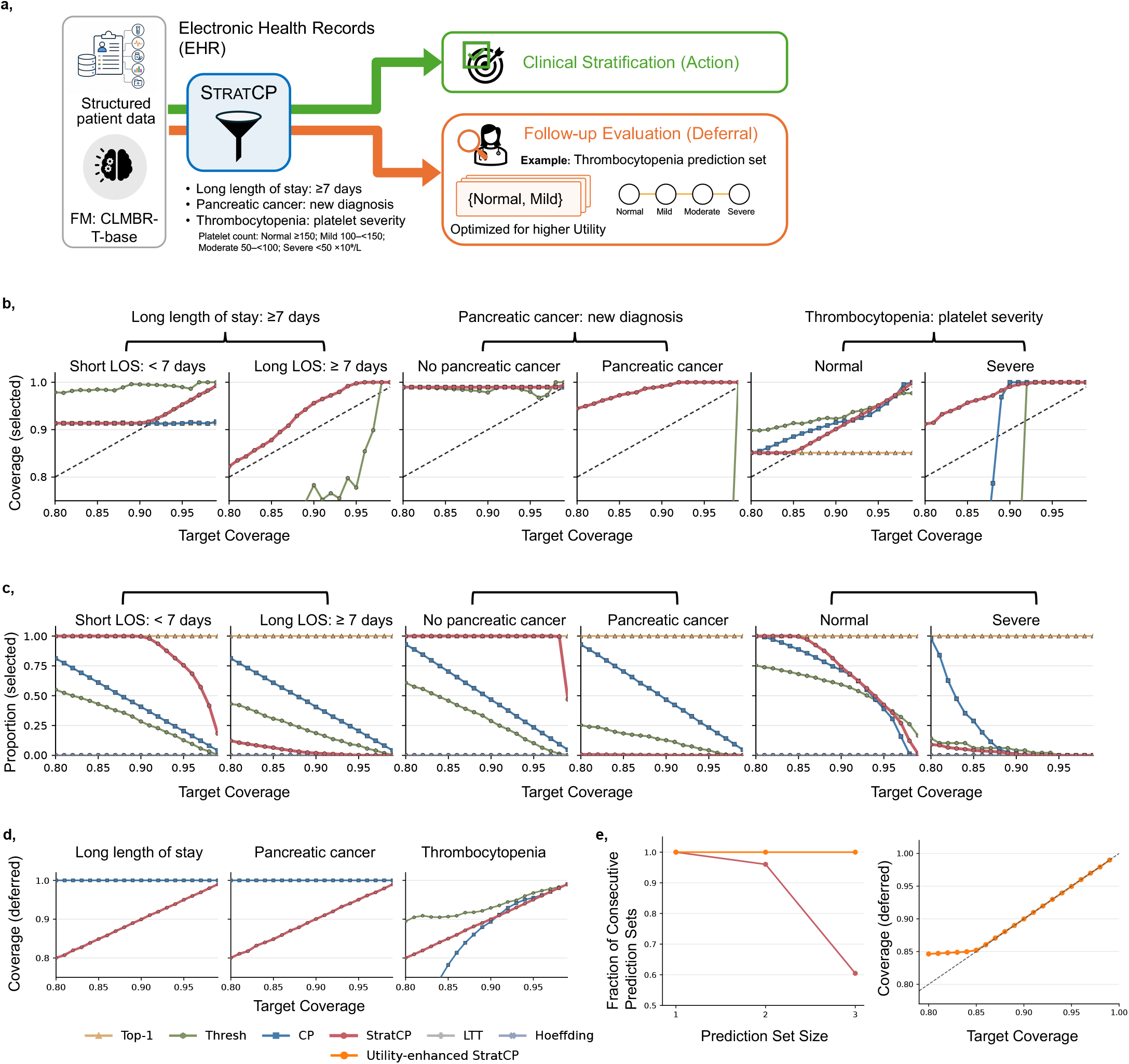
StratCP enables EHR-based clinical stratification with calibrated action and deferral decisions. **a**, Schematic of the EHR workflow. Structured longitudinal EHR events are encoded using the fixed CLMBR-T-base foundation model [37], and task-specific prediction heads generate probability scores that are evaluated with StratCP to select predictions for the action arm or defer patients for follow-up evaluation. For thrombocytopenia, deferred prediction sets are further optimized using an ordinal platelet-severity utility graph that favors clinically coherent sets containing neighboring severity statuses, such as {normal, mild}. **b**, Selected-set coverage (1− FDR) across target coverage levels for three EHR prediction tasks: length of stay with two statuses, new pancreatic-cancer diagnosis with two statuses, and thrombocytopenia severity with four statuses. **c**, Fraction of patients selected into the action arm within each predicted status, where the denominator is all patients whose top-1 model prediction corresponds to that status. All four thrombocytopenia statuses were evaluated; Mild and Moderate are omitted from the status-specific panels in **b–c** for visual clarity. **d**, Coverage among deferred patients, aggregated within each task, across target coverage levels. **e**, Utility-enhanced StratCP, shown in orange, increases the fraction of deferred thrombocytopenia prediction sets containing consecutive severity statuses across prediction-set sizes, while maintaining empirical deferred coverage close to the target level. Prediction sets of size four are omitted because they contain all four severity statuses and are therefore consecutive by definition.

## Online Methods

Online Methods are organized into three parts. Section 1 describes the datasets, clinical end-points, preprocessing procedures, foundation models, and task-specific predictors used across retinal imaging, neuro-oncology, and longitudinal EHR settings. Section 2 presents the core StratCP algorithms for selecting confident predictions in the action arm and constructing calibrated prediction sets for deferred patients or slides in the deferral arm, together with the corresponding finite-sample guarantees under the stated assumptions. This section also describes the optional utility-enhancement procedure, which modifies the deferral-arm conformity score to preferentially group clinically related disease statuses. Finally, Section 3 provides calibration, evaluation, and implementation details for StratCP applications.

### 1 Models, datasets, and task-specific predictors

We describe the foundation models, datasets, preprocessing procedures, and task-specific predictors used across retinal imaging, neuro-oncology, and longitudinal EHR tasks. The selection of the foundation models was primarily determined by public availability, and we focused on well-established benchmark tasks to demonstrate the general applicability of StratCP.

#### 1.1 Models and datasets for retinal disease tasks

We utilized the fine-tuned RETFound models and datasets provided by [1] for three retinal disease tasks. The diabetic retinopathy diagnosis task uses the Kaggle APTOS-2019 dataset [59], with disease statuses defined according to the International Clinical Diabetic Retinopathy Severity Scale, spanning five stages from no retinopathy to proliferative diabetic retinopathy. The glaucoma diagnosis task uses the Glaucoma Fundus dataset [60], with three disease statuses: Mild, Early, and Advanced. The eye condition diagnosis task uses the JSIEC dataset [48], which includes 1,000 images across 39 categories of common referable fundus diseases and conditions.

For each task, we followed the data preprocessing and splits used in the original study [1]. The fine-tuned RETFound models were treated as fixed and independent of the data used for StratCP calibration and evaluation. The original test fold was randomly split into two equally sized subsets: a calibration fold and a test fold. The calibration fold provided labeled data for StratCP, whereas ground-truth labels in the test fold were reserved for evaluation. Given a fixed fine-tuned model, StratCP uses only its predictions on the calibration and test folds and does not require hidden model representations or further model fitting.

#### 1.2 Models and datasets for neuro-oncology tasks

##### Datasets and WSI preprocessing

We used H&E-stained whole-slide images (WSIs) from TCGA-LGG and TCGA-GBM, obtained through the NIH GDC portal [43], and from the EBRAINS repository [44]. For TCGA, we included only FFPE diagnostic slides; the majority were scanned at 40× magnification, with a minority at 20×. EBRAINS slides were scanned at 40× and include expert-assigned WHO-based diagnostic labels spanning 30× CNS tumor types.

We segmented and patched WSIs using the CLAM pipeline [61], with the CLAM “TCGA” preset for TCGA and the “BWH biopsy” preset for EBRAINS. When a slide did not contain a downsample level equivalent to 20×, patches were extracted at 40× using a 512 × 512-pixel window and resized to 224 × 224 for feature extraction, yielding an effective magnification comparable to 20× [2]. Patch-level embeddings were extracted using the UNI foundation model [2].

##### Task-specific predictors

Across neuro-oncology tasks, UNI patch embeddings were aggregated using attention-based multiple instance learning (ABMIL) [58], with task-specific prediction heads trained for each endpoint. IDH mutation status was formulated as binary classification (IDH mutant versus wild-type), WHO-based CNS tumor subtyping as 30-class classification, and H&E-only diagnosis in adult-type diffuse glioma as branch-specific classification stratified by morphology. Classification heads were trained using cross-entropy loss. For time-to-mortality prediction, we used a log-normal accelerated failure time (AFT) survival head producing patient-specific 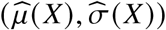 and optimized using a right-censoring-aware log-likelihood (Section 3.10).

##### Data splits and model selection

For each task, we partitioned the data into train/validation/conformal-calibration/test splits as follows: IDH mutation status (45/20/15/20%), WHO-based CNS tumor subtyping (45/20/15/20%), time-to-mortality prediction (45/20/20/15%), and H&E-only diagnosis in adult-type diffuse glioma (45/20/20/15%). Splitting was performed at the patient level to prevent leakage, ensuring that all slides from a given patient were assigned to a single split.

Hyperparameters were selected by random search based on validation performance with early stopping. The complete hyperparameter search spaces for the classification and time-to-mortality models are provided in Supplementary Methods. Task-specific experimental and evaluation details are provided in Section 3.

#### 1.3 Models and datasets for EHR tasks

##### Dataset and clinical endpoints

We used EHRSHOT [37], a benchmark of longitudinal electronic health records (EHRs) from 6,739 patients at Stanford Medicine. We evaluated three classification tasks spanning operational, diagnostic, and laboratory outcomes. Length of stay was defined as total hospital stay < 7 versus ≥ 7 days, with prediction at 23:59 on the day of admission and same-day discharges excluded. Pancreatic cancer was defined as a first pancreatic-cancer diagnosis within 365 days after inpatient discharge, with prediction at 23:59 on the day of discharge; encounters after a prior pancreatic-cancer diagnosis were excluded. Thrombocytopenia severity was defined from the next platelet measurement, with prediction immediately before the result was recorded, using four ordered classes: normal (≥ 150), mild (100–< 150), moderate (50–< 100), and severe (< 50 × 10^9^/L).

##### Foundation model and task-specific predictors

We used the released CLMBR-T-base [37], a 141-million-parameter foundation model pretrained on longitudinal structured EHR data. At each task-specific prediction time, CLMBR-T-base encodes the preceding sequence of coded clinical events, including demographics, diagnoses, procedures, laboratory measurements, medications, and other structured observations, into a 768-dimensional contextual patient representation. No clinical text or imaging data were used. The CLMBR-T-base backbone was kept fixed, and a task-specific logistic-regression head was trained on the frozen representations to generate class probabilities used by StratCP.

##### Model development splits

We retained the predefined EHRSHOT train, validation, and test partitions for downstream model development. The corresponding numbers of unique patients in the train/validation/test partitions were 1,377/1,240/1,238 for length of stay, 1,376/1,242/1,246 for pancreatic cancer, and 2,084/1,981/1,998 for thrombocytopenia. Task-specific heads were fit using the training partition with model selection on the validation partition; the predefined test partition was not used for model fitting or hyperparameter selection. Post-hoc calibration and evaluation of StratCP within the held-out test partition are described in Section 3.8.

### 2 STRATCP Algorithms

We describe the action and deferral algorithms underlying StratCP. Task-specific implementations are provided in Section 3.

#### 2.1 Notations

Throughout the paper, we consider a standard supervised learning setting where *X* ∈ *X* denotes the input features and *Y* ∈ *Y* the disease status of interest. For classification tasks (e.g., diabetic retinopathy diagnosis), *Y* is a finite set of categorical disease statuses; for regression tasks (e.g., survival prediction), we take *Y* = ℝ, the set of real-valued numbers.

Throughout StratCP, we take a pre-trained or fine-tuned foundation model, denoted as the function *f* : *X* → *O*, where *O* is the output space, which may differ from *Y* especially in classification problems. In classification problems, we assume that applying *f* to some feature value *x* ∈ *X* yields a vector *f* (*x*) ∈ Δ^|*Y*|^ of estimated probabilities, and with a slight abuse of notations, we assume *f* (*x, y*) is the predicted probability of label *y* ∈ *Y* given feature *x* ∈ *X*. In regression problems, we assume that applying *f* to some feature value *x* ∈ X yields a point prediction *f* (*x*) ∈ ℝ which predicts the unknown, continuous label.

We assume access to a labeled *calibration* set 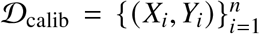, as well as a *test* set 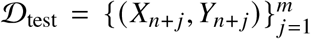, where *X*_*i*_ ∈ *X* is the feature and *Y*_*i*_ ∈ *Y* is the disease status of the *i*-th patient or slide. For brevity, we henceforth use the term patient to refer to each individual case, which may also correspond to an H&E slide in neuro-oncology tasks. In StratCP, only the calibration set and the test features 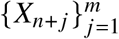 are available at inference, while the test disease status 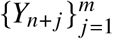 are reserved for evaluation.

The only assumptions needed in StratCP are (i) the calibration and test patients are independently and identically distributed (i.i.d.), and (ii) the training process of *f* is independent of *D*_calib_ and *D*_test_, so that *f* can be viewed as fixed whereas the calibration and test data are random.

### 2.2 Action arm: select predictions under a pre-specified budget

We next describe how StratCP selects confident predictions while controlling the false discovery rate (FDR), building on conformal selection methods [33, 36, 62]. Given a user-specified criterion defining a trustworthy prediction, StratCP selects a subset of test patients while controlling the expected proportion of selected predictions that violate this criterion.

#### High quality criterion

We begin with a user-specified criterion for a prediction to be “high quality” and can be directly trusted in deployment. The criterion is any function that can involve the foundation model *f*, the features *x* ∈ *X*, and the true disease status *y* ∈ *Y*. We denote the criterion as *I*( *f, x, y*) ∈ {0, 1}. For instance, in classification problems, we may take *I*( *f, x, y*) = **1**{*y* = argmax_*y*_′_∈Y_ *f* (*x, y*^′^)}, meaning that the prediction is confident if the top-1 predicted disease status coincides with the true disease status (unknown in the inference time). In regression problems (such as survival prediction), we may take I( *f, x, y*) = **1**{*y* ≥ *c*} indicating the true survival time *y* of the test patient exceeds a threshold *c* ∈ ℝ (i.e., exhibits favorable early survival). We aim to identify those test patients with *I*( *f, X*_*n*+*j*_, *Y*_*n*+*j*_) = 1 without observing *Y*_*n*+*j*_. Formally, we aim to select a subset *S* ⊆ {1, …, *m*}, such that

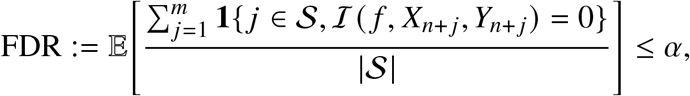

where *α* ∈ (0, 1) is a user-specified error level, and we use the convention 0/0 = 0. Intuitively, controlling the FDR below *α* means that on average, 1 − *α* fraction of the selected patients indeed satisfy the desired property.

##### Multiple criteria and feasibility groups (optional)

StratCP can accommodate multiple user-defined criteria by stratifying test patients into disjoint *feasibility groups*, and only those in the feasibility group will be considered for being selected to meet the corresponding criterion. Consider *K* distinct criteria *I*_*k*_ in total, which leads to *K* disjoint feasibility groups. For every *k* = 1, …, *K*, the membership in the *k*-th feasibility group is indicated by a binary function *G*_*k*_ (*f, x*) ∈ {0, 1}, and we denote the feasibility group of test patients as 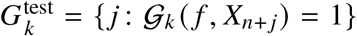 for *k* = 1, …, *K*. For instance, in classification problems, we might consider multiple criteria: the predicted disease status is *correctly* mild, the predicted disease status is *correctly* severe, and so on. For each of such disease statuses, we denote it as the fixed status value *y*_*k*_, the *k*-th disease status in *Y*. We may set 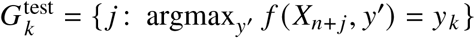 and *G*_*k*_ *f, x* = **1** {argmax_*y*_′ *f* (*X*_*n* + *j*_, *y*^′^) = *y*_*k*_} for the criterion *I*_*k*_ (*f, x, y*) = **1** {argmax_*y*_′ (*f X*_*n* + *j*_, *y*^′^) = *Y*_*n* + *j*_ = *y*_*k*_}. That is, for each disease status *y*_*k*_, only test patients whose top-1 prediction equals *y*_*k*_ can be selected under the criterion where the predicted disease status is *correctly* equal to *y*_*k*_. We consider multiple criteria in the tasks of diabetic retinopathy diagnosis, glaucoma diagnosis, and IDH mutation status prediction, where the number of disease statuses is relatively small. In eye condition diagnosis and CNS tumor subtyping tasks, we do not specify multiple feasibility groups due to the large number of disease statuses and limited sample sizes.

##### Scoring functions

Finally, StratCP employs any fixed *scoring* function *s*_*k*_ (*f, x*) ∈ ℝ for predicting *I*_*k*_ (*f, x, y*) based on the foundation model *f* and test features *x*. Roughly speaking, for a test sample *X*_*n*+*j*_, a higher score *s*_*k*_ (*f, X*_*n*+*j*_) should indicate a higher (predicted) likelihood of criterion *I*_*k*_ (*f, X*_*n*+*j*_, *Y*_*n*+*j*_). The error control of StratCP does not rely on the quality of such a score. Instead, the score provides uncalibrated, preliminary evidence for the reliability of a prediction, while StratCP calibrates the selections for FDR control. As an example, in classification problems (e.g., diabetic retinopathy diagnosis, glaucoma diagnosis, and IDH mutation status prediction) with *I*_*k*_ (*f, X*_*n*+*j*_, *Y*_*n*+*j*_) = **1** {argmax_*y*_′ *f* (*X*_*n* + *j*_, *y*^′^) = *Y*_*n*+*j*_} and *G*_*k*_ (*f, X*_*n*+*j*_) = **1** {argmax_*y*_′ *f* (*X*_*n*+*j*_, *y*^′^) = *y*_*k*_}, the choice of the score function in our experiments is *s*_*k*_ = (*f x, y*_*k*_), the predicted probability of the top-1 disease status *y*_*k*_.

##### Algorithm

StratCP leverages the conformal selection framework [36] to quantify the confidence in I_*k*_ ( *f, X*_*n*+*j*_, *Y*_*n*+*j*_) = 1 for each test patient in 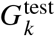 and calibrate whether the confidence level is strong enough to deem the prediction as high-quality. We denote 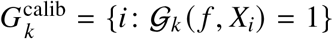 the set of calibration patients eligible for the *k*-th criterion. Then, for each test patient, we construct a conformal p-value following [36]:

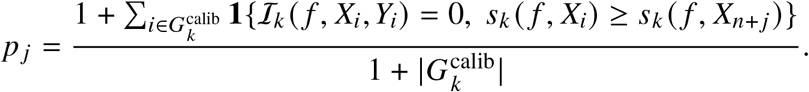

The p-values are then passed onward to the Benjamini–Hochberg procedure [63]. For each criterion *k*, we define the criterion-specific selected set 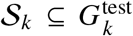 by taking *j* ∈ ***S***_*k*_ if and only if 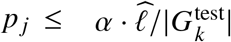, where 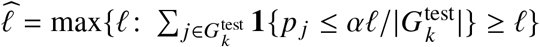. The overall action-arm selected set is then the union across criteria,

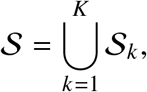

and test patients not in ***S*** are passed to the deferral arm.

The larger the score *s*_*k*_ (*f, X*_*n* + *j*_) is, the smaller the p-value *p* _*j*_ is, indicating a stronger confidence in the *j*-th test patient obeying the reliability criterion. The statistical validity and error control hinges on conformal inference and selective inference theory, which we do not elaborate here; see [36] for a detailed theoretical exposition. The general algorithm is summarized in Algorithm 1.

###### Algorithm 1

StratCP action arm: selection of confident predictions (criterion *k*)

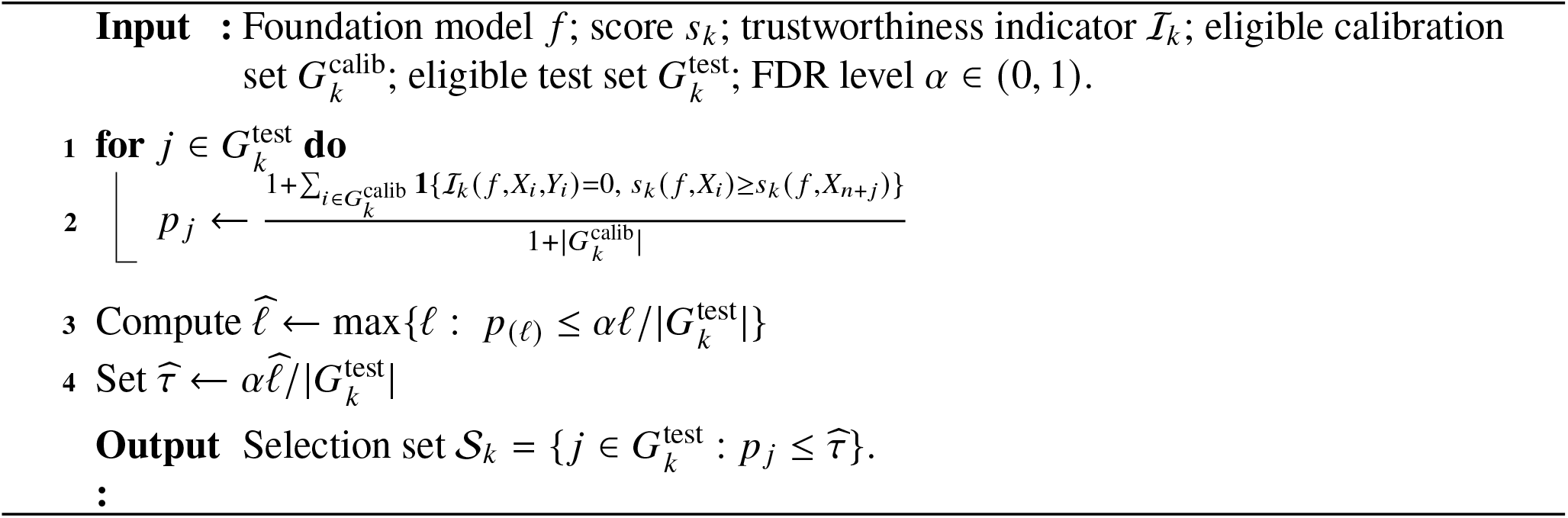

**Theoretical guarantee**. Recall that _*k*_ is the set of test patients deemed as confidently obeying the criterion _*k*_. The theory in [36] implies the following FDR control guarantee of StratCP in the first step.

###### Theorem 1.

Assume the two conditions specified at the end of Section 2.1. Then, for any foundation model *f* and any scoring functions {*s*_*k*_}, *k* = 1, …, *K*, it holds that

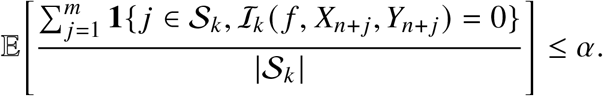

### 2.3 Deferral arm: constructing calibrated prediction set for deferred patients

In the deferral-arm step of StratCP, each test patient or slide that is not selected by the action arm will be issued a prediction set that quantifies a range of values the unknown disease status may take. It is guaranteed that with a prescribed probability, such as 1 − *α*, the true disease status falls within that range conditional on the patient not being selected in the first step of StratCP. The construction of such a prediction set builds upon the joint Mondrian conformal inference (JOMI) framework [35].

### Conformity score

We begin with the notion of conformity score [26] for defining the prediction set. Intuitively, given a foundation model *f* and a feature value *x*, a conformity score *V x, y* measures how well a disease status *y*∈ *Y* conforms to the model prediction. Popular choices of conformity score in multi-class classification problems include the adaptive prediction sets (APS) score [38], which is the score used in the five tasks in Fig. 3. Specifically, the APS score is

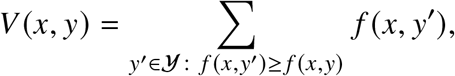

which is the cumulative predicted probability of all disease status with higher predicted probability than the status *y*. Given a conformity score *V* : X × Y → ℝ, our prediction set takes the form

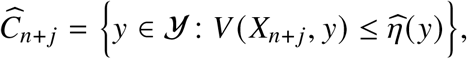

where 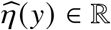 is a cutoff calibrated to achieve the desired coverage.

#### Algorithm

Following the JOMI framework [35], we calibrate the prediction set by searching for calibration patients that, when posited as a test patient, would also not be selected in the first step of StratCP. For each criterion *k*, let *A*_*k*_ denote the fixed selection mapping implemented by Algorithm 1. The corresponding criterion-specific selected set is

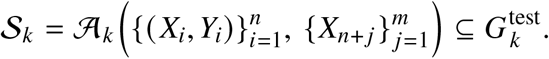

A prediction set will be issued for each 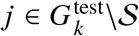.

We now explain the algorithm *A*_*k*_. Given any 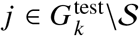, and for each hypothesized disease status *y* ∈ *Y*, we let 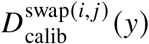 be the calibration set obtained by placing ( *X*_*n*+*j*_, *y*) in the place of the of the *i*-th calibration patient, and 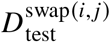 be the test set obtained by placing (*X*_*i*_, *Y*_*i*_) in the place *j*-th test patient. We then compute the set of reference calibration samples corresponding to *y* ∈ *Y* via

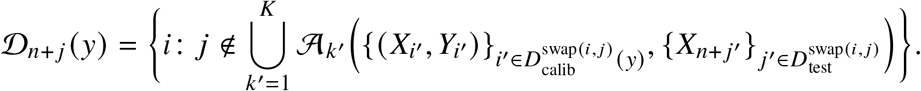

That is, *D*_*n*+*j*_ (*y*) collects those calibration patients which, when posited as the *j*-th test patient, would lead to the same selection decision (in the first step of StratCP). Finally, we define the prediction set for the *j*-th test patient at level 1 − *α* as

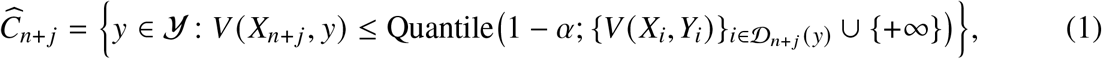

where Quantile (1− *α*; *D*) = {inf *z* ∈ ℝ: ∑_*d*∈ *D*_ **1** {*d* ≤ *z*} ≥ (1 − *α*) |*D*|} is the empirical (1− *α*)-th quantile. Intuitively, *Ĉ*_*n*_+_*j*_ is the collection of disease statuses such that the imputed test score falls within the high-frequency range defined by all the reference calibration scores. This general algorithm applies to both classification and regression problems, and is summarized in Algorithm 2.

##### Algorithm 2

StratCP deferral arm: Construction of prediction set for deferred patients

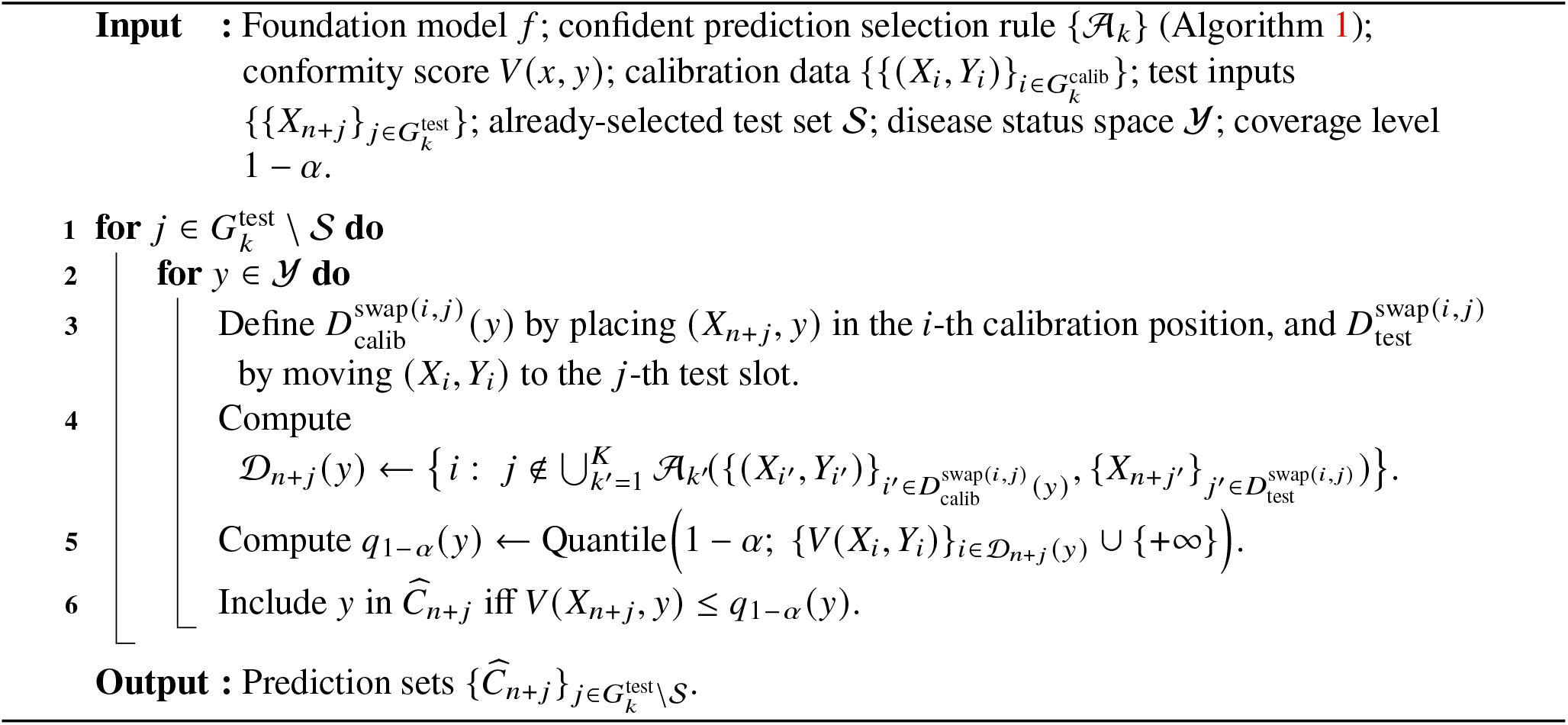

The computation of Algorithm 2 is not straightforward. In multi-class classification problems, it can be computed by repeatedly running the selection process and validating the conditions for each *y* ∈ *Y*. To address this issue, we implement equivalent versions of Algorithm 2 that can be efficiently computed for both classification and time-to-event regression problems. Details are provided in Supplementary Methods.

#### Theoretical guarantee

As an application of the theory of JOMI [35], we have the following finite-sample coverage guarantee for *Ĉ*_*n*+*j*_.

##### Theorem 2.

Assume the two conditions specified at the end of Section 2.1. Then, for any foundation model *f*, any action-arm scoring functions {*s*_*k*_} used in the selection, and any conformity score function *V*, the prediction set *Ĉ* _*n*+ *j*_ constructed in (1) obeys

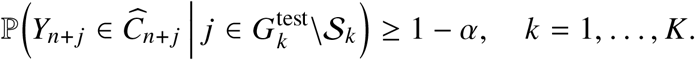

### 2.4 Utility enhancement for deferred prediction sets

For multiclass classification, StratCP can optionally incorporate clinical relationships among disease statuses by modifying the conformity score used in the deferral arm (Extended Data Fig. 1). Let *U* = (*Y, E*) denote a utility graph whose edge weights *E*_*y,y*_′ encode task-specific clinical relatedness.

Whereas the APS score orders disease statuses using predicted probability alone, the utility-enhanced procedure begins with the top-1 predicted disease status and iteratively ranks subsequent candidate statuses using the utility graph while restricting consideration to labels with high predicted probability. If *π*(1), …, *π*(*K*) denotes the resulting ordering, we define

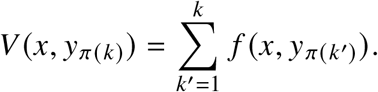

The resulting conformity scores are supplied to the same selection-conditional calibration procedure in Section 2.3. Thus, utility enhancement modifies prediction-set composition without changing the calibration procedure or target coverage.

The complete utility-ranking algorithm, task-specific utility graphs, and evaluation metrics are provided in Supplementary Methods.

## 3 Task-specific implementation and evaluation

### 3.1 Shared evaluation protocol for classification tasks

For all classification tasks, the trustworthiness criterion was correctness of the top-1 prediction,

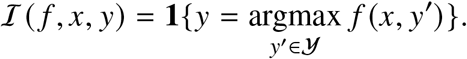

When disease-status-specific error control was used, patients or slides whose top-1 prediction was *y*_*k*_ formed the corresponding eligibility group, and StratCP was calibrated separately within each group using the predicted probability *f* (*x, y*_*k*_) as the confidence score. For eye-condition diagnosis and WHO-based CNS tumor subtyping, limited per-disease-status sample sizes precluded separate calibration for each disease status; selection was therefore calibrated globally across all top-1 predictions.

Unless otherwise specified, classification results were averaged over 500 repeated Monte Carlo stratified calibration/test splits. Splitting was performed at the patient level for patient-based cohorts, with all slides from the same patient kept within a single split. For the three retinal tasks, the original held-out RETFound test fold was stratified by disease status and divided equally into conformal-calibration and test folds in each repetition. For the primary neuro-oncology classification tasks, patient-level train/validation/conformal-calibration/test allocations were 45/20/15/20% for IDH mutation status and WHO-based CNS tumor subtyping and 45/20/20/15% for H&E-only diagnosis in adult-type diffuse glioma. For the H&E-only diagnosis task, 20% of the data were allocated to conformal calibration because of the relatively small held-out cohort. Exact task-specific sample counts are provided in Supplementary Table S11. For the EHR tasks, each repetition stratified patients in the predefined EHRSHOT test partition by disease status and divided them into 40% conformal-calibration and 60% evaluation sets. Cross-cohort and retrospective DFCI analyses used distinct evaluation designs described in their respective sections below.

The primary error level was *α* = 0.05. Action-arm performance was summarized by the false discovery proportion (FDP) among selected patients or slides and the number or proportion selected. FDR was estimated as the mean FDP across repetitions. Accordingly, the plotted coverage of selected predictions was defined as 1 − FDR, equivalently the mean of 1 − FDP across repetitions. For deferred patients or slides, we reported prediction-set coverage and mean prediction-set size. Overall marginal coverage was computed by assigning a singleton prediction set containing the top-1 prediction to selected patients or slides and the selection-conditionally calibrated prediction set to deferred patients or slides.

For deferred patients or slides, StratCP used the APS conformity score and the selection-conditional calibration procedure described in Section 2.3. We compared StratCP with `Top-1`, a probability-threshold prediction-set baseline (`Thresh`), and standard conformal prediction (`CP`) using APS scores. For action-arm selection, we additionally compared with learn-then-test (`LTT`) [42], which certifies a confidence threshold through hypothesis testing of the selected error rate, and a `Hoeffding` baseline, which certifies a threshold when a Hoeffding-type upper confidence bound on the selected error is below the target level [64]. Complete baseline definitions and implementation details are provided in Supplementary Methods.

### 3.2 Diabetic retinopathy diagnosis

For diabetic retinopathy, we used five disease-status-specific eligibility groups corresponding to the five severity levels and calibrated selection separately within each group. The primary analysis used *α* = 0.05; for the end-to-end coverage and efficiency analyses in Fig. 2d, we additionally evaluated *α* ∈ {0.025, 0.05, 0.1, 0.2}. Evaluation otherwise followed the shared classification protocol in Section 3.1. We additionally evaluated utility-enhanced prediction sets for deferred patients, using clinical adjacency of diabetic-retinopathy severity levels to encourage prediction sets containing related disease stages (Extended Data Fig. 1). The task-specific utility graph and evaluation procedure are provided in Supplementary Methods.

### 3.3 Glaucoma diagnosis

For glaucoma diagnosis, the three disease statuses were treated as separate disease-status-specific eligibility groups. Selection, deferral, and baseline evaluation otherwise followed the shared classification protocol in Section 3.1.

### 3.4 Eye condition diagnosis

The eye-condition task contains 39 disease statuses. Because of limited per-disease-status sample sizes, we did not calibrate separate disease-status-specific eligibility groups; instead, StratCP controlled the error rate globally among selected top-1 predictions. Action-arm, deferral-arm, and baseline evaluation otherwise followed Section 3.1. We additionally evaluated utility-enhanced prediction sets using similarity in downstream management actions across eye conditions (Extended Data Fig. 1). Construction of the management-action utility graph and the corresponding evaluation metrics are provided in Supplementary Methods.

### 3.5 IDH mutation status

We combined TCGA-LGG, TCGA-GBM, and EBRAINS slides with unambiguous IDH status. For TCGA, IDH status was derived from the TCGA molecular database following [65]. For EBRAINS, we included diagnoses for which IDH status was explicit. Disease-status mappings and sample counts are provided in Supplementary Table S4. The task was formulated as binary classification (IDH-mutant versus IDH-wildtype), with selection calibrated separately for the two predicted disease statuses at *α* = 0.05. Action-arm, deferral-arm, and baseline evaluation otherwise followed Section 3.1.

### 3.6 WHO-based CNS tumor subtyping

We used the full EBRAINS cohort with 30 expert-labeled WHO-based CNS tumor subtypes [45]. Subtype definitions and counts are provided in Supplementary Table S7. Because of limited per-disease-status sample sizes, selection was calibrated globally across top-1 predictions rather than separately for each subtype. Action-arm, deferral-arm, and baseline evaluation otherwise followed Section 3.1. We additionally evaluated utility-enhanced prediction sets based on WHO grade relationships among tumor subtypes (Extended Data Fig. 1). Subtypes without a unique numeric WHO grade were treated as ungraded for this analysis. Construction of the utility graph and eligibility for the grade-coherence evaluation are detailed in Supplementary Methods.

### 3.7 Cross-cohort calibration of a fixed IDH-status predictor

We evaluated whether StratCP could be calibrated after transfer of a fixed IDH-status predictor from a source to an independent target cohort. We trained a separate UNI-based ABMIL IDH-status predictor exclusively on TCGA, using 320 patients (589 slides) for training and 142 patients (293 slides) for validation and model selection. An independent TCGA holdout set comprised 250 patients (491 slides). Splits were patient-level, with no patient appearing in more than one partition. The predictor was then treated as fixed throughout the cross-cohort analysis, and no EBRAINS samples were used for model training, fine-tuning, or selection. Cross-cohort predictive performance of the fixed UNI-based IDH-status predictor is reported in Supplementary Tables S5– S6.

The target cohort comprised 795 EBRAINS patients (873 slides) with known IDH status. In each of 500 Monte Carlo repetitions, EBRAINS patients were stratified by IDH status and randomly divided equally into a calibration pool and a held-out test cohort. The test cohort contained 398 patients in each repetition (147 IDH-mutant and 251 IDH-wildtype). All slides from these patients were retained for evaluation (mean, 437 slides per repetition). From the remaining patients, we sampled without replacement exactly 50, 100, 200, 300, or 400 EBRAINS slides for target-cohort calibration. Calibration and test patients were disjoint, the held-out test cohort was fixed across calibration-set sizes within each repetition, and calibration subsets of different sizes were sampled independently rather than nested.

For each target-cohort calibration set, we applied the same disease-status-specific StratCP procedure as in the primary IDH-status analysis, separately calibrating IDH-mutant and IDH-wildtype selections at *α* = 0.05 for the action arm and constructing APS-based prediction sets for deferred slides. We evaluated selected-set coverage (1− FDR), the number of selected slides, and deferral-arm coverage on held-out EBRAINS slides across the 500 repetitions. `CP, LTT`, and `Hoeffding` were implemented as described in the shared classification protocol and Supplementary Methods. For Fig. 4b, patient-level representations were obtained by averaging slide embeddings within each patient. Dimensionality-reduction and UMAP parameters are provided in Supplementary Methods.

### 3.8 StratCP evaluation of EHR FM predictions

For each EHR task, the trained CLMBR-T-base predictor and task-specific head were treated as fixed. In each of 500 repetitions, patients in the predefined EHRSHOT test partition were stratified by disease status and randomly divided at the patient level into 40% conformal-calibration and 60% evaluation sets, following the repeated Monte Carlo stratified splitting procedure described in Section 3.1. Neither subset was used to fit or tune the foundation model or downstream prediction head. Predictive performance of the CLMBR-T-base–based task-specific predictors is reported in Supplementary Tables S8–S9.

Selection was calibrated separately for each predicted disease status, corresponding to both length-of-stay statuses, both pancreatic-cancer disease statuses, and all four thrombocytopenia severity statuses. Deferred prediction sets used the APS conformity score, and baseline methods followed Section 3.1. The primary analysis used *α* = 0.05. To evaluate performance across error budgets, we additionally varied *α* from 0.01 to 0.20 in increments of 0.01 and reported selected-set coverage (1− FDR), the proportion of evaluation patients selected, and coverage among deferred patients.

For thrombocytopenia, we additionally evaluated utility-enhanced prediction sets using the ordered normal–mild–moderate–severe severity scale, encouraging clinically contiguous severity ranges among deferred prediction sets (Extended Data Fig. 4e). The task-specific utility graph, definition of consecutiveness, and visualization criteria are provided in Supplementary Methods.

### 3.9 H&E-only diagnostic triage in adult-type diffuse glioma

We evaluated whether selected adult-type diffuse glioma diagnoses could be resolved from H&E morphology alone under a pre-specified error budget, reflecting a workflow in which initial H&E assessment narrows the differential diagnosis before molecular testing. TCGA and EBRAINS diagnostic labels were harmonized across WHO classifications, and slides were stratified into morphology-defined branches according to the presence of microvascular proliferation or necrosis, mitotic activity without these features, or neither feature. Separate UNI–ABMIL classifiers were trained within each branch. Full label harmonization, branch-specific diagnostic disease statuses, and slide counts are provided in Supplementary Methods and Supplementary Table S11. For the branch with neither microvascular proliferation/necrosis nor mitotic activity, only two disease statuses were considered because the cohort contained no diffuse astrocytoma, IDH-wildtype slides.

We applied disease-status-specific StratCP selection to each branch at *α* = 0.05. Using the 45/20/20/15% patient-level train/validation/conformal-calibration/test split described in Section 3.1, we estimated FDR and the number of selected slides over 500 repeated evaluations. Because this analysis specifically asked which diagnoses could be resolved from H&E alone, only the action arm was evaluated; deferred prediction sets were not constructed.

### 3.10 H&E-based survival prediction in adult-type diffuse glioma

In H&E-based survival prediction, we sought to identify patients with favorable early survival while providing reliable survival-time information for patients not selected for action. Such stratified predictions could support risk-adapted follow-up, monitoring, and clinical decision-making. We defined favorable early survival as survival for ≥ 18 months, a clinically relevant horizon exceeding the typical median overall survival of glioblastoma, an aggressive adult-type diffuse glioma subtype, under contemporary standard-of-care therapy [52]. Throughout this analysis, we set *c* = 18 months.

#### Study design and survival model

We used TCGA-LGG and TCGA-GBM [43], as patient-level survival follow-up required for this analysis was not available for EBRAINS [44]. For patient *i*, let *T*_*i*_ > 0 denote the true time to mortality and *C*_*i*_ > 0 the censoring time. We observe *Y*_*i*_ = min (*T*_*i*_, *C*_*i*_) and Δ_*i*_ = **1** { *T*_*i*_ ≤ *C*_*i*_}, where Δ_*i*_ = 1 denotes an observed death and Δ_*i*_ = 0 indicates that death was not observed before follow-up ended, for example because the patient remained alive at last follow-up or follow-up otherwise ended. Administrative censoring was applied at *t*_max_ = 5 years.

UNI patch embeddings [2] were aggregated using ABMIL [58], and the resulting patient-level representation was concatenated with age and biological sex. A log-normal accelerated failure time (AFT) head produced patient-specific location and scale parameters 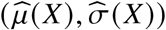, inducing the conditional CDF

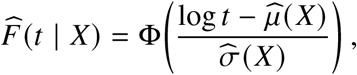

where F is the standard normal CDF. The survival head was trained by maximizing the right-censoring-aware log-likelihood

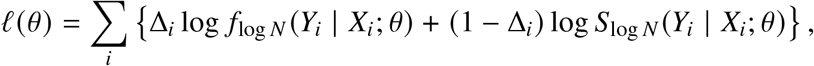

where *θ* denotes the trainable model parameters, *f*_log *N*_ (*t* | *X*; *θ*) is the conditional log-normal density of survival time, and 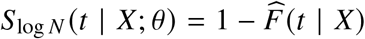 is the corresponding survival probability beyond time *t*. Thus, a patient with an observed death contributes the model probability density at the observed death time, whereas a right-censored patient contributes the probability of surviving beyond the last observed follow-up time. Data were partitioned at the patient level into 45/20/20/15% train/validation/conformal-calibration/test splits, with model selection based on validation negative log-likelihood and early stopping. Complete model-training and hyperparameter-search details are provided in Supplementary Methods.

#### Action arm under right censoring

For the action arm, we defined the favorable-early-survival score

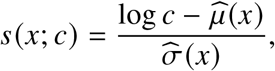

where smaller values correspond to a higher model-implied probability of surviving beyond the horizon *c*. For a candidate threshold *τ*, patients with *s* (*X*_*i*_; *c*) ≤ *τ* are therefore considered candidates for the favorable-early-survival action arm.

Because survival before *c* may be unobserved for censored patients, we adjusted action-arm calibration using inverse probability of censoring weighting (IPCW). Let

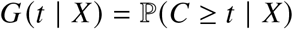

denote the probability that a patient with covariates *X* remains uncensored through time *t*. We estimated *G* by *Ĝ* using a Cox proportional hazards model [66] fit to the censoring process and defined the IPCW error surrogate

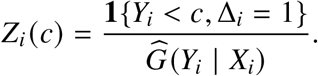

The numerator identifies an observed death before *c*, whereas inverse weighting by the estimated probability of remaining uncensored up to that time accounts for comparable deaths that may be unobserved because of censoring.

Using the IPCW error surrogates *Z*_*i*_ *c*, we calibrated a score threshold 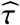 by evaluating candidate thresholds over the pooled calibration and test scores and selecting the largest threshold whose finite-sample-adjusted estimated error did not exceed *α*. The resulting action arm was

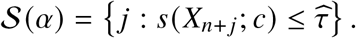

Thus, the procedure selects the most inclusive favorable-survival action rule supported at the target error level. The exact finite-sample adjustment and pooled-score threshold search are provided in Supplementary Methods.

The censoring model was cross-fit within the conformal-calibration cohort to obtain held-out calibration weights and then refit on the full calibration cohort for test-set evaluation. Under right censoring, action-arm error properties were evaluated empirically using IPCW.

#### Deferral arm

Patients not selected into the action arm were assigned a one-sided lower prediction bound on survival time using JOMI-based selection-conditional calibration [35]. For a deferred test patient *j*, the calibration reference set can depend on the hypothesized survival outcome because that outcome may alter the counterfactual action-arm selection. Since the action criterion

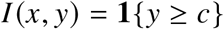

depends on the candidate survival time *y* only through whether *y* < *c* or *y* ≥ *c*, we construct two candidate-region-specific reference sets,

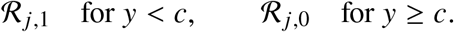

The efficient computation of these outcome-specific reference sets is detailed in Supplementary Algorithm 4.

Each reference set contains calibration patients whose counterfactual swap into test position *j* would leave that position deferred under the corresponding hypothesized outcome. Thus, the continuous outcome space reduces to two selection regimes rather than requiring enumeration over candidate survival times.

For calibration patient *i*, we use the one-sided conformity score

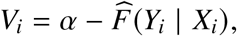

where 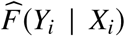 is the fitted survival CDF evaluated at the observed follow-up time. For each candidate-outcome region *b* ∈ {0, 1}, the selection-conditionally calibrated cutoff is

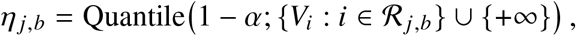

with corresponding model-CDF probability level

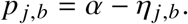

Using the fitted quantile function

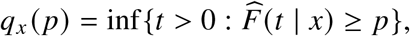

we obtain the region-specific lower-bound component

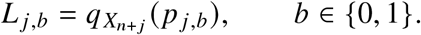

Because 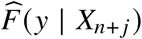 is nondecreasing in *y*, the two outcome regions define the raw prediction set

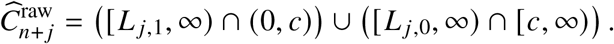

For reporting, we return the smallest closed upper interval containing this set,

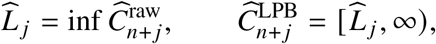

so that each deferred patient receives a single lower prediction bound. The exact reference-set construction and efficient implementation are provided in Supplementary Methods.

Under right censoring, deferral-arm coverage was evaluated empirically using inverse probability of censoring weighting (IPCW). The censoring-adjusted analyses assume exchangeability of calibration and test patients, conditional independent censoring *T* ⊥*C* | *X*, and positivity of the probability of remaining uncensored at times for which IPCW weights are evaluated.

##### Evaluation and baselines

Following the repeated Monte Carlo stratified splitting procedure used throughout the task-specific evaluations, we evaluated StratCP over 500 patient-level conformal-calibration/test splits and recalibrated StratCP for each evaluation. Within each evaluation, action-arm performance was summarized by the IPCW-adjusted FDP among patients assigned favorable early survival. FDR was estimated as the mean FDP across the 500 evaluations, and the plotted coverage of selected patients was defined as 1 − FDR, equivalently the mean of 1 − FDP across evaluations. For deferred patients, we evaluated IPCW-adjusted coverage of the lower prediction sets 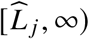. We additionally evaluated overall marginal coverage by assigning [*c*, ∞) to action-arm patients and 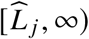, to deferred patients. Prediction-set efficiency was summarized using the interval length up to the administrative study end time *t*_max_ = 5 years, with lower endpoint *c* for action-arm patients and 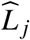 for deferred patients. Detailed IPCW evaluation estimators are provided in Supplementary Methods.

We compared StratCP with three survival baselines. `Top-1` assigned favorable early survival when the model-implied survival probability at *c* was at least 0.5. Standard conformal prediction (`CP`) constructed a marginal conformal lower prediction bound for every patient and assigned favorable early survival when this bound was at least *c*. The log-normal quantile baseline (`LNQ`) used the AFT model directly without conformal calibration, assigning favorable early survival according to its model-implied survival probability and using the corresponding parametric lower quantile for deferred patients. Complete baseline definitions are provided in Supplementary Methods.

##### Kaplan–Meier curves and grade stratification

For *α* = 0.05, we defined the action-arm group as the unique union of patients selected across the 500 evaluations, with each patient included only once. All remaining patients comprised the deferral group. We constructed Kaplan–Meier curves using each patient’s observed (*Y*, Δ) pair and assessed survival differences between the two groups using a log-rank test. Grade-specific analyses were performed after restricting to WHO CNS5 subtypes following molecular-data-based diagnostic remapping [65]. Grade 4 comprised glioblastoma, IDH-wildtype and astrocytoma, IDH-mutant, grade 4. Kaplan–Meier analyses were implemented using `lifelines` (v0.30.0).

### 3.11 Retrospective DFCI evaluation during pathology work-up

#### Cohort and clinical-note extraction

We retrospectively evaluated StratCP in 705 eligible adults evaluated at Dana-Farber Cancer Institute (DFCI) for suspected high-grade adult-type diffuse glioma. A total of 8,830 de-identified clinical notes (7,363 progress notes and 1,467 pathology/cytology notes) were grouped by patient and processed using GPT-4o through the Azure OpenAI Responses API (API version `2025-04-01-preview`; temperature 0) with a fixed structured-extraction prompt. For each patient, an anchor note was selected within the interval from 7 days before through 60 days after tumor sample collection when available. The anchor and at most the next 19 notes in chronological order were retained, yielding a maximum of 20 notes per patient. The model was instructed to use only explicit evidence in the supplied notes and to return structured patient-level information with source-note evidence. We extracted IDH status, molecular-testing results, integrated diagnosis, and their associated dates. Notes were ordered by date and empty notes were excluded. Each note included its report identifier, date, report type, procedure description, and de-identified text. Inputs exceeding the model context limit were divided at note boundaries, extracted separately, and reconciled into a patient-level structured output. The complete extraction prompt and output schema are provided in Supplementary Methods.

#### Diagnostic timelines and whole-slide image linkage

For timeline construction, we defined a harmonized tumor sample collection time using the extracted surgery or biopsy date when it occurred within six months of the structured sample-collection date and otherwise used the structured date. This value defined day 0 for all subsequent analyses. The IDH-status date was the earliest qualifying IDH immunohistochemistry, the molecular-testing date was the molecular-result date, and the integrated-diagnosis date was the corresponding diagnostic sign-out date. Time to each milestone was recomputed as the calendar-day difference from day 0. Whole-slide images (WSIs) were linked to patients and specimens using patient, sample, and block identifiers. When multiple eligible WSIs were available for a patient, we selected one deterministically by prioritizing a primary-tumor sample, followed by the slide whose procedure date was closest to tumor sample collection, with slide identifier used as the final tie-breaker. The integrated-diagnosis and IDH-status modeling cohorts contained 350 and 348 patients, respectively. Of the 348 patients in the IDH-status cohort, 284 had a valid IDH-status date for the corresponding diagnostic-timeline analyses. A total of 282 patients had available integrated-diagnosis, IDH-status, and molecular-result dates and constituted the clinician-review-eligible cohort. Detailed diagnostic harmonization, reference-cohort construction, whole-slide image linkage, and one-slide-per-patient selection are provided in Supplementary Methods.

#### H&E-based predictors and temporal evaluation

Both H&E-based predictors were trained exclusively on TCGA and EBRAINS and were kept fixed before application to DFCI. No DFCI data were used for model training, hyperparameter selection, or checkpoint selection. DFCI H&E WSIs were segmented and tiled using the CLAM “BWH biopsy” preset, with 256× 256-pixel patches extracted at 20× magnification. WSIs were represented using UNI-derived patch embeddings and aggregated with task-specific ABMIL classifiers trained using cross-entropy loss for IDH status and integrated diagnosis. To emulate prospective deployment, patients were divided chronologically by tumor sample collection date. For integrated diagnosis, patients from January 3, 2014 through September 5, 2017 formed the calibration pool and patients from September 6, 2017 through March 4, 2019 formed the fixed held-out test cohort. The calibration set contained 200 patients and the test cohort contained 150 patients. The IDH-status analysis used the corresponding H&E-aligned cohort under the same temporal partition, yielding 199 calibration and 149 test patients.

#### StratCP evaluation

For the primary analysis, we applied the same disease-status-specific StratCP selection procedure described for IDH-status prediction (Section 3.5) at *α* = 0.05, using the maximal temporal calibration set. Selection was calibrated separately for each predicted disease status. Standard `CP` with APS prediction sets served as the comparator, with singleton prediction sets treated as selected predictions. Because the DFCI analysis used a single prespecified temporal split rather than repeated random splits, we report the selected error rate, defined as the proportion of selected patients whose predicted disease status was incorrect, rather than an FDR averaged across repeated splits. For each task, method, and predicted disease status, uncertainty in both the selected error rate and the number of selected patients was quantified using 2,000 patient-level bootstrap replicates with replacement. Error bars show ±1 bootstrap standard error, computed as the standard deviation of the finite bootstrap estimates across replicates.

#### Clinician validation and displayed timelines

To assess LLM-based pathology-report extraction quality, 30 patient cases were randomly selected from the clinician-review-eligible cohort (*n* = 282) with available integrated-diagnosis, IDH-status, and molecular-result dates. Sampling was stratified by integrated diagnosis, with at least five cases from each less common diagnosis disease status, and subsequently adjusted by random replacement to ensure inclusion of 10 IDH-mutant patients. The final sample comprised 20 glioblastoma, IDH-wildtype, 5 glioblastoma, IDH-mutant, and 5 anaplastic oligodendroglioma, IDH-mutant and 1p/19q-codeleted cases. A board-certified clinician at the Dana-Farber Cancer Institute, blinded to the LLM outputs, reviewed the same clinical records provided to the LLM and independently assessed disease statuses and associated milestone dates. Disagreements were subsequently reviewed unblinded and adjudicated, with agreement evaluated for categorical disease status and calendar-day differences between LLM-extracted and clinician-adjudicated milestone dates. For Fig. 6c, the 25 displayed patients were randomly selected from patients chosen by StratCP for both IDH-status and integrated-diagnosis and with the relevant diagnostic intervals between 0 and 60 days. Diagnostic-interval summaries in Fig. 6d were computed from the full eligible cohort with the corresponding milestone available, rather than from the 25 displayed patients.

## References

1. Zhou, Y. et al. A foundation model for generalizable disease detection from retinal images. Nature 622, 156–163 (2023).

2. Chen, R. J. et al. Towards a general-purpose foundation model for computational pathology. Nature medicine 30, 850–862 (2024).

3. Xu, H. et al. A whole-slide foundation model for digital pathology from real-world data. Nature 630, 181–188 (2024).

4. Renc, P. et al. Zero shot health trajectory prediction using transformer. NPJ digital medicine 7, 256 (2024).

5. Fallahpour, A., Alinoori, M., Ye, W., Cao, X., Afkanpour, A. & Krishnan, A. Ehrmamba: Towards generalizable and scalable foundation models for electronic health records. arXiv preprint 2405.14567 (2024).

6. Makarov, N. et al. Large language models forecast patient health trajectories enabling digital twins. npj Digital Medicine 8, 588 (2025).

7. Kraljevic, Z. et al. Foresight—a generative pretrained transformer for modelling of patient timelines using electronic health records: a retrospective modelling study. The Lancet Digital Health 6, e281–e290 (2024).

8. Jiang, L. Y. et al. Health system-scale language models are all-purpose prediction engines. Nature 619, 357–362 (2023).

9. Waxler, S. et al. Generative medical event models improve with scale. 2508.12104 (2025).

10. Shmatko, A. et al. Learning the natural history of human disease with generative transformers. Nature, 1–9 (2025).

11. Campanella, G. et al. Real-world deployment of a fine-tuned pathology foundation model for lung cancer biomarker detection. Nature Medicine, 1–9 (2025).

12. Kondepudi, A. et al. Health system learning enables generalist neuroimaging models. Nature Medicine, 1–7 (2026).

13. Kompa, B., Snoek, J. & Beam, A. L. Second opinion needed: communicating uncertainty in medical machine learning. NPJ Digital Medicine 4, 4 (2021).

14. Ehrmann, D. E., Joshi, S., Goodfellow, S. D., Mazwi, M. L. & Eytan, D. Making machine learning matter to clinicians: model actionability in medical decision-making. NPJ Digital Medicine 6, 7 (2023).

15. Kim, Y.-T. et al. Defining operational safety in clinical artificial intelligence systems. NPJ Digital Medicine 9, 281 (2026).

16. Wong, A. et al. External validation of a widely implemented proprietary sepsis prediction model in hospitalized patients. JAMA internal medicine 181, 1065–1070 (2021).

17. Obermeyer, Z., Powers, B., Vogeli, C. & Mullainathan, S. Dissecting racial bias in an algorithm used to manage the health of populations. Science 366, 447–453 (2019).

18. Chua, M. et al. Tackling prediction uncertainty in machine learning for healthcare. Nature Biomedical Engineering 7, 711–718 (2023).

19. Peng, Y. et al. Enhancing AI reliability: A foundation model with uncertainty estimation for optical coherence tomography-based retinal disease diagnosis. Cell Reports Medicine 6 (2025).

20. Zhao, J. et al. Uncertainty-aware ensemble of foundation models differentiates glioblastoma from its mimics. Nature Communications 16, 8341 (2025).

21. Kendall, A. & Gal, Y. What uncertainties do we need in bayesian deep learning for computer vision? Advances in neural information processing systems 30 (2017).

22. Guo, C., Pleiss, G., Sun, Y. & Weinberger, K. Q. On calibration of modern neural networks. In International conference on machine learning 1321–1330 (2017).

23. Ovadia, Y. et al. Can you trust your model’s uncertainty? evaluating predictive uncertainty under dataset shift. Advances in neural information processing systems 32 (2019).

24. Kömen, J. et al. Towards robust foundation models for digital pathology. Nature Communications 17, 5218 (2026).

25. Muneer, A. et al. Foundation models in biomedical imaging: turning hype into reality. Nature Biomedical Engineering 10, 1557–1575 (2026).

26. Vovk, V., Gammerman, A. & Shafer, G. Algorithmic learning in a random world (Springer, 2005).

27. Angelopoulos, A. N. & Bates, S. A gentle introduction to conformal prediction and distribution-free uncertainty quantification. arXiv preprint 2107.07511 (2021).

28. Zhang, X. et al. Implementing trust in non-small cell lung cancer diagnosis with a conformalized uncertainty-aware AI framework. Nature Biomedical Engineering, 1–18 (2026).

29. Vazquez, J. & Facelli, J. C. Conformal prediction in clinical medical sciences. Journal of Healthcare Informatics Research 6, 241–252 (2022).

30. Olsson, H. et al. Estimating diagnostic uncertainty in artificial intelligence assisted pathology using conformal prediction. Nature communications 13, 7761 (2022).

31. Sreenivasan, A. P. et al. Conformal prediction enables disease course prediction and allows individualized diagnostic uncertainty in multiple sclerosis. npj Digital Medicine 8, 224 (2025).

32. Shen, Z., Chakraborti, T., Banerji, C. R. & Ding, X. Conformal prediction quantifies wearable cuffless blood pressure with certainty. Scientific Reports 15, 26697 (2025).

33. Gui, Y., Jin, Y. & Ren, Z. Conformal alignment: Knowing when to trust foundation models with guarantees. Advances in Neural Information Processing Systems 37, 73884–73919 (2024).

34. Mehrtens, H., Bucher, T. & Brinker, T. J. Pitfalls of conformal predictions for medical image classification. In International workshop on uncertainty for safe utilization of machine learning in medical imaging 198–207 (2023).

35. Jin, Y. & Ren, Z. Confidence on the focal: Conformal prediction with selection-conditional coverage. Journal of the Royal Statistical Society Series B: Statistical Methodology, qkaf016 (2025).

36. Jin, Y. & Candès, E. J. Selection by prediction with conformal p-values. Journal of Machine Learning Research 24, 1–41 (2023).

37. Wornow, M., Thapa, R., Steinberg, E., Fries, J. & Shah, N. Ehrshot: An ehr benchmark for few-shot evaluation of foundation models. Advances in Neural Information Processing Systems 36, 67125–67137 (2023).

38. Romano, Y., Sesia, M. & Candes, E. Classification with valid and adaptive coverage. Advances in neural information processing systems 33, 3581–3591 (2020).

39. Vorontsov, E. et al. A foundation model for clinical-grade computational pathology and rare cancers detection. Nature medicine 30, 2924–2935 (2024).

40. Shi, D. et al. A multimodal visual–language foundation model for computational ophthalmology. npj Digital Medicine 8, 381 (2025).

41. Guo, L. L. et al. A multi-center study on the adaptability of a shared foundation model for electronic health records. NPJ digital medicine 7, 171 (2024).

42. Angelopoulos, A. N., Bates, S., Candès, E. J., Jordan, M. I. & Lei, L. Learn then test: Calibrating predictive algorithms to achieve risk control. The Annals of Applied Statistics 19, 1641–1662 (2025).

43. Heath, A. P. et al. The NCI genomic data commons. Nature genetics 53, 257–262 (2021).

44. Roetzer-Pejrimovsky, T. et al. The digital brain tumour atlas, an open histopathology resource. Scientific Data 9, 55 (2022).

45. Louis, D. N. et al. The 2016 World Health Organization classification of tumors of the central nervous system: a summary. Acta neuropathologica 131, 803–820 (2016).

46. Ohno-Matsui, K. et al. International photographic classification and grading system for myopic maculopathy. American journal of ophthalmology 159, 877–883 (2015).

47. Ueta, T., Makino, S., Yamamoto, Y., Fukushima, H., Yashiro, S. & Nagahara, M. Pathologic myopia: an overview of the current understanding and interventions. Global health & medicine 2, 151–155 (2020).

48. Cen, L.-P. et al. Automatic detection of 39 fundus diseases and conditions in retinal photographs using deep neural networks. Nature communications 12, 4828 (2021).

49. Price, M. et al. CBTRUS statistical report: primary brain and other central nervous system tumors diagnosed in the United States in 2017–2021. Neuro-oncology 26, vi1–vi85 (2024).

50. Weller, M. et al. EANO guidelines on the diagnosis and treatment of diffuse gliomas of adulthood. Nature reviews Clinical oncology 18, 170–186 (2021).

51. Louis, D. N. et al. The 2021 WHO classification of tumors of the central nervous system: a summary. Neuro-oncology 23, 1231–1251 (2021).

52. Stupp, R. et al. Effect of tumor-treating fields plus maintenance temozolomide vs maintenance temozolomide alone on survival in patients with glioblastoma: a randomized clinical trial. Jama 318, 2306–2316 (2017).

53. Goetz, L., Seedat, N., Vandersluis, R. & van der Schaar, M. Generalization—a key challenge for responsible AI in patient-facing clinical applications. NPJ Digital Medicine 7, 126 (2024).

54. Xian, R. P. et al. Robustness tests for biomedical foundation models should tailor to specifications. npj Digital Medicine 8, 557 (2025).

55. Noori, A. et al. A global log for medical AI. 2510.04033 (2025).

56. Liu, S. et al. Isocitrate dehydrogenase (IDH) status prediction in histopathology images of gliomas using deep learning. Scientific reports 10, 7733 (2020).

57. Ciecierski-Holmes, T., Singh, R., Axt, M., Brenner, S. & Barteit, S. Artificial intelligence for strengthening healthcare systems in low-and middle-income countries: a systematic scoping review. NPJ digital medicine 5, 162 (2022).

58. Ilse, M., Tomczak, J. & Welling, M. Attention-based deep multiple instance learning. In International conference on machine learning 2127–2136 (2018).

59. Karthik, M. & Dane, S. APTOS 2019 Blindness Detection https://kaggle.com/competitions/aptos2019-blindness-detection. 2019.

60. Ahn, J. M., Kim, S., Ahn, K.-S., Cho, S.-H., Lee, K. B. & Kim, U. S. A deep learning model for the detection of both advanced and early glaucoma using fundus photography. PloS one 13, e0207982 (2018).

61. Lu, M. Y., Williamson, D. F., Chen, T. Y., Chen, R. J., Barbieri, M. & Mahmood, F. Data-efficient and weakly supervised computational pathology on whole-slide images. Nature biomedical engineering 5, 555–570 (2021).

62. Jin, Y. & Candès, E. J. Model-free selective inference under covariate shift via weighted conformal p-values. arXiv preprint 2307.09291 (2023).

63. Benjamini, Y. & Hochberg, Y. Controlling the false discovery rate: a practical and powerful approach to multiple testing. Journal of the Royal statistical society: series B (Methodological) 57, 289–300 (1995).

64. Hoeffding, W. Probability inequalities for sums of bounded random variables. Journal of the American statistical association 58, 13–30 (1963).

65. Zakharova, G. et al. Reclassification of TCGA diffuse glioma profiles linked to transcriptomic, epigenetic, genomic and clinical data, according to the 2021 WHO CNS tumor classification. International journal of molecular sciences 24, 157 (2022).

66. Cox, D. R. Regression models and life-tables. Journal of the Royal Statistical Society: Series B (Methodological) 34, 187–202 (1972).

