## Supplementary Methods, Supplementary Figure 1, and Supplementary Tables 1 to 13. for "Error-Controlled Decision Policies for Medical Foundation Models"

This PDF file includes:

Supplementary Methods

Supplementary Figure 1

Supplementary Tables 1 to 13

### Supplementary Methods

#### Predictor training and model-selection details

##### Neuro-oncology classification predictors

Across the neuro-oncology classification tasks, UNI [1] patch embeddings were aggregated using attention-based multiple instance learning (ABMIL), and task-specific classification heads were optimized with cross-entropy loss. Hyperparameters were selected by random search, and both model selection and early stopping were based on validation loss. The checkpoint with the lowest validation loss was retained. The search space comprised learning rate $\{1 \times 10^{-4}, 2 \times 10^{-4}, 1 \times 10^{-3}\}$, weight decay $\{1 \times 10^{-5}, 1 \times 10^{-4}, 1 \times 10^{-3}\}$, optimizer {`adam`, `sgd`}, maximum epochs $\{50, 100, 200\}$, dropout $\{0, 0.25, 0.5\}$, and early-stopping patience $\{5, 10\}$. IDH mutation status was optimized as a binary classification task, whereas WHO-based CNS tumor subtyping and the three-class H&E-only diagnostic branches used multiclass cross-entropy. The two-class H&E-only branch used binary cross-entropy.

##### Time-to-mortality predictor

For the time-to-mortality model, the ABMIL-aggregated UNI representation was concatenated with age and biological sex before the survival head. The log-normal accelerated failure time (AFT) head returned patient-specific location and scale parameters $(\hat{\mu}(X), \hat{\sigma}(X))$. Hyperparameters were selected by random search, and both model selection and early stopping were based on validation negative log-likelihood. The checkpoint with the lowest validation loss was retained. The search space comprised learning rate $\{1\times10^{-4}, 2\times10^{-4}, 1\times10^{-3}\}$, weight decay $\{1\times10^{-5}, 1\times10^{-4}, 1\times 10^{-3}\}$, optimizer {`adam`, `sgd`}, maximum epochs $\{20, 30, 50\}$, early-stopping patience $\{5, 10\}$, and number of MLP layers in the survival head $\{1, 2\}$.

##### EHR prediction heads

The released CLMBR-T-base backbone [2] was kept fixed, and task-specific logistic-regression heads were fitted to the frozen 768-dimensional representations using the predefined EHRSHOT training partition, with model selection on the predefined validation partition. The predefined EHRSHOT test partition was not used for head fitting or hyperparameter selection and was subsequently repartitioned for post-hoc StratCP calibration and evaluation as described in Online Methods.

#### Classification baselines and additional implementation details

##### Adaptive prediction-set score

For a classification input $x$, let $\pi_x(1), \ldots, \pi_x(K)$ order the disease statuses by decreasing predicted probability,

$$f(x, y_{\pi_x(1)}) \geq f(x, y_{\pi_x(2)}) \geq \cdots \geq f(x, y_{\pi_x(K)}).$$

We used the non-randomized adaptive prediction-set (APS) conformity score

$$V(x, y_{\pi_x(k)}) = \sum_{\ell=1}^{k} f(x, y_{\pi_x(\ell)}).$$

For standard conformal prediction, the calibration threshold was

$$\hat{\eta} = \text{Quantile}\big(1-\alpha; \{V(X_i, Y_i)\}_{i=1}^{n} \cup \{+\infty\}\big),$$

and the prediction set for test input $X_{n+j}$ was

$$\widehat{C}^{\text{CP}}_{n+j} = \{y \in \mathcal{Y} : V(X_{n+j}, y) \le \hat{\eta}\}.$$

A standard-CP prediction was treated as selected in the action-arm comparison when the resulting prediction set was a singleton. Otherwise the patient or slide was treated as deferred.

**Top-1 and probability-threshold baselines**

The `Top-1` baseline returned the highest-probability disease status for every test patient or slide,

$$\widehat{Y}_{n+j} = \arg\max_{y \in \mathcal{Y}} f(X_{n+j}, y),$$

and therefore acted on every prediction.

For the `Thresh` baseline, disease statuses were ordered by decreasing model probability. Let $\pi_x(1), \ldots, \pi_x(K)$ denote this ordering and define

$$k^\star(x) = \min\left\{k : \sum_{\ell=1}^{k} f(x, y_{\pi_x(\ell)}) \ge 1-\alpha\right\}.$$

The prediction set was

$$\widehat{C}^{\text{Thresh}}(x) = \{y_{\pi_x(1)}, \ldots, y_{\pi_x(k^\star(x))}\}.$$

Thus, the disease status whose inclusion first caused the cumulative probability to reach or exceed $1-\alpha$ was included in the prediction set. A prediction was treated as selected only when this set was a singleton; non-singleton sets were treated as deferred.

**Learn-then-test baseline**

The learn-then-test (`LTT`) baseline [3] calibrates a single global confidence threshold by testing whether the error rate among selected predictions exceeds the target level $\alpha$. For calibration patient $i$, let

$$\widehat{Y}_i = \arg\max_{y \in \mathcal{Y}} f(X_i, y), \qquad s_i = \max_{y \in \mathcal{Y}} f(X_i, y)$$

denote the top-1 predicted disease status and its predicted probability. For a candidate global threshold $\lambda$, define

$$N_\lambda = \sum_{i=1}^{n} \mathbf{1}\{s_i \ge \lambda\}, \qquad E_\lambda = \sum_{i=1}^{n} \mathbf{1}\{s_i \ge \lambda,\ Y_i \ne \widehat{Y}_i\},$$

the number of selected calibration predictions and the number of errors among them.

Let

$$R_\lambda = \Pr\Big(Y \neq \widehat{Y} \mid s(X) \geq \lambda\Big)$$

denote the population error rate under threshold $\lambda$. For each candidate threshold, `LTT` tests

$$H_\lambda : R_\lambda > \alpha.$$

At the boundary $R_\lambda = \alpha$, the number of errors among the $N_\lambda$ selected calibration predictions follows a Binomial$(N_\lambda, \alpha)$ distribution. We therefore compute the lower-tail $p$-value

$$p_\lambda = \Pr\{B_\lambda \leq E_\lambda\}, \qquad B_\lambda \sim \text{Binomial}(N_\lambda, \alpha).$$

A small $p_\lambda$ indicates that observing as few as $E_\lambda$ errors would be unlikely if the selected error rate were at least $\alpha$, providing evidence against $H_\lambda$ and in favor of $R_\lambda \leq \alpha$.

Candidate thresholds were evaluated using the fixed-sequence procedure of [3] at family-wise error level $\delta = 0.01$. A candidate threshold was certified when

$$p_\lambda < \delta,$$

and we retained the smallest, hence most permissive, certified threshold $\widehat{\lambda}$. Candidate thresholds were taken from 100 equally spaced values on $[0, 1]$. If no threshold was certified, no predictions were selected.

A single threshold was calibrated globally using all calibration predictions, rather than separately by predicted disease status. The resulting rule

$$\mathcal{S}^{\text{LTT}} = \Big\{j : s_{n+j} \geq \widehat{\lambda}\Big\}$$

was applied to all test predictions. For disease-status-specific reporting, selected patients or slides were subsequently stratified by their top-1 predicted disease status; no disease-status-specific `LTT` thresholds were fitted.

**Hoeffding baseline**

The `Hoeffding` baseline [3, 4] used the same top-1 confidence scores, candidate global thresholds, and target error level $\alpha$ as `LTT`, but certified thresholds using a simultaneous uniform concentration bound rather than binomial hypothesis tests.

For candidate threshold $\lambda$, let $N_\lambda$ and $E_\lambda$ denote the number of selected calibration predictions and the number of errors among them, as defined above. We used the transformed empirical risk

$$\widehat{\mu}_\lambda = \alpha + \frac{E_\lambda - \alpha N_\lambda}{n}.$$

If

$$q_\lambda = \Pr\{s(X) \geq \lambda\}$$

is the population selection probability and $R_\lambda$ is the corresponding selected error rate, the population analogue is

$$\mu_\lambda = \alpha + q_\lambda(R_\lambda - \alpha).$$

For $q_\lambda > 0$,

$$\mu_\lambda < \alpha \quad \Longleftrightarrow \quad R_\lambda < \alpha,$$

so certification can equivalently be performed on $\mu_\lambda$.

Following the uniform-concentration procedure in [3], the auxiliary parameter $\eta$ and concentration radius $t$ were determined numerically. The implementation searches over a fixed logarithmic grid of candidate $\eta$ values and, for each candidate, numerically determines the concentration radius required to attain the specified tail probability. After selecting $\eta^\star$, the final concentration radius $t^\star$ was recomputed at the prespecified failure probability $\delta = 0.01$.

The simultaneous upper confidence bound for each candidate threshold was

$$U_\lambda = \widehat{\mu}_\lambda + t^\star\sqrt{\widehat{\mu}_\lambda + \eta^\star + \frac{(t^\star)^2}{4}} + \frac{(t^\star)^2}{2}.$$

A threshold was certified when

$$U_\lambda < \alpha,$$

and the smallest certified threshold $\widehat{\lambda}$ was retained. A single threshold was calibrated globally across all disease statuses and applied to all test predictions. Selected patients or slides were subsequently stratified by their top-1 predicted disease status for reporting; no disease-status-specific `Hoeffding` thresholds were fitted. If no threshold was certified, no predictions were selected.

**Deferred-set and end-to-end metrics**

For any method that produces an action set $\mathcal{S}$ and prediction sets for deferred patients or slides, deferred-set coverage was computed as

$$\widehat{\mathrm{Cov}}_{\mathrm{def}} = \frac{\sum_{j \notin \mathcal{S}} \mathbf{1}\{Y_{n+j} \in \widehat{C}_{n+j}\}}{\max\{1, |\mathcal{S}^c|\}},$$

and mean deferred prediction-set size as

$$\widehat{\mathrm{Size}}_{\mathrm{def}} = \frac{\sum_{j \notin \mathcal{S}} |\widehat{C}_{n+j}|}{\max\{1, |\mathcal{S}^c|\}}.$$

For end-to-end marginal evaluation, selected predictions were represented by singleton sets containing the top-1 prediction and deferred patients or slides by their calibrated prediction sets. Marginal coverage and mean prediction-set size were then computed over the full evaluation cohort. When a method yielded no deferred observations for a mathematically undefined metric, that replicate was excluded from aggregation for that metric rather than assigned an arbitrary value.

##### Efficient computation of selection-conditional prediction sets

The direct implementation of the selection-conditional calibration procedure in Online Methods requires reconstructing a test-specific reference set for each deferred test observation and candidate disease status. We used computational shortcuts that reuse action-arm threshold calculations rather than rerunning the full selection procedure from scratch for every swap.

**Classification shortcut.** For multiclass classification, the implementation maintains, for each candidate disease status $y$, an indicator matrix identifying which calibration observations belong to the reference set of each deferred test observation. For each deferred test observation $j$ and action-arm criterion $k$, the algorithm evaluates the counterfactual action-arm threshold under the swap configurations induced by the candidate disease status assigned to $j$ and by the error status of the calibration observation moved to the test set. Calibration observations for which the swapped data would cause $j$ to be selected are removed from the corresponding reference set. The final prediction set is then calibrated using only the retained reference observations.

**Algorithm 3:** Computation shortcut of Algorithm 2 for classification

**Input :** Foundation model $f$; confident prediction selection rule $\mathcal{A}_k$ (Algorithm 1); conformity score $V(x, y)$; calibration data $\{(X_i, Y_i)\}_{i=1}^n$; test inputs $\{X_{n+j}\}_{j=1}^m$; already-selected test set $\mathcal{S}_k$; trustworthiness criteria $\{\mathcal{I}_k(\cdot)\}$; eligibility criteria $\{\mathcal{G}_k(\cdot)\}$; disease status space $\mathcal{Y}$; coverage level $1 - \alpha$.

Initialize matrices $R^{(y)} \leftarrow \mathbf{1}_{m \times n}$ for all $y \in \mathcal{Y}$.
Set $R^{(y)}[j, :] \leftarrow 0$ for all $j \in \bigcup_k \mathcal{S}_k$, $y \in \mathcal{Y}$.
**for** $j \in [m] \setminus \left(\bigcup_k \mathcal{S}_k\right)$ **do**
  // Construct reference sets $\mathcal{D}_{n+j}(y)$ for deferred patients
  **for** $k = 1, \ldots, K$ **do**
    Compute sample sizes $n_k = |G_k^{\text{calib}}|$, $m_k = |G_k^{\text{test}}|$.
    Compute error indicators $L_i = 1 - \mathcal{I}_k(f, X_i, Y_i)$ for $i \in G_k^{\text{calib}}$.
    **if** $\mathcal{G}_k(f, X_{n+j}) = 1$ **then**
      Compute thresholds $\tau_{ab} = \min\{t : R_{ab}(t) \leq \alpha\}$, for $a, b \in \{0, 1\}$, where

$$R_{ab}(t) = \frac{a + \sum_{i \in G_k^{\text{calib}}} L_i \mathbf{1}\{s_k(X_i) \geq t\} + b\mathbf{1}\{s_k(X_{n+j}) \geq t\}}{n_k + 1} \cdot \frac{m_k}{\sum_{\ell \in G_k^{\text{test}} \setminus \{j\}} \mathbf{1}\{s_k(X_{n+\ell}) \geq t\}}.$$

      **for** $y \in \mathcal{Y}$ **do**
        If $\mathcal{I}(f, X_{n+j}, y) = 1$, set $R^{(y)}[j, i] = 0$ for $i \in [n]$ with

$$(E_i^k = 1, L_i^k = 1, s_k(X_i) \geq \tau_{10}) \text{ or } (E_i^k = 1, L_i^k = 0, s_k(X_i) \geq \tau_{00}).$$

        If $\mathcal{I}(f, X_{n+j}, y) = 0$, set $R^{(y)}[j, i] = 0$ for $i \in [n]$ with

$$(E_i^k = 1, L_i^k = 1, s_k(X_i) \geq \tau_{11}) \text{ or } (E_i^k = 1, L_i^k = 0, s_k(X_i) \geq \tau_{01}).$$

    **else**
      Compute thresholds $\tau_a = \min\{t : R_a(t) \leq \alpha\}$ for $a \in \{0, 1\}$, where

$$R_a(t) = \frac{a + \sum_{i \in G_k^{\text{calib}}} L_i \mathbf{1}\{s_k(X_i) \geq t\}}{n_k + 1} \cdot \frac{m_k - 1}{\sum_{\ell \in G_k^{\text{test}}} \mathbf{1}\{s_k(X_{n+\ell}) \geq t\}}.$$

      **for** $y \in \mathcal{Y}$ **do**
        Set $R^{(y)}[j, i] = 0$ for $i \in G_k^{\text{calib}}$ with

$$(L_i^k = 1, s_k(X_i) \geq \tau_1) \text{ or } (L_i^k = 0, s_k(X_i) \geq \tau_0).$$

  // Construct prediction sets
  Compute

$$\widehat{C}_{n+j} = \left\{y : V(X_{n+j}, y) \leq \text{Quantile}\Big(1 - \alpha; \{V(X_i, Y_i)\}_{R^{(y)}[j,i]=1} \cup \{+\infty\}\Big)\right\}.$$

**Output :** Prediction sets $\widehat{C}_{n+j}$ for all $j \in [m] \setminus (\cup_k \mathcal{S}_k)$.

**Time-to-event shortcut.** For a deferred test patient $j$, the hypothesized survival time affects the counterfactual selection only through whether it falls below or at/above the clinical horizon $c$. We therefore consider two outcome regions, indexed by $b = 1$ for $y < c$ and $b = 0$ for $y \geq c$.

To determine which calibration patients remain valid references for patient $j$ in each region, the shortcut also accounts for whether the observed follow-up time of the calibration patient moved into the test set lies below the clinical horizon, $L_i = \mathbf{1}\{Y_i < c\}$. These two binary quantities give four patient-specific counterfactual selection thresholds $\tau_{ab}^{(j)}$, $a, b \in \{0, 1\}$, defined in Algorithm 4. For each outcome region $b$, the appropriate threshold is chosen according to $L_i$ to form the reference set $D_{j,b}$. Thus, the four counterfactual thresholds yield two patient-specific reference sets, $D_{j,1}$ for $y < c$ and $D_{j,0}$ for $y \geq c$. Each reference set determines a conformity-score cutoff $\eta_{j,b}$, and the accepted survival times from the two regions are combined into a raw prediction set. We finally report the smallest one-sided interval containing this set, $[\widehat{L}_j, \infty)$.

**Algorithm 4:** Computation shortcut of Algorithm 2 for time-to-event analysis

**Input :** Foundation model $f$ (estimates survival CDF $\widehat{F}(t \mid x)$); estimated censoring survival function $\widehat{G}(t \mid x)$; confident prediction selection procedure $\mathcal{A}$ (Algorithm 1); conformity score $V(x, y) = \alpha - \widehat{F}(y \mid x)$; calibration data $\{(X_i, Y_i, \Delta_i)\}_{i=1}^{n}$ with $Y_i = \min(T_i, C_i)$; test inputs $\{X_{n+j}\}_{j=1}^{m}$; already-selected test set $\mathcal{S}$; trustworthiness criterion $\mathcal{I}(x, y) = \mathbf{1}\{y \geq c\}$ for some survival time $c > 0$; coverage level $1 - \alpha$.

**for** $j \in [m] \setminus \mathcal{S}$ **do**

- // Construct reference sets $\mathcal{D}_{n+j}(y)$ for deferred patients
- // IPCW error surrogates at the clinical horizon $c$
- Define $\widetilde{G}(t \mid x) = \max\{\widehat{G}(t \mid x), 0.05\}$ and compute $Z_i(c) = \mathbf{1}\{Y_i < c, \Delta_i = 1\}/\widetilde{G}(Y_i \mid X_i)$ for $i \in [n]$.
- // Favorable-early-survival score at horizon $c$ (smaller $\Rightarrow$ longer survival). For log-normal AFT use a standardized log-time; for other survival heads, any score monotonically increasing in $\widehat{F}(c \mid x)$ is valid.
- Define $s(x; c) = \dfrac{\log c - \widehat{\mu}(x)}{\widehat{\sigma}(x)}$ // log-normal AFT case
- Set $s_i = s(X_i; c)$ and $L_i = \mathbf{1}\{Y_i < c\}$ for $i \in [n]$, and $u_\ell = s(X_{n+\ell}; c)$ for $\ell \in [m]$.
- Define $\mathcal{T} = \{s_i : i \in [n]\} \cup \{u_\ell : \ell \in [m]\}$ and $A(t) = \sum_{i=1}^{n} Z_i(c)\mathbf{1}\{s_i \leq t\}$ for $t \in \mathcal{T}$.
- // Selection thresholds with test patient $j$ posited as calibration
- Compute $B_{-j}(t) = \sum_{\ell \in [m]\setminus\{j\}} \mathbf{1}\{u_\ell \leq t\}$ for $t \in \mathcal{T}$.
- // $b$ indexes the hypothesized outcome of patient $j$, with $b = 1$ for $y < c$ and $b = 0$ for $y \geq c$; for calibration observation $i$, $a = 1 - L_i$.
- Compute four thresholds $\tau_{ab}^{(j)} = \max\{t \in \mathcal{T} : R_{ab}^{(j)}(t) \leq \alpha\}$, $a, b \in \{0, 1\}$, with $\max \varnothing = -\infty$, where

$$R_{ab}^{(j)}(t) = \frac{a + A(t) + b\mathbf{1}\{u_j \leq t\}}{n + 1} \cdot \frac{m}{1 + B_{-j}(t)}.$$

- **for** $b \in \{0, 1\}$ **do**
  - // Construct the deferral reference set
  - Compute $D_{j,b} = \{i \in [n] : L_i = 1,\ s_i > \tau_{0b}^{(j)}\} \cup \{i \in [n] : L_i = 0,\ s_i > \tau_{1b}^{(j)}\}$.
  - Compute $\eta_{j,b} = \mathrm{Quantile}\Big(1 - \alpha; \{V(X_i, Y_i)\}_{i \in D_{j,b}} \cup \{+\infty\}\Big)$.
- // Construct the selection-conditional prediction set
- Compute $\widehat{C}_{n+j}^{\mathrm{raw}} = \{y \in (0, c) : V(X_{n+j}, y) \leq \eta_{j,1}\} \cup \{y \in [c, \infty) : V(X_{n+j}, y) \leq \eta_{j,0}\}$.
- // Convert to the reported one-sided lower prediction interval
- Set $\widehat{L}_j = \inf \widehat{C}_{n+j}^{\mathrm{raw}}$ and $\widehat{C}_{n+j}^{\mathrm{LPB}} = [\widehat{L}_j, \infty)$, with $\inf \varnothing = +\infty$.

**Output :** $\widehat{C}_{n+j}^{\mathrm{LPB}}$ for all $j \in [m] \setminus \mathcal{S}$.

##### Utility-enhanced prediction sets

##### Utility-informed label ordering

Let $\mathcal{U} = (\mathcal{Y}, E)$ denote a task-specific utility graph with edge weight $E_{y,y'}$ encoding the clinical relatedness of disease statuses $y$ and $y'$. Utility enhancement modifies only the order in which disease status enter the cumulative-probability conformity score. The same selection-conditional calibration procedure is then applied to the resulting scores.

For an input $x$, the first ranked disease status is the top-1 prediction,

$$\pi(1) = \arg\max_{k \in [K]} f(x, y_k).$$

Let $\mathcal{L} = [K] \setminus \{\pi(1)\}$ be the unranked disease statuses. At each subsequent step, candidate disease statuses are restricted to the $N$ highest-probability disease statuses remaining in $\mathcal{L}$, where $N$ is a prespecified search radius. Among these candidates, the next disease status is chosen to maximize its strongest utility edge to a previously ranked disease status,

$$\pi(\ell + 1) \in \arg\max_{k \in C_\ell} \max_{r \le \ell} E_{y_{\pi(r)}, y_k}.$$

After obtaining $\pi(1), \ldots, \pi(K)$, the utility-enhanced conformity score is

$$V_{\text{util}}(x, y_{\pi(k)}) = \sum_{\ell=1}^{k} f(x, y_{\pi(\ell)}).$$

Thus, utility enhancement is a utility-informed ordering heuristic constrained by model probability. It does not imply global optimization of an average pairwise set-utility objective. The resulting conformity scores are supplied to the same selection-conditional calibration procedure used for the standard APS score. In the greedy implementation, ties were resolved implicitly by NumPy ordering: candidate labels were ordered by descending model probability using reverse `np.argsort`, and `np.argmax` selected the first candidate attaining the maximal utility. No additional clinically defined tie-breaking rule was imposed.

##### Diabetic retinopathy utility

For diabetic retinopathy, the five severity stages were treated as an ordered chain. We set $E_{y,y'} = 1$ for immediately adjacent disease stages and $E_{y,y'} = 0$ otherwise and used search radius $N = K = 5$. Clinical coherence was evaluated by whether all disease statuses in a deferred prediction set formed a contiguous interval on the five-stage severity ordering. Deferred-set coverage was evaluated in parallel to confirm that utility enhancement did not compromise empirical calibration.

##### Eye-condition management-action utility

For the 39-class eye-condition task, utility was defined by overlap in downstream management actions. Using the condition descriptions supplied with the dataset, the GPT-o1 API was queried to generate ($M = 10$) candidate management actions for each disease status. Semantically synonymous actions were manually merged across conditions, yielding 89 distinct actions. Let $(A_y)$

denote the resulting set of management actions for disease status ($y$). Pairwise utility was defined as

$$E_{y,y'} = |A_y \cap A_{y'}|.$$

such that disease statuses requiring more similar downstream management had higher utility. Supplementary Fig. S1 illustrates this action-overlap structure and its effect on the composition of a deferred prediction set.

The utility-informed conformity score was used for selection-conditional calibration of deferred prediction sets. The greedy utility algorithm used the default candidate window (N=3), such that each expansion step considered the three highest-probability disease statuses not yet ranked. Management-action coherence was compared between standard and utility-enhanced StratCP prediction sets after stratification by prediction-set size.

**WHO-grade utility for CNS tumor subtyping**

For WHO-based CNS tumor subtyping, we defined a subtype-level WHO grade mapping using the annotations summarized in Supplementary Table S7. A subtype was assigned a numeric WHO grade only when its available nonmissing grade annotations agreed on a single grade (I–IV). Subtypes with no numeric grade or with conflicting grade annotations were treated as ungraded. In particular, Haemangiopericytoma was treated as ungraded because specimens with this reference diagnosis spanned WHO grades I–III in the source metadata.

Let $g(y) \in \{1,2,3,4\} \cup \{\varnothing\}$ denote the resulting subtype-level grade assignment. The utility graph was defined as

$$E_{y,y'} = \begin{cases} 1, & y = y', \\ 1, & y \neq y',\ g(y) = g(y') \in \{1,2,3,4\}, \\ 0, & \text{otherwise.} \end{cases}$$

Thus, subtypes sharing the same numeric WHO grade were connected, whereas subtypes from different grades and distinct ungraded subtypes were not connected. Ungraded subtypes retained only their self-relationship. The greedy utility algorithm used the default candidate window $N = 3$.

Grade coherence was evaluated only for prediction sets in which every included subtype had an assigned numeric WHO grade. Prediction sets containing one or more ungraded subtypes were excluded from both the numerator and denominator of this metric; their original prediction-set size was retained for size-bin assignment. Among eligible sets, a prediction set was considered grade coherent if it contained the top-1 predicted subtype and

$$\max_{y \in \widehat{C}} g(y) - \min_{y \in \widehat{C}} g(y) < 2.$$

Accordingly, the coherence metric regarded subtypes from the same or adjacent WHO grades as coherent, whereas the construction graph connected only subtypes sharing the same numeric WHO grade.

**Thrombocytopenia utility**

For thrombocytopenia, the four disease statuses were ordered as normal, mild, moderate, and severe. We defined a chain utility graph with $E_{y,y'} = 1$ for the adjacent pairs normal–mild, mild–moderate, and moderate–severe, and $E_{y,y'} = 0$ otherwise. For a prediction set $\widehat{C}$, let $I(y) \in \{1, 2, 3, 4\}$ denote the ordinal severity index. A prediction set was defined as consecutive when

$$\max_{y \in \widehat{C}} I(y) - \min_{y \in \widehat{C}} I(y) + 1 = |\widehat{C}|.$$

The greedy utility algorithm used the default candidate window $N = 3$. The fraction of consecutive deferred prediction sets was reported separately by prediction-set size. Prediction sets of size four were omitted from this visualization because they necessarily contain all four disease statuses and are consecutive by definition. Deferred-set coverage was evaluated in parallel across target error levels.

**Cross-cohort IDH-status visualization**

For the cohort visualization in the cross-cohort IDH-status analysis, the fixed ABMIL predictor produced a 512-dimensional representation for each slide. Slide representations were averaged within each patient to obtain a patient-level representation. Features were standardized using `StandardScaler`, followed by principal-component analysis with

$$n_{\text{components}} = \min\{50, n_{\text{samples}}, n_{\text{features}}\},$$

which yielded 50 principal components for the analyzed cohort. UMAP was then applied with 30 neighbors, minimum distance 0.1, and cosine distance. The analysis environment used `scikit-learn` v1.7.2, and `umap-learn` v0.5.12.

**H&E-only diagnostic triage in adult-type diffuse glioma**

**Diagnostic-label harmonization**

TCGA-LGG and TCGA-GBM cases were reclassified using available molecular markers following the molecular-data-based mapping described by [5] and subsequently mapped to WHO 2016 terminology to align with the EBRAINS labels [6, 7]. The full harmonized disease-status set and slide counts are reported in Supplementary Table S11. Cases with NOS diagnoses or otherwise unavailable labels required for the branch-specific endpoint were excluded. Here, 1p/19q codeletion denotes combined whole-arm loss of 1p and 19q.

**Morphology-defined branches**

Diagnostic H&E slides were stratified according to the presence of microvascular proliferation (MVP) or necrosis, mitotic activity in the absence of MVP/necrosis, or neither feature. Separate UNI–ABMIL classifiers were trained within each branch using the disease statuses represented in Supplementary Table S11:

(i) **MVP or necrosis present:** anaplastic oligodendroglioma, IDH-mutant and 1p/19q-codeleted; glioblastoma, IDH-mutant; and glioblastoma, IDH-wildtype.

(ii) **Mitotic activity without MVP/necrosis:** anaplastic astrocytoma, IDH-mutant; anaplastic astrocytoma, IDH-wildtype; and anaplastic oligodendroglioma, IDH-mutant and 1p/19q-codeleted.

(iii) **Neither MVP/necrosis nor mitotic activity:** diffuse astrocytoma, IDH-mutant; and oligodendroglioma, IDH-mutant and 1p/19q-codeleted.

Only two disease statuses were modeled in the third branch because the analyzed cohort contained no diffuse astrocytoma, IDH-wildtype slides satisfying that branch definition. NOS diagnoses and cases lacking the labels required for the branch-specific endpoint were excluded.

**Branch-specific training and evaluation**

Each branch-specific classifier used UNI-derived patch embeddings with an ABMIL prediction head and the classification hyperparameter search described above. Data were split at the patient level into 45/20/20/15% train/validation/conformal-calibration/test sets. StratCP selection was calibrated separately for each predicted disease status at $\alpha = 0.05$. The analysis evaluated only the action arm because its purpose was to identify diagnoses that could be resolved from H&E morphology alone; deferred prediction sets were not constructed.

#### Additional methods for right-censored survival prediction

##### Notation and data structure

For each patient, let $T > 0$ denote the true time from diagnosis to mortality and $C > 0$ the censoring time. We observe

$$Y = \min(T, C), \qquad \Delta = \mathbf{1}\{T \le C\},$$

where $\Delta = 1$ denotes an observed death and $\Delta = 0$ denotes right censoring. Administrative censoring was applied at $t_{\max} = 5$ years. Let $X$ denote the patient-level predictors used by the survival model, comprising the ABMIL-aggregated UNI representation together with age and biological sex.

For a given evaluation, the conformal-calibration data are

$$\mathcal{D}_{\text{calib}} = \{(X_i, Y_i, \Delta_i)\}_{i=1}^{n},$$

and the held-out test data are

$$\mathcal{D}_{\text{test}} = \{(X_{n+j}, Y_{n+j}, \Delta_{n+j})\}_{j=1}^{m}.$$

Only the test covariates $X_{n+j}$ are used when producing action or deferral decisions; $(Y_{n+j}, \Delta_{n+j})$ are reserved for evaluation.

We fixed the favorable-early-survival horizon at

$$c = 18 \text{ months}$$

and used $\alpha \in (0, 1)$ to denote the target action-arm error level and deferral-arm miscoverage level; the primary analysis used $\alpha = 0.05$.

**Log-normal AFT model**

Conditional on patient covariates $X = x$, the log-normal accelerated failure time (AFT) model assumes

$$\log T \mid X = x \sim \mathcal{N}\Big(\mu_\theta(x), \sigma_\theta^2(x)\Big),$$

where $\theta$ denotes the trainable model parameters. After fitting, we denote the patient-specific location and scale estimates by $\widehat{\mu}(x)$ and $\widehat{\sigma}(x)$.

The fitted conditional cumulative distribution function (CDF) and survival function are

$$\widehat{F}(t \mid x) = \Phi\left(\frac{\log t - \widehat{\mu}(x)}{\widehat{\sigma}(x)}\right), \qquad \widehat{S}(t \mid x) = 1 - \widehat{F}(t \mid x),$$

where $\Phi$ denotes the standard normal CDF. The corresponding log-normal density is

$$f_{\log N}(t \mid x) = \frac{1}{t\,\widehat{\sigma}(x)\sqrt{2\pi}} \exp\left[-\frac{(\log t - \widehat{\mu}(x))^2}{2\widehat{\sigma}^2(x)}\right], \qquad t > 0.$$

The survival model was fitted on the training data by maximizing the right-censoring-aware log-likelihood

$$\ell(\theta) = \sum_{r \in \mathcal{D}_{\text{train}}} \left\{\Delta_r \log f_{\log N}(Y_r \mid X_r; \theta) + (1 - \Delta_r) \log S_{\log N}(Y_r \mid X_r; \theta)\right\}.$$

Thus, an observed death contributes the log density at its observed survival time, whereas a right-censored observation contributes the log probability of surviving beyond its last observed follow-up time.

**Censoring model and inverse probability of censoring weighting**

Let

$$G(t \mid x) = \Pr(C \geq t \mid X = x)$$

denote the true conditional censoring survival function. We estimated $G$ using a Cox proportional hazards model fitted to the censoring process, treating $1 - \Delta$ as the censoring-event indicator.

Within the conformal-calibration cohort, the censoring model was cross-fitted so that the censoring probability assigned to each calibration patient was obtained from a model that did not use that patient for fitting. The censoring model was subsequently refitted on the complete conformal-calibration cohort for evaluation of test patients. Let $\widehat{G}(t \mid x)$ denote the resulting estimated censoring survival probability. For numerical stability, the probability used for weighting was

$$\widetilde{G}(t \mid x) = \max\{\widehat{G}(t \mid x), 0.05\}.$$

At the clinical horizon $c$, the IPCW surrogate for the action-arm error event $\{T_i < c\}$ was

$$Z_i(c) = \frac{\mathbf{1}\{Y_i < c, \Delta_i = 1\}}{\widetilde{G}(Y_i \mid X_i)}, \qquad i = 1, \ldots, n.$$

The numerator records an observed death before $c$, whereas inverse weighting accounts for comparable deaths that may be unobserved because of censoring.

With the true $G$ and under conditional independent censoring,

$$T \perp C \mid X,$$

the corresponding IPCW quantity targets the probability of death before $c$. In practice, $G$ is replaced by the estimated and clipped $\widetilde{G}$, and censoring-adjusted action- and deferral-arm properties were therefore evaluated empirically. The analysis additionally assumes positivity over the evaluated time range, so that $G(t \mid X) > 0$ wherever IPCW weights are required.

**Action-arm calibration under right censoring**

For patient covariates $x$, define the favorable-early-survival score

$$s(x; c) = \frac{\log c - \widehat{\mu}(x)}{\widehat{\sigma}(x)}.$$

Because

$$\widehat{F}(c \mid x) = \Phi\big(s(x; c)\big),$$

smaller values of $s(x; c)$ correspond to a lower model-implied probability of death before $c$ and therefore a higher probability of surviving at least to $c$.

For a candidate threshold $\tau$, calibration patients with $s(X_i; c) \leq \tau$ satisfy the corresponding favorable-survival action rule. For intuition, the calibration-only censoring-adjusted plug-in estimate of the selected error rate is

$$\widehat{R}(\tau) = \frac{\sum_{i=1}^{n} Z_i(c)\mathbf{1}\{s(X_i; c) \leq \tau\}}{\max\left\{1, \sum_{i=1}^{n} \mathbf{1}\{s(X_i; c) \leq \tau\}\right\}}.$$

Thus, $\widehat{R}(\tau)$ estimates the censoring-adjusted probability of death before $c$ among calibration patients satisfying the candidate action rule.

The action-arm threshold was determined using a finite-sample-adjusted selection statistic that additionally accounts for how many test patients would be selected at each candidate threshold. Let

$$\mathcal{T} = \{s(X_i; c) : i \in [n]\} \cup \{s(X_{n+j}; c) : j \in [m]\}$$

denote the pooled set of calibration and test scores. For each candidate threshold $\tau \in \mathcal{T}$, define

$$\widehat{R}_{\text{sel}}(\tau) = \frac{1 + \sum_{i=1}^{n} Z_i(c)\mathbf{1}\{s(X_i; c) \leq \tau\}}{\max\left\{1, \sum_{j=1}^{m} \mathbf{1}\{s(X_{n+j}; c) \leq \tau\}\right\}} \frac{m}{n+1}.$$

The numerator is the cumulative IPCW calibration-error contribution, with the finite-sample correction of 1, and the denominator is the number of test patients satisfying the candidate action rule. The factor $m/(n + 1)$ accounts for the relative test and calibration sample sizes.

We then choose the largest admissible threshold

$$\widehat{\tau} = \max \left\{ \tau \in \mathcal{T} : \widehat{R}_{\text{sel}}(\tau) \leq \alpha \right\},$$

with $\widehat{\tau} = -\infty$ if no candidate threshold satisfies the target error level. In the implementation, the cumulative statistic is evaluated along the pooled scores ordered in increasing order; tied scores are not explicitly grouped.

The resulting test action arm is

$$\mathcal{S}(\alpha) = \left\{ j \in \{1, \ldots, m\} : s(X_{n+j}; c) \leq \widehat{\tau} \right\},$$

and its complement

$$\mathcal{D}(\alpha) = \{1, \ldots, m\} \setminus \mathcal{S}(\alpha)$$

is the deferral arm.

**Selection-conditional lower prediction bound.**

For each deferred test patient $j \in \mathcal{D}(\alpha)$, the reference set used for selection-conditional calibration can depend on whether the hypothesized survival time lies below or at/above the clinical horizon. Because

$$I(x, y) = \mathbf{1}\{y \geq c\},$$

we consider two outcome regions,

$$b = 1 \quad \text{for } y < c, \qquad b = 0 \quad \text{for } y \geq c.$$

To determine which calibration patients remain valid references for patient $j$ in each region, Algorithm 4 considers four patient-specific counterfactual selection thresholds

$$\tau_{ab}^{(j)}, \qquad a, b \in \{0, 1\}.$$

Here, $b$ indexes the hypothesized outcome region for patient $j$, while $a = 1 - L_i$ indexes the horizon status of calibration patient $i$, with

$$L_i = \mathbf{1}\{Y_i < c\}.$$

For each outcome region $b$, the appropriate threshold is therefore chosen according to $L_i$, yielding the reference set $D_{j,b}$ in Algorithm 4. We write

$$\mathcal{R}_{j,b} = D_{j,b}, \qquad b \in \{0, 1\},$$

so that $\mathcal{R}_{j,1}$ corresponds to hypothesized survival below $c$ and $\mathcal{R}_{j,0}$ to survival at or above $c$.

For calibration patient $i$, define the fitted CDF value at the observed follow-up time

$$U_i = \widehat{F}(Y_i \mid X_i)$$

and the one-sided conformity score

$$V_i = \alpha - U_i.$$

For each outcome region $b \in \{0, 1\}$, let

$$r_{j,b} = |\mathcal{R}_{j,b}|$$

and define the selection-conditionally calibrated cutoff

$$\eta_{j,b} = \mathrm{Quantile}\big(1 - \alpha; \{V_i : i \in \mathcal{R}_{j,b}\} \cup \{+\infty\}\big).$$

Equivalently, if the $r_{j,b}$ finite conformity scores are ordered as

$$V_{(1)}^{j,b} \le \cdots \le V_{(r_{j,b})}^{j,b},$$

the conformal order-statistic index is

$$k_{j,b} = \lceil (1-\alpha)(r_{j,b} + 1) \rceil,$$

with the usual $+\infty$ convention when the required order statistic exceeds the number of finite reference scores.

Because $V_i = \alpha - U_i$, define the corresponding fitted-CDF probability level as

$$p_{j,b} = \alpha - \eta_{j,b}.$$

Let

$$q_x(p) = \inf\{t > 0 : \widehat{F}(t \mid x) \ge p\}$$

denote the generalized inverse of the fitted survival-time CDF. The lower-bound component associated with outcome region $b$ is

$$L_{j,b} = q_{X_{n+j}}(p_{j,b}), \qquad b \in \{0, 1\}.$$

Because $\widehat{F}(y \mid X_{n+j})$ is nondecreasing in $y$, the accepted survival times from the two outcome regions form the raw selection-conditional prediction set

$$\widehat{C}_{n+j}^{\mathrm{raw}} = \big([L_{j,1}, \infty) \cap (0, c)\big) \cup \big([L_{j,0}, \infty) \cap [c, \infty)\big).$$

For reporting a single one-sided lower prediction interval, we take the smallest closed upper interval containing this set:

$$\widehat{L}_j = \inf \widehat{C}_{n+j}^{\mathrm{raw}}, \qquad \widehat{C}_{n+j}^{\mathrm{LPB}} = [\widehat{L}_j, \infty).$$

For the fitted log-normal AFT model, each branch-specific lower-bound component is

$$L_{j,b} = \exp\left\{\widehat{\mu}(X_{n+j}) + \widehat{\sigma}(X_{n+j})\Phi^{-1}(p_{j,b})\right\}, \qquad b \in \{0, 1\},$$

where $\Phi^{-1}$ denotes the standard normal quantile function.

The JOMI reference-set construction provides the selection-conditional calibration mechanism described in Online Methods. Because survival outcomes are right-censored and the censoring distribution is estimated, coverage of the latent survival time was evaluated empirically using IPCW rather than claimed to satisfy an exact finite-sample guarantee under censoring.

**IPCW evaluation estimators**

For test patient $j$, define the IPCW contribution corresponding to violation of an action-arm favorable-survival declaration as

$$W_j^{\mathrm{act}} = \frac{\mathbf{1}\{Y_{n+j} < c, \Delta_{n+j} = 1\}}{\widetilde{G}(Y_{n+j} \mid X_{n+j})}.$$

For one evaluation, the censoring-adjusted false discovery proportion among patients selected into the action arm is

$$\widehat{\mathrm{FDP}}_{\mathrm{IPCW}} = \frac{\sum_{j \in \mathcal{S}(\alpha)} W_j^{\mathrm{act}}}{\max\{1, |\mathcal{S}(\alpha)|\}}.$$

Across the 500 evaluations, FDR was estimated as the mean of $\widehat{\mathrm{FDP}}_{\mathrm{IPCW}}$, and selected-set coverage was reported as

$$1 - \mathrm{FDR}.$$

For a deferred patient $j \in \mathcal{D}(\alpha)$ with lower prediction bound $\widehat{L}_j$, define

$$W_j^{\mathrm{def}} = \frac{\mathbf{1}\{Y_{n+j} < \widehat{L}_j, \Delta_{n+j} = 1\}}{\widetilde{G}(Y_{n+j} \mid X_{n+j})}.$$

This quantity represents the IPCW contribution of an observed death occurring below the claimed lower survival bound. Deferral-arm coverage was computed as one minus the censoring-adjusted violation rate among deferred patients.

For overall marginal evaluation, define the lower endpoint assigned to test patient $j$ as

$$\theta_j = \begin{cases} c, & j \in \mathcal{S}(\alpha), \\ \widehat{L}_j, & j \in \mathcal{D}(\alpha). \end{cases}$$

Overall marginal coverage was computed from the corresponding IPCW-adjusted rate of violations $T_{n+j} < \theta_j$. Prediction-set efficiency was summarized by the interval length truncated at the administrative study end,

$$\ell_j = \max\{0, t_{\max} - \theta_j\},$$

averaged over the test cohort.

**Survival baselines**

The `Top-1` baseline used the fitted survival probability

$$\widehat{S}(c \mid X) = 1 - \widehat{F}(c \mid X)$$

and assigned favorable early survival when

$$\widehat{S}(c \mid X) \geq 0.5.$$

Standard conformal prediction (CP) constructed a marginal conformal lower prediction bound $\widehat{L}^{\text{CP}}$ for every test patient, without conditioning calibration on the action-arm selection event. A patient was assigned favorable early survival when

$$\widehat{L}^{\text{CP}} \geq c.$$

The log-normal quantile baseline (LNQ) used the fitted AFT distribution directly without conformal calibration. It assigned favorable early survival when

$$\widehat{S}(c \mid X) \geq 1 - \alpha$$

and otherwise returned the parametric lower prediction bound

$$\widehat{L}^{\text{LNQ}} = \exp\left\{\widehat{\mu}(X) + \widehat{\sigma}(X)\Phi^{-1}(\alpha)\right\},$$

yielding the interval

$$[\widehat{L}^{\text{LNQ}}, \infty).$$

**Kaplan–Meier analyses**

Let $\mathcal{S}^{(b)}(0.05)$ denote the action-arm patients selected in evaluation $b$, for $b = 1, \ldots, 500$. For the descriptive Kaplan–Meier analysis, we defined the action-arm group as the unique union

$$\mathcal{S}_{\text{KM}} = \bigcup_{b=1}^{500} \mathcal{S}^{(b)}(0.05),$$

with each patient represented only once. All remaining patients in the pooled evaluation population formed the deferral group,

$$\mathcal{D}_{\text{KM}} = \mathcal{U}_{\text{eval}} \setminus \mathcal{S}_{\text{KM}},$$

where $\mathcal{U}_{\text{eval}}$ denotes the unique set of patients appearing in the evaluation cohorts.

Kaplan–Meier curves were constructed using each patient's single observed pair $(Y, \Delta)$, and survival differences between the two mutually exclusive groups were assessed using a log-rank test. Grade-specific analyses were restricted to WHO CNS5 subtypes following molecular-data-based diagnostic remapping [5]. Grade 4 comprised glioblastoma, IDH-wildtype and astrocytoma, IDH-mutant, grade 4. Kaplan–Meier analyses were implemented using `lifelines` v0.30.0 [8].

**Retrospective DFCI evaluation during pathology work-up**

The following sections provide additional implementation details for the retrospective DFCI evaluation, including clinical-note extraction, diagnostic-timeline reconstruction, reference-cohort construction, and whole-slide image selection.

##### Clinical-note cohort and overview

We retrospectively processed de-identified clinical and pathology notes from 705 adults evaluated at Dana-Farber Cancer Institute (DFCI) for suspected high-grade adult-type diffuse glioma. Across these patients, 8,830 nonempty notes were submitted to the large language model (LLM), including 7,363 progress notes and 1,467 pathology and cytology notes. The 8,830 notes therefore represent the notes supplied to the LLM rather than the complete underlying clinical-note repository.

The extraction pipeline used GPT-4o through the Azure OpenAI Responses API to obtain patient-level, evidence-grounded structured information from the supplied notes. Extracted information included pathology and integrated diagnosis, IDH1 R132H immunohistochemistry (IHC), molecular-testing results, relevant procedures, diagnostic-result dates, and source-note evidence. These extracted results were subsequently consolidated at the patient level to reconstruct diagnostic milestones during routine pathology work-up.

##### LLM implementation and API configuration

We used GPT-4o through the Azure OpenAI Responses API (API version `2025-04-01-preview`). The model temperature was fixed at 0.0 and the maximum output length was 4,000 tokens. Structured JSON output was requested for each patient. The structured-output specification defined the required top-level fields while allowing the detailed nested clinical information described in the prompt template.

The maximum input length used by the pipeline was 80,000 characters. Patient charts exceeding this limit were divided at note boundaries, with a target chunk size of 30,000 characters. Individual notes were not split internally and could therefore exceed the nominal target chunk size.

##### Clinical-note selection

For each patient, an externally recorded tumor sample-collection date was used to define an anchor note. We centered note selection on the tumor sample-collection date to prioritize clinical and pathology documentation surrounding the tissue sampling associated with the H&E WSI used in the downstream analysis, thereby focusing extraction on information most relevant to that specimen. The preferred anchor was the earliest note occurring within the inclusive interval from 7 days before through 60 days after tumor sample collection. If no note occurred within this interval, the chronologically closest dated note was selected, with the earlier note chosen in the event of a tie. The anchor and at most the next 19 notes in chronological order were retained, yielding a maximum of 20 notes per patient. Subsequent retained notes were not required to fall within 60 days after tumor sample collection.

No filtering based on report type or procedure description was applied by the patient-level extraction pipeline. Notes with empty text were excluded. Within each patient, notes were ordered chronologically, with undated notes placed last.

##### Production extraction prompt

The following system prompt was used verbatim for production extraction:

```
You are reviewing de-identified clinical and pathology notes for an
 ↪  adult-type diffuse glioma patient.
```

```
Extract only explicitly documented information from the supplied notes.
Do not infer beyond the notes. Do not use outside medical knowledge to
 ↪  fill missing fields.
Every populated finding/event object must include a short evidence
 ↪  phrase and report_source metadata.
Use the note metadata headers to populate report_source exactly.
Prefer exact dates from report metadata or note text. If only report
 ↪  date is available, use the report date.
If multiple notes disagree, include the most specific
 ↪  evidence-supported value and mention uncertainty in the value or
 ↪  evidence.
Keep the JSON sparse: omit unsupported singleton fields and use empty
 ↪  arrays for unsupported list-type fields.
Treatment entries must use start_date, never date.
Output valid JSON only, with no markdown or commentary.
```

The production user-prompt wrapper was:

```
Review this de-identified patient chart and return sparse
 ↪  evidence-grounded JSON.

Patient ID:
{patient_id}
{chunk_text}
Output schema/template:
{PATIENT_OUTPUT_TEMPLATE}

Rules:
- The patient_id in your JSON must be "{patient_id}".
- Only populate fields with concrete evidence from the supplied notes.
- Every populated object must include evidence and report_source.
- report_source must contain RPT_ID, RPT_DATE_DT, RPT_TYPE, and
 ↪  PROC_DESC from the note metadata header.
- Use result_date for pathology, IHC/histology, molecular,
 ↪  CSF/cytology, and classification result fields.
- Use date for imaging, procedures, supportive/goals-of-care, and
 ↪  timeline events.
- Use start_date for all treatment-related entries. Do not use date
 ↪  inside treatment entries.
- The four top-level likely_* fields are required and must be "yes",
 ↪  "no", or "unclear".
- If evidence is insufficient for a likely_* field, use "unclear".
- Do not include placeholders such as "not reported", "unknown", or
 ↪  generic disease facts.
```

```
Aggregated notes:
<<<NOTES_START>>>
{aggregated_notes}
<<<NOTES_END>>>
```

Each clinical note was supplied to the model using the following representation:

```
===== NOTE {note_number} =====
RPT_ID: {RPT_ID}
RPT_DATE_DT: {timestamp_or_null}
RPT_TYPE: {RPT_TYPE_or_null}
PROC_DESC: {PROC_DESC_or_null}
NOTE_TEXT:
{note_text_deid}
```

For patients processed in multiple chunks, the following instruction was additionally inserted:

```
This is chunk {i} of {n}. Extract only evidence present in this chunk.
```

**Structured output schema**

The structured-output schema required a patient identifier and four top-level chart-classification fields while allowing the detailed nested objects specified in the prompt template:

```
{
  "type": "object",
  "additionalProperties": true,
  "required": [
    "patient_id",
    "likely_newly_sequenced_tumor_for_initial_diagnosis",
    "likely_existing_tumor_profiled_or_reprofiled",
    "likely_nonsequencing_pathology_only",
    "likely_recurrence_or_progression_workup"
  ],
  "properties": {
    "patient_id": {"type": "string"},
    "overall_disease_context": {
      "type": "object",
      "additionalProperties": true
    },
    "initial_tumor_history": {
      "type": "object",
      "additionalProperties": true
    },
    "procedures": {
      "type": "object",
      "additionalProperties": true
```

```
},
"pathology": {
  "type": "object",
  "additionalProperties": true
},
"ihc_and_histology": {
  "type": "object",
  "additionalProperties": true
},
"molecular_testing": {
  "type": "object",
  "additionalProperties": true
},
"csf_and_cytology": {
  "type": "object",
  "additionalProperties": true
},
"imaging_milestones": {
  "type": "object",
  "additionalProperties": true
},
"treatment": {
  "type": "object",
  "additionalProperties": true
},
"supportive_and_goals_of_care": {
  "type": "object",
  "additionalProperties": true
},
"classification": {
  "type": "object",
  "additionalProperties": true
},
"timeline": {
  "type": "array",
  "items": {
    "type": "object",
    "additionalProperties": true
  }
},
"likely_newly_sequenced_tumor_for_initial_diagnosis": {
  "enum": ["yes", "no", "unclear"]
},
"likely_existing_tumor_profiled_or_reprofiled": {
  "enum": ["yes", "no", "unclear"]
},
"likely_nonsequencing_pathology_only": {
  "enum": ["yes", "no", "unclear"]
},
"likely_recurrence_or_progression_workup": {
```

```
      "enum": ["yes", "no", "unclear"]
    }
  }
}
```

The four required top-level `likely_*` fields were included to provide contextual information on the likely nature of tumor sampling, distinguishing newly sequenced tissue obtained for the initial diagnosis, profiling or re-profiling of an existing tumor, non-sequencing pathology evaluation, and recurrence- or progression-related work-up. These fields were retained for descriptive characterization and were not used in cohort inclusion, exclusion, or downstream filtering.

The prompt additionally supplied a detailed nested patient-output template covering diagnoses, procedures, pathology and IHC findings, molecular testing, imaging, treatment, disease classification, and longitudinal timeline events.

Pathology, IHC, molecular, CSF, and classification results were represented using objects of the following general form:

```
{
  "result_date": "YYYY-MM-DD",
  "value": "...",
  "evidence": "...",
  "report_source": [{
    "RPT_ID": "...",
    "RPT_DATE_DT": "...",
    "RPT_TYPE": "...",
    "PROC_DESC": "..."
  }]
}
```

**Chunking and reconciliation of patient-level extractions**

Patient charts containing no more than 80,000 characters were submitted to GPT-4o in a single request. Longer charts were partitioned at note boundaries, with a target chunk size of 30,000 characters. Notes themselves were not subdivided.

Each chunk was independently processed using the same system prompt and patient-level output template. When multiple chunks were required, a second GPT-4o request reconciled the chunk-level JSON output objects. This reconciliation call received the structured chunk outputs but not the original note text. The merge prompt was:

```
Merge the following chunk-level JSON extractions into one final sparse
 ↪  patient-level JSON object.

Patient ID:
{patient_id}
```

```
Output schema/template:
{PATIENT_OUTPUT_TEMPLATE}

Rules:
- Preserve report_source metadata from the chunk outputs.
- Keep only evidence-grounded findings/events.
- Remove duplicates when the same finding/event is represented multiple
 ↪  times.
- Prefer the most specific evidence-supported value when chunk outputs
 ↪  conflict.
- Treatment entries must use start_date, never date.
- The four top-level likely_* fields are required and must be "yes",
 ↪  "no", or "unclear".
- Output valid JSON only.

Chunk-level JSON outputs:
{chunk_outputs}
```

No deterministic voting procedure or date-based tie-break was applied when chunk outputs disagreed. Instead, the reconciliation prompt instructed GPT-4o to retain the most specific evidence-supported value.

If an extracted object was malformed or rejected by downstream validation, one additional GPT-4o repair call was permitted at temperature 0. The repair procedure was restricted to information already represented in the draft extraction and was not permitted to introduce new clinical information. All final patient-level JSON outputs passed downstream validation.

**Extraction and consolidation of diagnostic milestones**

After LLM extraction, diagnostic milestones were consolidated deterministically at the patient level.

For IDH1 R132H IHC, candidate events included the dedicated `idh1_r132h_ihc` result and other marker results referring to IDH together with IHC or R132H. The earliest qualifying result date was retained.

For molecular testing, candidate dates included clinical OncoPanel [9] results, cytogenetic or array-CGH results, MGMT-promoter methylation results, and other molecular tests. The earliest qualifying molecular-result date was retained.

For integrated diagnosis, candidate events included the directly extracted integrated-diagnosis result and qualifying pathology, molecular, timeline, or overall-disease-context entries. The earliest qualifying integrated-diagnosis result date and associated disease status were retained.

For surgery or biopsy, the earliest brain surgery or biopsy date, qualifying surgical timeline event, or matching relevant procedure was retained.

**Clinician validation of extracted diagnostic information**

To assess the accuracy of pathology-report extraction, 30 patient cases were randomly selected from the clinician-review-eligible cohort ($n$ = 282), defined as patients with available integrated-diagnosis, IDH-status, and molecular-result dates. Sampling was stratified by integrated diagnosis, with at least five cases from each less common diagnosis disease status, and subsequently adjusted by random replacement to ensure inclusion of 10 IDH-mutant patients. The final sample comprised 20 glioblastoma, IDH-wildtype, 5 glioblastoma, IDH-mutant, and 5 anaplastic oligodendroglioma, IDH-mutant and 1p/19q-codeleted cases. By IDH status, the same 30 cases comprised 20 IDH-wildtype and 10 IDH-mutant patients. A board-certified clinician at the Dana-Farber Cancer Institute, blinded to the LLM outputs, reviewed the same clinical records provided to the extraction pipeline and independently assessed the extracted disease statuses and associated result dates. Agreement was evaluated for categorical disease status and by calendar-day differences between LLM-extracted and clinician-reviewed dates. Disagreements were subsequently reviewed unblinded and adjudicated. The review results for this validation are provided in the Supplementary Materials.

**Reference-cohort construction**

The retrospective DFCI source cohort comprised 705 patients whose clinical records underwent the LLM-based extraction procedure described above. For the integrated-diagnosis evaluation, we defined three eligible disease statuses based on WHO 2016 guideline [7]. These disease statuses were selected because the downstream pathology analysis focused on clinically aggressive gliomas in which high-grade histomorphologic features, including microvascular proliferation and necrosis, are diagnostically and clinically relevant:

- glioblastoma, IDH-wildtype, WHO grade IV;
- glioblastoma, IDH-mutant, WHO grade IV; and
- anaplastic oligodendroglioma, IDH-mutant and 1p/19q-codeleted, WHO grade III.

The task-specific reference-cohort flows were:

- Integrated diagnosis: (705 → 359 → 350).
- IDH restricted: (705 → 359 → 357 → 348).

Here, 359 denotes patients with an eligible integrated-diagnosis disease status. 350 denotes those patients with a retained available H&E WSI. 357 denotes patients with both an eligible integrated diagnosis and a molecular IDH reference label. Finally, 348 denotes those patients with a retained available H&E WSI.

Thus, inclusion in the integrated-diagnosis cohort required an eligible integrated-diagnosis disease status and an available H&E WSI, whereas inclusion in the IDH-status cohort additionally required a molecular IDH reference label. The exact patient-and-slide intersection of the two cohorts comprised 348 patients.

##### Molecular IDH reference status

IDH immunohistochemistry (IHC) results extracted from neuropathology reports using the LLM were used only to reconstruct the diagnostic timeline, specifically to estimate the time from tissue collection to availability of an IDH result and the corresponding potential time savings in the neuropathology work-up. In contrast, evaluation of StratCP error rates used an independent molecular reference label derived from OncoPanel sequencing [9] of the same tissue block from which the H&E slide used as foundation-model input was obtained. OncoPanel results were mapped to IDH-mutant or IDH-wildtype status. A negative IDH IHC result alone was therefore not treated as evidence of molecular IDH-wildtype status. This separation allowed clinical-note-derived IHC information to inform diagnostic timing while preserving an independent molecular ground truth for StratCP evaluation.

##### Harmonization of integrated diagnosis

LLM-extracted integrated diagnoses were normalized using a curated diagnosis-mapping procedure and subsequently mapped to the three prespecified high-grade adult-type diffuse glioma disease statuses listed above. For consistency with the diagnostic framework used for the retrospective evaluation, endpoint disease statuses are reported using WHO 2016 terminology [7].

Of the 705 source patients, 359 satisfied the prespecified integrated-diagnosis endpoint. These included:

- 298 patients with glioblastoma, IDH-wildtype, WHO grade IV;
- 32 patients with glioblastoma, IDH-mutant, WHO grade IV; and
- 29 patients with anaplastic oligodendroglioma, IDH-mutant and 1p/19q-codeleted, WHO grade III.

Of the remaining 346 patients, 227 had no available harmonized integrated diagnosis and 119 had an extracted diagnosis that did not map to one of the three prespecified disease statuses. Among the latter were 21 diffuse astrocytomas, IDH-mutant, WHO grade II; 21 anaplastic astrocytomas, IDH-mutant, WHO grade III; 16 oligodendrogliomas, IDH-mutant and 1p/19q-codeleted, WHO grade II; and 61 patients with other non-endpoint diagnoses, including diagnoses with unavailable grade, NOS/NEC designations, pediatric-type or glioneuronal tumors, H3-altered tumors, or other disease statuses outside the prespecified high-grade endpoint.

Of the 359 patients satisfying the integrated-diagnosis endpoint, two lacked an external molecular IDH reference label, leaving 357 patients with both an eligible integrated diagnosis and molecular IDH reference status.

##### Whole-slide image linkage

Pathology-image metadata contained patient, specimen, block, requisition, and slide-accession identifiers. Patient-level linkage was performed using DFCI patient identifiers. Source patient identifiers were converted to randomized identifiers for analysis and internally namespaced to preserve uniqueness.

Among the 359 patients satisfying the integrated-diagnosis endpoint, 9 had no retained available WSI after slide-availability filtering, yielding 350 patients for the integrated-diagnosis cohort. Among the 357 patients additionally having a molecular IDH reference label, 9 had no retained available WSI, yielding 348 patients for the IDH-status cohort.

**Representative tumor sample date and one-slide-per-patient selection**

For deterministic selection of one representative WSI per patient, we first defined a representative tumor sample date. When an LLM-extracted surgery or biopsy date occurred within six months of the externally recorded tumor sample-collection date, the extracted surgery or biopsy date was used; otherwise, the external tumor sample-collection date was retained. We prioritized the clinically documented surgery or biopsy date when temporally concordant because it directly reflects the procedure associated with tissue acquisition in the clinical record, whereas the external sample-collection date was derived independently from structured specimen metadata. The six-month concordance window was used to avoid assigning an extracted procedure from a different episode of tumor sampling to the WSI under consideration.

Across the 705 patients, 419 used the extracted surgery or biopsy date and 286 used the external tumor sample-collection date. Among the latter, 213 patients had an extracted surgery date that fell outside the six-month concordance window and therefore retained the external tumor sample-collection date.

Candidate slides were then ordered according to the following deterministic priorities:

1. primary-tumor samples before non-primary samples;
2. smallest absolute difference between the WSI procedure date and tumor sample-collection date; and
3. alphabetically smaller slide accession as the final tie-break.

9 patients had 2 candidate slides. Deterministic selection therefore removed 9 duplicate slide records but did not remove any additional patients.

**Final reference cohorts**

The resulting integrated-diagnosis cohort comprised 350 patients with one selected WSI per patient: 292 glioblastomas, IDH-wildtype, WHO grade IV; 30 glioblastomas, IDH-mutant, WHO grade IV; and 28 anaplastic oligodendrogliomas, IDH-mutant and 1p/19q-codeleted, WHO grade III.

The IDH-status cohort comprised 348 patients with one selected WSI per patient, including 290 IDH-wildtype and 58 IDH-mutant patients. The exact patient-and-slide intersection of the integrated-diagnosis and IDH-status cohorts also comprised 348 patients. Within this paired cohort, the integrated-diagnosis disease-status distribution was 290 glioblastomas, IDH-wildtype, WHO grade IV; 30 glioblastomas, IDH-mutant, WHO grade IV; and 28 anaplastic oligodendrogliomas, IDH-mutant and 1p/19q-codeleted, WHO grade III.

Valid milestone dates were additionally required for analyses involving the corresponding diagnostic intervals but were not required for inclusion in either reference cohort.

**Bootstrap uncertainty estimation**

Because the retrospective DFCI analysis used a single prespecified temporal split rather than repeated random calibration/test splits, selected error was reported directly on the held-out temporal test cohort rather than as an FDR averaged over repetitions. For each task, method, and predicted disease status, uncertainty in both the selected error rate and the number of selected patients was quantified using 2,000 patient-level bootstrap replicates sampled with replacement. Error bars show $\pm 1$ bootstrap standard error, computed as the sample standard deviation of the finite bootstrap estimates across replicates. Replicates containing no selected patients were excluded when estimating the standard error of the selected error rate.

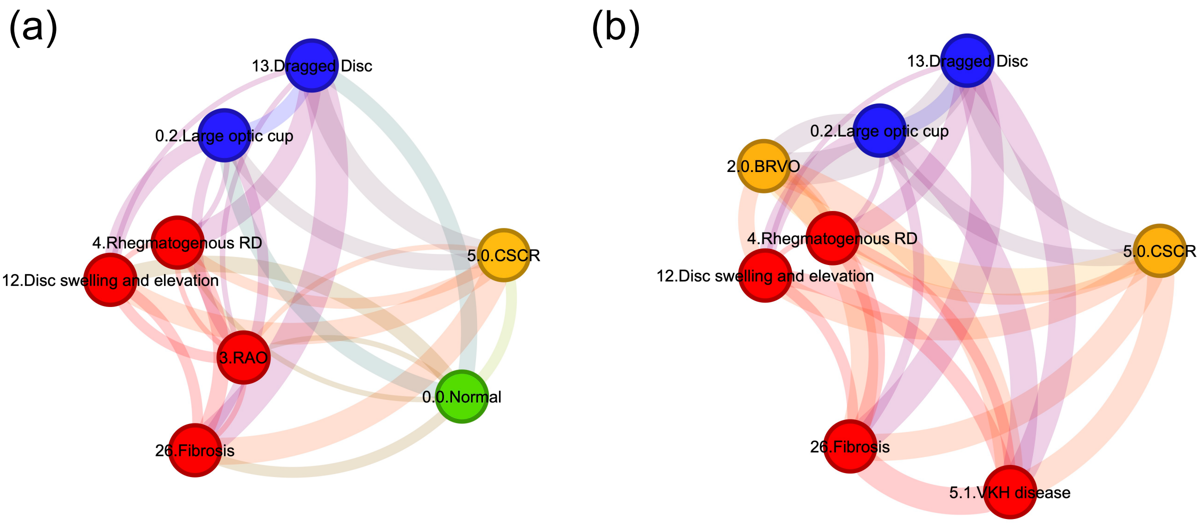


**Figure S1: Example prediction sets for a deferred patient from StratCP in eye condition diagnosis. a,** Standard APS-based prediction set for an example patient deferred by StratCP. Each node represents a candidate disease status, with node color indicating clinical urgency. Edges connect disease statuses with overlapping downstream management actions, with edge width proportional to the number of shared management actions. **b,** Utility-enhanced prediction set for the same patient. Incorporating management-action utility changes the composition of the prediction set toward candidate disease statuses with greater overlap in their downstream management actions. Node colors and edge widths are defined as in **a**.

**Table S1:** Sample counts and FM performance by true and predicted disease stage classes for Diabetic Retinopathy Diagnosis task. The sample size combines test and calibration folds. Each sample corresponds to a unique patient.

| Class | Sample Size (By True Class) | FM Accuracy (By True Class) | Sample Size (By Predicted Class) | FM Accuracy (By Predicted Class) |
|---|---|---|---|---|
| Normal | 542 | 97.23% | 533 | 98.87% |
| Mild | 111 | 48.65% | 76 | 71.05% |
| Moderate | 300 | 91.67% | 390 | 70.51% |
| Severe | 58 | 34.48% | 45 | 44.44% |
| Proliferative | 89 | 50.56% | 56 | 80.36% |
| **All** | **1100** | **83.72%** | **1100** | **83.72%** |

**Table S2:** Sample counts and FM performance by true and predicted disease stage classes for Glaucoma Diagnosis task. The sample size combines test and calibration folds. Each sample corresponds to a unique patient.

| Class | Sample Size (By True Class) | FM Accuracy (By True Class) | Sample Size (By Predicted Class) | FM Accuracy (By Predicted Class) |
|---|---|---|---|---|
| Mild | 237 | 91.56% | 244 | 88.93% |
| Early | 87 | 45.98% | 54 | 74.07% |
| Advanced | 141 | 98.58% | 167 | 83.23% |
| **All** | **465** | **85.16%** | **465** | **85.16%** |

**Table S3:** Sample counts and FM performance by true and predicted eye condition classes for Eye Condition Diagnosis task. The sample size combines test and calibration folds. Each sample corresponds to a unique patient.

| Class | Sample Size (By True Class) | FM Accuracy (By True Class) | Sample Size (By Predicted Class) | FM Accuracy (By Predicted Class) |
|---|---|---|---|---|
| Normal | 12 | 58.33% | 7 | 100.00% |
| Tessellated fundus | 4 | 100.00% | 4 | 100.00% |
| Large optic cup | 15 | 86.67% | 19 | 68.42% |
| DR1 (Mild non-proliferate diabetic retinopathy) | 6 | 100.00% | 8 | 75.00% |
| DR2 (Moderate non-proliferate diabetic retinopathy) | 15 | 100.00% | 18 | 83.33% |
| DR3 (Severe non-proliferate diabetic retinopathy) | 12 | 58.33% | 9 | 77.78% |
| Possible glaucoma | 4 | 50.00% | 2 | 100.00% |
| Optic atrophy | 4 | 75.00% | 3 | 100.00% |
| Severe hypertensive retinopathy | 5 | 80.00% | 4 | 100.00% |
| Disc swelling and elevation | 4 | 100.00% | 5 | 80.00% |
| Dragged Disc | 3 | 100.00% | 3 | 100.00% |
| Congenital disc abnormality | 3 | 0.00% | 3 | 0.00% |
| Retinitis pigmentosa | 7 | 85.71% | 6 | 100.00% |
| Bietti crystalline dystrophy | 3 | 100.00% | 3 | 100.00% |
| Peripheral retinal degeneration and break | 5 | 80.00% | 5 | 80.00% |
| Myelinated nerve fiber | 4 | 100.00% | 6 | 66.67% |
| Vitreous particles | 5 | 80.00% | 4 | 100.00% |
| Fundus neoplasm | 3 | 0.00% | 1 | 0.00% |
| BRVO (Branch retinal vein occlusion) | 14 | 92.86% | 13 | 100.00% |
| CRVO (Central retinal vein occlusion) | 7 | 100.00% | 7 | 100.00% |
| Massive hard exudates | 4 | 75.00% | 3 | 100.00% |
| Yellow-white spots-flecks | 9 | 66.67% | 7 | 85.71% |
| Cotton-wool spots | 3 | 100.00% | 3 | 100.00% |
| Vessel tortuosity | 5 | 60.00% | 3 | 100.00% |
| Chorioretinal atrophy-coloboma | 5 | 60.00% | 3 | 100.00% |
| Preretinal hemorrhage | 3 | 100.00% | 3 | 100.00% |
| Fibrosis | 3 | 0.00% | 1 | 0.00% |
| Laser Spots | 6 | 100.00% | 7 | 85.71% |
| Silicon oil in eye | 6 | 83.33% | 5 | 100.00% |
| Blur fundus without PDR | 34 | 100.00% | 36 | 94.44% |
| Blur fundus with suspected PDR | 14 | 100.00% | 14 | 100.00% |
| RAO (Retinal artery occlusion) | 5 | 80.00% | 4 | 100.00% |
| Rhegmatogenous RD | 18 | 94.44% | 21 | 80.95% |
| CSCR (Central serous chorioretinopathy) | 5 | 80.00% | 7 | 57.14% |
| VKH disease | 5 | 100.00% | 6 | 83.33% |
| Maculopathy | 23 | 100.00% | 28 | 82.14% |
| ERM (Epiretinal membrane) | 8 | 87.50% | 8 | 87.50% |
| MH (Macular hole) | 7 | 85.71% | 8 | 75.00% |
| Pathological myopia | 17 | 100.00% | 18 | 94.44% |
| **All** | **315** | **86.35%** | **315** | **86.35%** |

**Table S4:** Slide counts and held-out performance (precision and recall) by cohort and IDH molecular class for the IDH mutation status prediction task. Development combines training and validation, while held-out combines test and calibration. "All (Combined)" denotes support-weighted performance across IDH statuses after pooling both cohorts.

| Cohort | Molecular class | Development (train+val) | Held-out (test+calib) | Precision | Recall |
|---|---|---|---|---|---|
| EBRAINS | IDH-wildtype | 351 | 189 | 0.949 | 0.878 |
| | IDH-mutant | 218 | 115 | 0.822 | 0.922 |
| TCGA LGG/GBM | IDH-wildtype | 415 | 191 | 0.884 | 0.916 |
| | IDH-mutant | 526 | 241 | 0.932 | 0.905 |
| | **All (Combined)** | **1,510** | **736** | **0.904** | **0.904** |

**Table S5:** Cross-cohort performance of the fixed UNI-based IDH mutation status predictor. This analysis uses a different data split from Table S4: the predictor was developed using TCGA LGG/GBM slides only and evaluated on an independent TCGA held-out set and the full eligible EBRAINS target cohort, without retraining or model selection on EBRAINS. Development combines training and validation slides. Precision, recall, and F1 are reported separately for IDH-wildtype and IDH-mutant statuses. "All (Combined)" denotes support-weighted performance across statuses.

| Cohort | IDH status | Development (train+val) | Held-out | Precision | Recall | F1 |
|---|---|---|---|---|---|---|
| TCGA LGG/GBM | IDH-wildtype | 390 | 216 | 0.845 | 0.861 | 0.853 |
| | IDH-mutant | 492 | 275 | 0.889 | 0.876 | 0.883 |
| | **All (Combined)** | **882** | **491** | **0.870** | **0.870** | **0.870** |
| EBRAINS | IDH-wildtype | – | 540 | 0.921 | 0.869 | 0.894 |
| | IDH-mutant | – | 333 | 0.805 | 0.880 | 0.841 |
| | **All (Combined)** | – | **873** | **0.877** | **0.873** | **0.874** |

**Table S6:** Cross-cohort discrimination performance of the fixed UNI-based IDH mutation status predictor. Weighted area under the receiver operating characteristic curve (AUROC) and area under the precision–recall curve (AUPRC) are reported for the independent TCGA LGG/GBM holdout cohort and the full eligible EBRAINS target cohort. The predictor was developed using TCGA LGG/GBM slides only and was applied to EBRAINS without retraining or model selection.

| Cohort | Evaluation slides | Weighted AUROC | Weighted AUPRC |
|---|---|---|---|
| TCGA LGG/GBM | 491 | 0.951 | 0.954 |
| EBRAINS | 873 | 0.941 | 0.936 |

**Table S7:** Slide counts by WHO 2016–based central nervous system (CNS) diagnosis [7] for the CNS Tumor Subtyping task (EBRAINS only), along with abbreviations used in the utility-graph schematic (Extended Data Fig. 1) and held-out performance (precision and recall) for each subtype. The development set combines training and validation, while the held-out set combines test and calibration. "All (Combined)" denotes support-weighted performance across disease statuses. A period ( . ) denotes diagnoses without a unique class-level WHO grade for the grade-based utility analysis. Haemangiopericytoma was treated as ungraded because samples with this reference diagnosis spanned WHO grades I–III in the source metadata.

| WHO 2016–based CNS diagnosis | Abbrev. | Grade | Development (train+val) | Held-out (test+calib) | Precision | Recall |
|---|---|---|---|---|---|---|
| Glioblastoma, IDH-wildtype | GliBW | IV | 310 | 164 | 0.846 | 0.872 |
| Pilocytic astrocytoma | PilAst | I | 115 | 58 | 0.895 | 0.879 |
| Meningothelial meningioma | MenTel | I | 66 | 38 | 0.531 | 0.447 |
| Pituitary adenoma | PitAd | . | 64 | 35 | 0.971 | 0.971 |
| Anaplastic oligodendroglioma, IDH-mutant and 1p/19q codeleted | AnODG | III | 57 | 34 | 0.576 | 0.559 |
| Ganglioglioma | Gangl | I | 57 | 31 | 0.730 | 0.871 |
| Haemangioblastoma | HaemBl | I | 58 | 30 | 0.938 | 1.000 |
| Adamantinomatous craniopharyngioma | AdamCr | I | 55 | 30 | 1.000 | 0.933 |
| Oligodendroglioma, IDH-mutant and 1p/19q codeleted | OligDG | II | 58 | 27 | 0.515 | 0.630 |
| Atypical meningioma | AtyMen | II | 54 | 29 | 0.379 | 0.379 |
| Schwannoma | Schwan | I | 52 | 29 | 0.824 | 0.966 |
| Diffuse astrocytoma, IDH-mutant | DifAst | II | 41 | 29 | 0.833 | 0.517 |
| Transitional meningioma | TraMen | I | 43 | 25 | 0.367 | 0.440 |
| Diffuse large B-cell lymphoma of the CNS | CNSLym | . | 39 | 20 | 1.000 | 0.950 |
| Gliosarcoma | Gliosa | IV | 34 | 25 | 0.636 | 0.560 |
| Fibrous meningioma | FibMen | I | 36 | 21 | 0.563 | 0.429 |
| Anaplastic ependymoma | AnEpen | III | 33 | 17 | 0.688 | 0.647 |
| Metastatic tumours | Metast | . | 31 | 16 | 0.667 | 0.750 |
| Anaplastic astrocytoma, IDH-wildtype | AnAstW | III | 29 | 18 | 0.263 | 0.278 |
| Anaplastic astrocytoma, IDH-mutant | AnAstM | III | 29 | 18 | 0.412 | 0.389 |
| Ependymoma | Epend | II | 29 | 17 | 0.600 | 0.529 |
| Anaplastic meningioma | AnMen | III | 31 | 15 | 0.368 | 0.467 |
| Secretory meningioma | SecMen | I | 26 | 15 | 0.800 | 0.800 |
| Lipoma | Lipom | . | 25 | 13 | 0.733 | 0.846 |
| Haemangiopericytoma | HaemPC | . | 18 | 16 | 0.733 | 0.688 |
| Glioblastoma, IDH-mutant | GliBM | IV | 22 | 12 | 0.500 | 0.500 |
| Medulloblastoma, non-WNT/non-SHH | MednSH | IV | 21 | 11 | 0.917 | 1.000 |
| Langerhans cell histiocytosis | LCH | . | 21 | 11 | 0.900 | 0.818 |
| Angiomatous meningioma | AngMen | I | 20 | 11 | 1.000 | 0.818 |
| Haemangioma | HaemAn | . | 20 | 10 | 0.889 | 0.800 |
| **All (Combined)** | – | | **1494** | **825** | **0.733** | **0.728** |

**Table S8:** Held-out performance of CLMBR-T-base–based predictors across longitudinal EHR tasks. Task-specific predictors were trained on frozen CLMBR-T-base representations using the predefined EHRSHOT development partitions [2] and evaluated on the held-out test partition. Development combines training and validation examples. For each task, disease-status-specific rows report the number of examples with the corresponding true outcome in the development and held-out sets, together with held-out precision, recall, and F1 score. "All (Combined)" denotes support-weighted performance across disease statuses within each task.

| Task | Disease status | Development (train+val) | Held-out | Precision | Recall | F1 |
|---|---|---|---|---|---|---|
| Length of stay | Short LOS | 1,982 | 936 | 0.914 | 0.746 | 0.821 |
| | Long LOS | 635 | 302 | 0.498 | 0.781 | 0.608 |
| | **All (Combined)** | **2,617** | **1,238** | **0.812** | **0.754** | **0.769** |
| Pancreatic cancer diagnosis | No pancreatic cancer | 2,444 | 1,206 | 0.990 | 0.789 | 0.878 |
| | Pancreatic cancer | 174 | 40 | 0.105 | 0.750 | 0.185 |
| | **All (Combined)** | **2,618** | **1,246** | **0.961** | **0.787** | **0.855** |
| Thrombocytopenia severity | Normal | 3,359 | 1,645 | 0.851 | 0.953 | 0.899 |
| | Mild | 413 | 213 | 0.179 | 0.070 | 0.101 |
| | Moderate | 150 | 90 | 0.227 | 0.056 | 0.089 |
| | Severe | 143 | 50 | 0.520 | 0.520 | 0.520 |
| | **All (Combined)** | **4,065** | **1,998** | **0.743** | **0.808** | **0.768** |

**Table S9:** Discrimination performance of CLMBR-T-base–based predictors across longitudinal EHR tasks. Weighted area under the receiver operating characteristic curve (AUROC) and area under the precision–recall curve (AUPRC) are reported on the held-out EHRSHOT test partition [2]. Class-specific AUROC and AUPRC were computed in a one-vs-rest manner, and the values reported here are support-weighted averages across disease statuses within each task.

| Task | Held-out examples | Weighted AUROC | Weighted AUPRC |
|---|---|---|---|
| Length of stay | 1,238 | 0.840 | 0.864 |
| Pancreatic cancer diagnosis | 1,246 | 0.797 | 0.962 |
| Thrombocytopenia severity | 1,998 | 0.765 | 0.808 |

**Table S10:** The number of patients by WHO grade and follow-up status, along with held-out performance in concordance index (C-idx). Only the TCGA dataset (LGG and GBM cohorts) is used for this task. For this analysis, one slide was used per patient. When multiple slides were available, one was selected at random. The development set combines training and validation, while the held-out set combines test and calibration. Molecular diagnoses are identified using available molecular data following the approach described in [5]. Grade 4 corresponds to the following diagnoses: "Glioblastoma, IDH-wildtype. Grade 4" and "Astrocytoma, IDH-mutant. Grade 4." The Grade 2 & 3 group corresponds to the following diagnoses: "Astrocytoma, IDH-mutant. Grade 2," "Astrocytoma, IDH-mutant. Grade 3," "Oligodendroglioma, IDH-mutant, 1p/19q-codeleted. Grade 3," and "Oligodendroglioma, IDH-mutant, 1p/19q-codeleted. Grade 2." WHO grade unclassified denotes cases with nonspecific diagnoses that could not be mapped to a WHO grade.

| WHO Grade | Follow-up status | Development (train+val) | Held-out (test+calib) | C-idx |
|---|---|---|---|---|
| 4 | Follow-up > 18 months | 76 | 32 | 0.631 |
| | ≤ 18 months | 137 | 72 | |
| 2 & 3 | Follow-up > 18 months | 142 | 76 | 0.557 |
| | ≤ 18 months | 74 | 42 | |
| WHO grade unclassified | Follow-up > 18 months | 50 | 36 | 0.700 |
| | ≤ 18 months | 87 | 49 | |
| **All (Combined)** | | **566** | **307** | 0.767 |

**Table S11:** Slide counts by pathology feature group, cohort, and diagnosis for the H&E-only diagnosis task in adult-type diffuse glioma, along with per-diagnosis performance within each pathology feature group. A given diagnosis may occur in more than one pathology feature group. Each feature group subset is split separately into development and held-out data and therefore may have different split counts. The development set combines training and validation data, whereas the held-out set combines test and calibration data. "All (Combined)" denotes support-weighted performance across disease statuses.

| Pathology feature group | Cohort | Diagnosis | Development (train+val) | Held-out (test+calib) | Precision | Recall |
|---|---|---|---|---|---|---|
| Microvascular proliferation (MVP) or necrosis present | EBRAINS | Anaplastic oligodendroglioma, IDH-mutant, 1p/19q codeleted | 60 | 31 | 0.788 | 0.839 |
| | EBRAINS | Glioblastoma, IDH-mutant | 21 | 13 | 0.538 | 0.538 |
| | EBRAINS | Glioblastoma, IDH-wildtype | 310 | 164 | 0.969 | 0.957 |
| | TCGA LGG/GBM | Anaplastic oligodendroglioma, IDH-mutant, 1p/19q codeleted | 67 | 44 | 0.968 | 0.682 |
| | TCGA LGG/GBM | Glioblastoma, IDH-mutant | 39 | 20 | 0.688 | 0.550 |
| | TCGA LGG/GBM | Glioblastoma, IDH-wildtype | 349 | 191 | 0.894 | 0.974 |
| **All (Combined)** | | | **846** | **463** | 0.897 | 0.901 |
| Mitotic activity present, without MVP/necrosis | EBRAINS | Anaplastic astrocytoma, IDH-mutant | 34 | 13 | 0.692 | 0.692 |
| | EBRAINS | Anaplastic astrocytoma, IDH-wildtype | 30 | 17 | 0.706 | 0.706 |
| | EBRAINS | Anaplastic oligodendroglioma, IDH-mutant, 1p/19q codeleted | 66 | 25 | 0.880 | 0.880 |
| | TCGA LGG/GBM | Anaplastic astrocytoma, IDH-mutant | 45 | 30 | 0.871 | 0.900 |
| | TCGA LGG/GBM | Anaplastic oligodendroglioma, IDH-mutant, 1p/19q codeleted | 51 | 60 | 0.966 | 0.933 |
| **All (Combined)** | | | **226** | **145** | 0.872 | 0.869 |
| Neither MVP/necrosis nor mitotic activity | EBRAINS | Diffuse astrocytoma, IDH-mutant | 46 | 24 | 0.880 | 0.917 |
| | EBRAINS | Oligodendroglioma, IDH-mutant, 1p/19q codeleted | 54 | 31 | 0.933 | 0.903 |
| | TCGA LGG/GBM | Diffuse astrocytoma, IDH-mutant | 88 | 33 | 0.703 | 0.788 |
| | TCGA LGG/GBM | Oligodendroglioma, IDH-mutant, 1p/19q codeleted | 54 | 65 | 0.885 | 0.831 |
| **All (Combined)** | | | **242** | **153** | 0.854 | 0.850 |

**Table S12:** Patient counts and predictive performance in the retrospective DFCI pathology cohorts. Metrics were computed across the complete calibration-plus-test cohorts using the reference labels and fixed model predictions. Each patient contributed one H&E whole-slide image. Patient count denotes true-class support. Precision and recall are reported for each disease status, and "All (Combined)" denotes support-weighted performance across disease statuses.

| Task | Disease status | Patient count (test+calib) | Precision | Recall |
|---|---|---|---|---|
| Integrated diagnosis | Anaplastic oligodendroglioma, IDH-mutant and 1p/19q-codeleted | 28 | 0.917 | 0.393 |
| | Glioblastoma, IDH-mutant | 30 | 0.500 | 0.433 |
| | Glioblastoma, IDH-wildtype | 292 | 0.913 | 0.976 |
| | **All (Combined)** | **350** | **0.878** | **0.883** |
| IDH status | IDH-mutant | 58 | 0.829 | 0.586 |
| | IDH-wildtype | 290 | 0.922 | 0.976 |
| | **All (Combined)** | **348** | **0.906** | **0.911** |

**Table S13: De-identified DFCI reference cohorts and clinician validation data.** Provided as a separate Excel file (`Supplementary_Table_13.xlsx`). The workbook contains three worksheets: `Clinician_agreement_review`, containing case-level LLM–clinician agreement and adjudication results for the 30 reviewed patients, `Integrated diagnosis`, containing randomized patient identifiers and reference integrated-diagnosis disease statuses for the 350-patient cohort, and `IDH status`, containing randomized patient identifiers and reference IDH-status disease statuses for the 348-patient cohort. All patient identifiers in the workbook are randomized identifiers used for the study analyses.

#### Supplementary references


1. Chen, R. J. *et al.* Towards a general-purpose foundation model for computational pathology. *Nature medicine* **30**, 850–862 (2024).

2. Wornow, M., Thapa, R., Steinberg, E., Fries, J. & Shah, N. Ehrshot: An ehr benchmark for few-shot evaluation of foundation models. *Advances in Neural Information Processing Systems* **36**, 67125–67137 (2023).

3. Angelopoulos, A. N., Bates, S., Candès, E. J., Jordan, M. I. & Lei, L. Learn then test: Calibrating predictive algorithms to achieve risk control. *The Annals of Applied Statistics* **19**, 1641–1662 (2025).

4. Hoeffding, W. Probability inequalities for sums of bounded random variables. *Journal of the American statistical association* **58**, 13–30 (1963).

5. Zakharova, G. *et al.* Reclassification of tcga diffuse glioma profiles linked to transcriptomic, epigenetic, genomic and clinical data, according to the 2021 who cns tumor classification. *International journal of molecular sciences* **24**, 157 (2022).

6. Roetzer-Pejrimovsky, T. *et al.* The digital brain tumour atlas, an open histopathology resource. *Scientific Data* **9**, 55 (2022).

7. Louis, D. N. *et al.* The 2016 world health organization classification of tumors of the central nervous system: a summary. *Acta neuropathologica* **131**, 803–820 (2016).

8. Davidson-Pilon, C. lifelines: survival analysis in python. *Journal of Open Source Software* **4**, 1317 (2019).

9. Garcia, E. P. *et al.* Validation of oncopanel: a targeted next-generation sequencing assay for the detection of somatic variants in cancer. *Archives of pathology & laboratory medicine* **141**, 751–758 (2017).
